# Modeling the interplay between disease spread, behaviors, and disease perception with a data-driven approach

**DOI:** 10.1101/2024.04.10.24305600

**Authors:** Alessandro De Gaetano, Alain Barrat, Daniela Paolotti

**Author notes:** A.B. and D.P. contributed equally to this work.

## Abstract

Individuals’ perceptions of disease influence their adherence to preventive measures, shaping the dynamics of disease spread. Despite extensive research on the interaction between disease spread, human behaviors, and interventions, few models have incorporated real-world behavioral data on disease perception, limiting their applicability. This study novelly integrates disease perception, represented by perceived severity, as a critical determinant of behavioral change into a data-driven compartmental model to assess its impact on disease spread. Using survey data, we explore scenarios involving a competition between a COVID-19 wave and a vaccination campaign, where individuals’ behaviors vary based on their perceived severity of the disease. Results demonstrate that behavioral heterogeneities influenced by perceived severity affect epidemic dynamics, with high heterogeneity yielding contrasting effects. Longer adherence to protective measures by groups with high perceived severity provides greater protection to vulnerable individuals, while premature relaxation of behaviors by low perceived severity groups facilitates virus spread. Epidemiological curves reveal that differences in behavior among groups can eliminate a second infection peak, resulting in a higher first peak and overall more severe outcomes. The specific modeling approach for how perceived severity modulates behavior parameters does not strongly impact the model’s outcomes. Sensitivity analyses confirm the robustness of our findings, emphasizing the consistent impact of behavioral heterogeneities across various scenarios. Our study underscores the importance of integrating risk perception into infectious disease transmission models and highlights the necessity of extensive data collection to enhance model accuracy and relevance.

## 1 Introduction

The propagation patterns of infectious diseases are shaped by human interactions, movements, and individual conduct. Reciprocally, the dynamic unfolding of contagious illnesses can impact human behavior [1, 2, 3]. Therefore, the interplay between disease spreading, human behaviors, and interventions, both pharmaceutical and non-pharmaceutical, has been largely studied in literature during the last twenty years (for reviews see [3, 4, 5]). Several works have integrated these different elements in various mathematical modeling frameworks, to gain quantitative insights and provide predictions and projections. Nonetheless, most of these models were limited to theoretical investigations and were not informed by representative, real-world, timely data on the behavioral aspects, largely due to the lack of availability of such data. This situation has however evolved since the emergence of SARS-CoV-2 and the resulting COVID-19 pandemic, which stimulated important data collection efforts to better inform models and gain insights into the spread of SARS-CoV-2 at various scales [6, 7, 8, 9, 10]. In particular, numerous studies have focused during this pandemic on evaluating the effectiveness of Non-Pharmaceutical Interventions (NPIs) and government-imposed restrictions to mitigate the contagion [11, 12, 13, 14, 15], as well as on assessing the benefits of vaccination campaigns [16, 17, 18].

In the current post-pandemic period, however, top-down emergency measures have been discontinued, and the responsibility for adopting protective measures is left to individuals. Possible protective measures encompass both aspects of each individual’s social life, such as reducing social gatherings, and hygiene- or health-related practices, including mask-wearing and vaccination decisions [19]. Disease perception, i.e., the way individuals perceive how the disease might impact them, plays an important role in the adoption of such protective behaviors as evidenced by numerous studies and taken into account in psychological models (e.g. Health Belief Model) [20, 21, 22]. For instance, individuals who perceive a higher risk are more likely to adopt recommended hygiene and avoidance behaviors [23], and this relationship strengthens throughout an epidemic [24]. However, and despite the vast literature that proves its influence on behavioral aspects [23, 24, 25, 26, 27], risk perception has rarely been included in data-driven modeling frameworks in particular for lack of data. Consequently, the collection of data on disease perception and social contacts during the pandemic presents an invaluable opportunity to investigate the impact of risk-perception-driven behaviors and behavioral changes on the propagation of a disease in a population.

In this paper, we leverage data collected during the pandemic to build a data-driven mathematical modeling framework and investigate the complex relationship between self-adopted protective behaviors, disease spreading, and risk perception. In particular, the scenario we envision presents a competition between an ongoing wave of COVID-19 and a vaccination campaign. We however consider a context similar to the current post-pandemic period where top-down emergency measures are absent, and individuals are responsible for their protection. In this context, we develop a deterministic compartmental model that incorporates a feedback loop between behaviors, vaccines, and the contagion process and that includes risk perception as a key determinant of the adoption or relaxation of individual protective behaviors.

To inform the model, we use data from the CoMix survey [7] along two different lines. On the one hand, we model different levels of compliance with protective behaviors by two different contact matrices giving the average number of contacts between individuals of different age groups. On the other hand, we stratify the population according not only to age classes but also to how individuals perceive whether the disease poses a threat to them. Specifically, we classify individuals according to their “perceived severity”, i.e., on their belief of the seriousness of the disease if they were to catch it. Perceived severity has indeed been found to be one of the most important factors impacting self-initiated behavioral changes in the context of the recent COVID-19 pandemic [23, 24, 28, 29, 30, 31, 32], including a strong association with a reduction in the number of contacts [33]. We then hypothesize that the adoption and the relaxation of protective behaviors depend not only on objective indicators, given respectively by the burden of the epidemic on the hospitals and by the fraction of vaccinated individuals in the population, but can also be influenced by perceived severity. We can thus investigate within such a framework whether taking into account the population differences in perceived severity leads to differences in the epidemic dynamics and the propagation outcome in terms of various metrics such as the overall death rate and ICU peak height and date. Moreover, we consider several possible methods of informing the model’s parameters by the perceived severity of individuals and explore whether they impact the model’s outcome. We explore several scenarios encompassing distinct behavioral and epidemiological conditions to validate the robustness of our findings.

Results reveal that introducing differences in behavioral change parameters based on perceived severity produces differences in the timing of the epidemic curves, with an earlier peak of infections if compared to the scenario where the entirety of the population behaves in the same way. This has a crucial consequence on a model’s predicted outcome as, in a progressively vaccinated population, an early peak of infections leads to a higher number of deaths. We also find that the precise way of modeling how the perceived severity modulates the parameters ruling the adoption and relaxation of behaviors does not strongly impact the model’s outcome.

These results have direct public health implications: on the one hand, they highlight a certain robustness with respect to some unavoidable arbitrariness in modeling choices. On the other hand, they emphasize the need to gather extensive data on the perception of risks and its correlation with behavioral change in various populations and possibly different epidemiological contexts, to build data-informed models taking into account risk perception.

## 2 Materials and Methods

### 2.1 Data

We leverage data collected during the pandemic period, namely through the CoMix survey [7, 33] initiative, to (i) generate age-stratified contact matrices corresponding to different levels of compliance with respect to self-adopted protective measures and (ii) additionally stratify the population according to the perception of individuals of the risk posed by the disease. The CoMix study developed a longitudinal survey approach for the collection of data aiming at a better comprehension of the behaviors of individuals throughout the COVID-19 pandemic. The surveys, administered every two weeks, captured the evolving awareness, attitudes, and behaviors of participants in response to COVID-19, together with comprehensive information on age, gender, occupation, physical contacts, COVID-19 testing, and self-isolation. It was launched initially in March 2020 in Belgium, the Netherlands, and the United Kingdom, and further expanded its reach to an additional 17 European countries.

We focused our study on the case of Italy where, during the winter between 2020 and 2021, the government implemented a tiered regional system of restrictive measures, with progressively stricter tiers – yellow, orange, and red zones [34]. We use the contact patterns measured in the red zones as a proxy for contacts among individuals adopting highly restrictive protective measures, while contact matrices measured among individuals living in areas where the yellow zone restrictions were implemented represent a situation where individuals adopted less restrictive measures and have a higher number of contacts with respect to what happens in the red zones. We first stratify the population into seven age groups (0-4, 5-17, 18-29, 30-39, 40-49, 50-59, and 60+), using data about the population distribution in Italy across the various age groups from the 2019 United Nations World Population Prospects [35]. We then use the data from the CoMix survey for Italy to generate two contact matrices: the one resulting from data collected in the red zone is used in the model to reflect the contacts of individuals who adopt protective behaviors (named in the rest of the paper as “compliant” individuals), while we use the one built from the yellow zone data to describe the contacts of what we call “non-compliant” individuals (see [36] for more detailed information on the specific top-down restrictions imposed by health authorities during the data collection).

In addition to questions related to their contacts (e.g. total number, frequency, location, etc. - see [36] for detailed information on the contact surveys), participants over 18 years old answered questions related to their risk perception. Concerning perceived severity, participants were asked to indicate their agreement with the statement “Coronavirus would be a serious illness for me” using a 5-point Likert scale ranging from strongly disagree to strongly agree. This information allowed us to further stratify the population in each age group into 5 specific perceived severity subgroups based on participants’ responses (denoted by 1 for low perceived severity, i.e. for the “strongly disagree” answer to 5 for the “strongly agree” denoting a high perceived severity) [7, 33]. For individuals under 18, given the absence of direct data on perceived severity, we assumed that their perceptions were predominantly shaped by their parents. Thus, we categorized age groups 0-4 and 5-17 into subgroups based on the perceived severity responses provided by individuals of parental age (20 to 50 years old). This decision is supported by [37], in which an analysis conducted on adolescents between 13 and 20 years old in Italy during the lockdown evidenced a distribution of perceived severity among them similar to the one we obtained aggregating the data of individuals aged 20 to 50.

Having stratified individuals within each of the 7 age groups into 5 perceived severity groups, we obtain an overall population divided into 35 subgroups, for which we build 35 × 35 contact matrices by multiplying each element of the age-stratified contact matrices by the fraction of the population in each perceived severity group. Specifically, the numbers of contacts between an individual in age group *i* and perceived severity group *j* with an individual in age group *i*^*′*^ and perceived severity group *j*^*′*^, indicated with 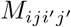,can be obtained with the formula 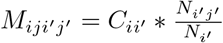, where *C* is the age-stratified contact matrix and *N*_*g*_ indicates the number of individuals in group *g*.

Interestingly, the CoMix survey participants also expressed their perceived susceptibility and risk with the following two statements: “I am likely to catch coronavirus” and “I am worried that I might spread coronavirus to someone who is vulnerable”. However, we did not include these variables in the analysis because their association with the number of contacts, in the context of the COVID-19 emergency, was found to be less relevant in [33].

### 2.2 Model definition

We consider a deterministic compartmental model describing the propagation of SARS-CoV-2 in a population stratified by age and perceived severity of the disease (7 age classes and 5 perceived severity classes), similar to the one used in [17]. The model incorporates on the one hand a vaccination process and on the other hand a behavioral component. The latter describes the possibility of modifying one’s contact patterns depending on the unfolding of the epidemic and it is modulated by a data-informed perceived severity attribute. Figure 1 presents a diagrammatic sketch of the model, which we now describe in detail. A more in-depth explanation of the scenario explored and the sources of the parameters are reported in Section 2.3. We moreover report in the Supplementary Material the full set of evolution equations describing the model.

**Figure 1:**
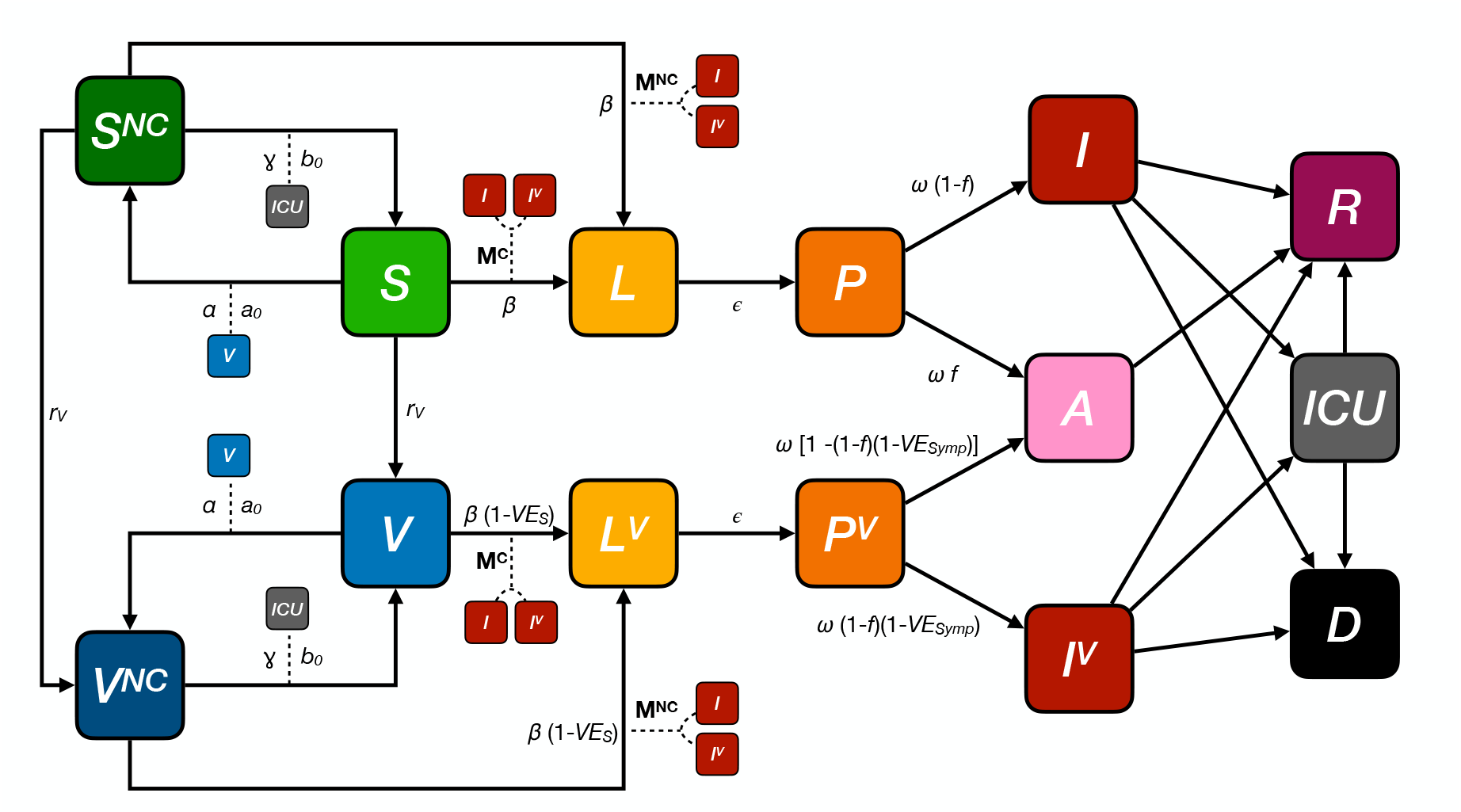
Model diagram. The model is an extension of a standard SLIR (Susceptible-Latent-Infected-Recovered) with the addition of individuals who are pre-symptomatic (*P*) and asymptomatic (*A*) and also individuals in Intensive Care Units (*ICU*), and individuals who die (*D*). Furthermore, we introduced an additional series of compartments for vaccinated individuals (*V, L*^*V*^, *P*^*V*^, *I*^*V*^) and two compartments (*S*^*NC*^ and *V* ^*NC*^) for individuals that relax their protective behaviors. Those individuals have a higher risk of infection, modeled using a contact matrix *M*^*NC*^ with a greater number of contacts than the one used for compliant compartments (*S* and *V*), *M*^*C*^. Both matrices *M*^*C*^ and *M*^*NC*^ capture contact patterns of the winter period 2020-2021 in Italy, for regions with, respectively, high and small stringent restrictions.

#### 2.2.1 Compartmental model

Each individual can transition from one compartment to the other depending on their status with respect to the disease, their vaccination status, and their current behavior.

Susceptible individuals (*S* compartment) in contact with infectious individuals can be infected and transition into the latent (*L* compartment). They then enter the pre-symptomatic (*P*) stage of the infection with a constant rate *ϵ*. They leave the pre-symptomatic stage at a constant rate *ω*, reaching either the asymptomatic compartment *A* with age-dependent probability *f*, or the symptomatic infectious compartment *I* (with probability 1− *f*). Asymptomatic individuals recover at rate *µ*, entering the recovered compartment *R*. Symptomatic infectious individuals can either recover, be hospitalized in the Intensive Care Unit (*ICU* compartment), or die (*D* compartment), with rate *µ* in all cases. The respective probabilities are determined, as described in detail in the Supplementary Material, by the three following age-stratified parameters: the Infection ICU Ratio (*IICUR*) the Infection Fatality Rate (*IFR*), and the Probability of deaths among ICU (*PICUD*). Finally, individuals leave the *ICU* compartment at rate 1*/*Δ, where Δ represents the mean number of days of hospitalization. They then either die with probability *PICUD* or recover with probability 1 − *PICUD*.

The force of infection resulting from contacts between susceptible and symptomatic infectious individuals (compartment *I*) is determined by an age- and perceived severity-stratified “compliant” contact matrix (corresponding to Italy’s red zones) *M*^*C*^, whose elements 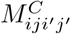 represent the average number of contacts that an individual in age group *i* and perceived severity group *j* has with individuals in age group *i*^*′*^ and perceived severity group *j*^*′*^ per day, and multiplied by a transmission rate *β*. The transmission rate from pre-symptomatic and asymptomatic individuals is reduced by a factor *χ <* 1.

We moreover model a vaccination process as follows: each day, a fraction of the susceptible population receives a vaccine and transitions to the *V* compartment. The rollout rate *r*_*V*_ represents the number of daily available doses expressed as a percentage of the total population. As in previous works [38, 39], we assume the vaccine can reduce susceptibility with efficacy *V E*_*S*_, the probability of developing symptoms with efficacy *V E*_*Symp*_, and severe symptoms leading to death with efficacy *V E*_*D*_. In practice in the model, this means that the infection rate for individuals in the *V* compartment is reduced by a factor (1 − *V E*_*S*_), the probability 1 − *f* of becoming infected *I* instead of asymptomatic *A* is reduced by a factor (1 − *V E*_*Symp*_), and the probability of transitioning from *I*^*V*^ to the *ICU* compartment and the *IFR* are both reduced by a factor (1 − *V E*_*D*_). The overall efficacy of the vaccine is expressed by the following formula *V E* = 1 − (1 − *V E*_*S*_)(1 − *V E*_*Symp*_)(1 − *V E*_*D*_).

#### 2.2.2 Coupling disease dynamics, behavior and vaccination

We assume that individuals, during the epidemic, can change behavior. Specifically, there is a possibility that individuals, both susceptible (*S*) and vaccinated (*V*), may abandon safe behaviors and expose themselves to higher risks of infection. To incorporate this behavioral change, we introduce two additional compartments, *S*^*NC*^ and *V* ^*NC*^, where *NC* stands for non-compliant individuals. We thus assume that these non-compliant individuals have contacts described by Italy’s yellow zone contact matrix *M*^*NC*^ (thus with more contacts than for the *S* and *V* compartments and consequently a higher probability of being infected). For the convenience of notation, we will hereafter denote by *C* the union of the compliant compartments *S* and *V*, and by *NC* the non-compliant compartments *S*^*NC*^ and *V* ^*NC*^.

Our model’s crucial hypothesis concerns the interplay between behavioral change, vaccination, and the unfolding of the disease spread, which determines the transitions between compliant and non-compliant compartments. We first assume that the transition rate from *C* to *NC*, 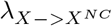 (*X* = *S* or = *V*) describing the relaxation of protective behavior, increases with the fraction of vaccinated people *v*_*t*_ in the population. This expresses the idea that individuals may consider that the threat posed by the disease is lower if more people are vaccinated. Second, we assume on the other hand that the transition rate from *NC* to *C*, 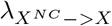 (*X* = *S* or = *V*), depends on the occupancy of beds in intensive care *ICU*_*t*_, taken as a quantitative indicator of the overall seriousness of the spread. Finally, to incorporate all the aforementioned variables, we model both transition rates using logistic functions, described each by two parameters: a slope (*α* for the *C* to *NC* transition and *γ* for the *NC* to *C* one) and a midpoint (*a*_0_ for the *C* to *NC* transition and *b*_0_ for the *NC* to *C* one). The expressions of these rates are as follows (for *X* = *S* and *X* = *V*):

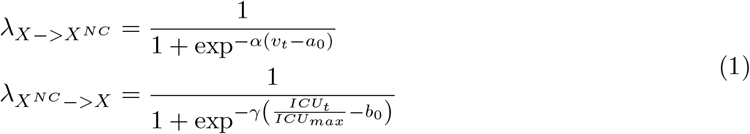

Figure 2 shows the functional dependence of 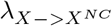 with the fraction of vaccinated individuals and illustrates how the functional shape depends on the slope *α* and the midpoint *a*_0_. Similar plots and considerations stand for the rate 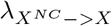 as *γ* and *b*_0_ vary. The way these parameters alter the transition’s shape makes them suitable for variation across different population groups, enabling the introduction of behavioral differences, as we will discuss next.

**Figure 2:**
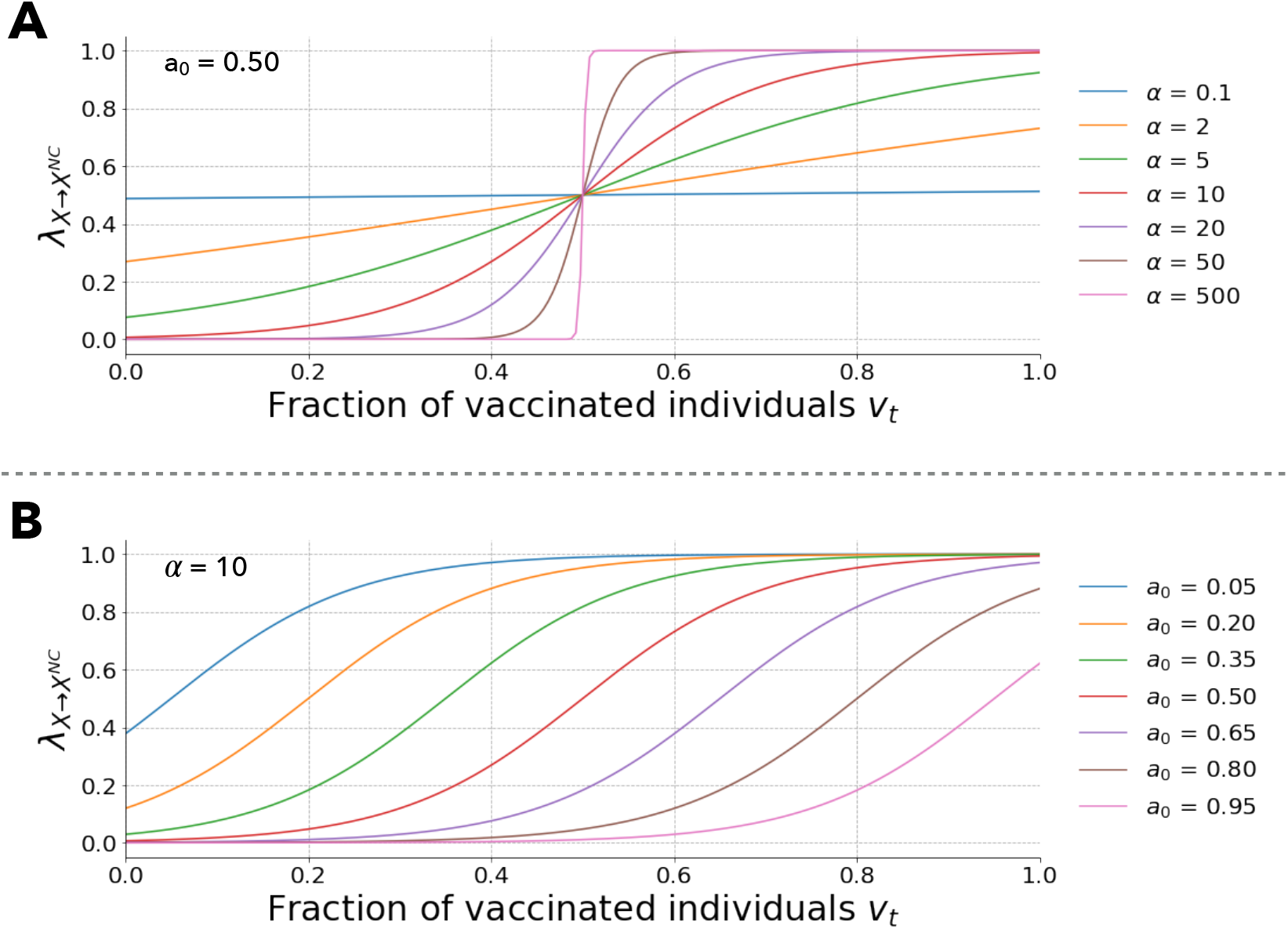
Rate of transition from compliant to non-compliant compartments as a function of the fraction of vaccinated population for several values of A) the slope *α* with a fixed midpoint of *a*_0_ = 0.5, and B) the midpoint *a*_0_ with a fixed slope of *α* = 10.

#### 2.2.3 Modulating behavioral change by risk perception

The final element of our model is the introduction of disease perception as a determinant of behavioral change. In particular, as shown in literature [40, 33, 41], individuals with high perceived severity have a smaller number of contacts, which in our framework corresponds to compliant behavior. For this reason, we assume that the midpoints *a*_0_ and *b*_0_ of the logistic functions of Equation 1, giving the transition rates between compliant and non-compliant compartments, depend on the perceived severity. We instead fix for simplicity the values of the slopes *α* and *γ*.

As individuals with higher perceived severity should be more reluctant either to relax their behavior or more prone to adopt protective measures (with thus fewer contacts), we assume that the midpoint *a*_0_ is an increasing function of perceived severity (= 1, …, 5), while *b*_0_ is instead a decreasing function. How *a*_0_ and *b*_0_ precisely depend on the perceived severity represents however an a priori arbitrary modeling choice. Here we explore possible dynamics for these variables, to explore the different impacts different regimes might have on the overall unfolding of the epidemic. We thus considered five possible different functional forms for the relationship between contacts and perceived severity, sketched in Fig. 3. These five functions all include a linear part with respect to the perceived severity, and we denote them in the remainder of the study by the following names, which describe the location of this linear part: Linear, Center Linear, Start End Linear, Start Linear, and End Linear. Naming 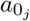 the value of *a*_0_ for the perceived severity group *j* (*j* = 1 to 5), we have:

**Figure 3:**
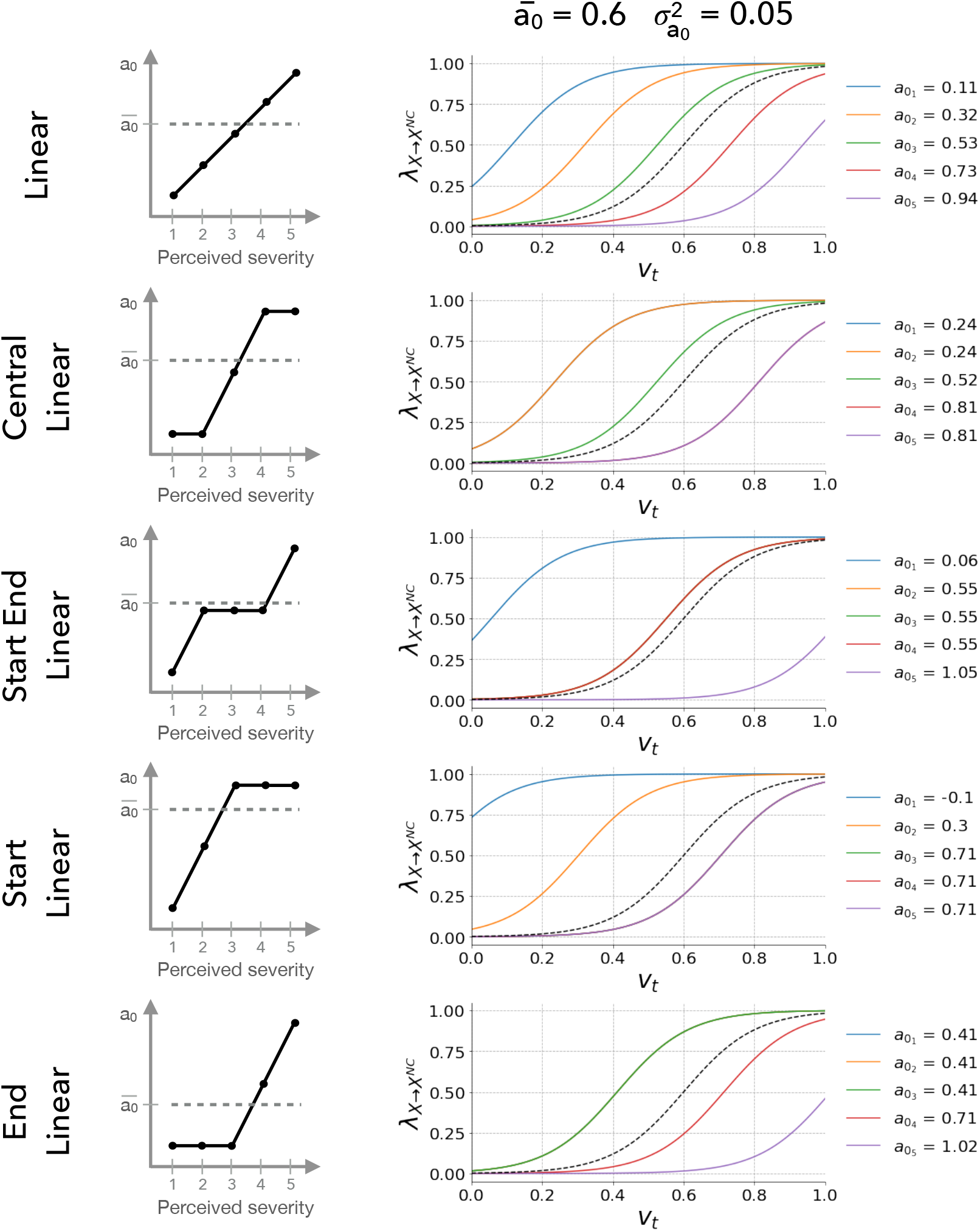
Left column: Sketch of the 5 functions used to model the dependency of the midpoint from perceived severity. From top to bottom, Linear, Central Linear, Start End Linear, Start Linear, End Linear. The horizontal dashed lines give the average 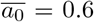. Right column: logistic curves as a function of the fraction of vaccinated individuals, for the various midpoints obtained with a fixed average 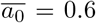 and either variance 0 (dashed line, in which case all midpoints are equal to the average) or variance 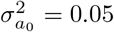.

- Linear: linear growth of the midpoint for all five perceived severity groups 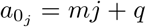
- Central Linear: linear growth for the three central perceived severity groups and the same value of the parameters for the two lowest perceived severity groups and for the two highest 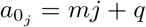 for *i*=2, 3, 4, with 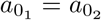 and 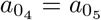.
- Start End Linear: linear growth for the two lowest perceived severity groups and for the two highest and the same value of the parameters for the three central perceived severity groups 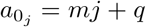 for *i* = 1, 2 and for *i* = 4,5, with 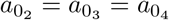.
- Start Linear: linear growth for the three lowest perceived severity groups and the same value of the parameters for the others 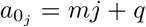 for *i* = 1,2,3, with 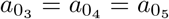.
- End Linear: linear growth for the three highest perceived severity groups and the same value of the parameters for the others 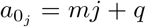 for *i* = 3,4,5, with 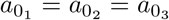.

We show in the Supplementary material how to express the slope *m* and the intercept *q* of each function as a function of the weighted mean 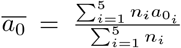 and the weighted variance 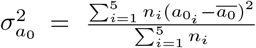 of the midpoints, where *n*_*i*_ is the population in perceived severity group i. Similar calculations apply for the parameter *b*_0_ with the only difference that the midpoint decreases with perceived severity and, thus, the slope *m* of the linear part of each function is negative. In this way, by fixing the mean value of the midpoint (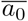or 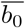) and by changing the variance (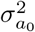 or 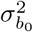), we can model how differences in perceived severity lead to differences in behavioral changes, as determined by the transition rates between compliant and non-compliant compartments. Figure 3 gives a concrete example of this framework. With a fixed mean value of the midpoint 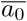, if the variance 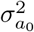 is equal to 0, every perceived severity group has the same midpoint, independently from the function used (dashed line). However, if we increase the variance (i.e. 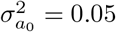 in the figure), different perceived severity groups have different midpoints, leading to different transition rates at a fixed fraction of vaccinated people. Moreover, these transition rates depend on the specific function considered in the model. Note that some values of the midpoints can be negative or larger than 1, resulting in the population in that group being almost entirely non-compliant or compliant for the whole simulation, as their transition rate to the non-compliant compartment is then either always very high or always very small.

Figure 3 focuses on the transition from the compliant to the non-compliant compartments, but similar sketches can be drawn for the dependence of *b*_0_ with perceived severity, albeit with decreasing shapes. We will focus in the main text on the impact of dependence of *a*_0_ on the perceived severity, varying the mean value of the midpoint 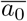 and the variance 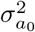, at fixed values of the slopes *α* = 10 and *γ* = 5, and considering the case of 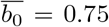 with 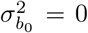 (i.e., no dependence on perceived severity for the transition rate to the compliant compartments). In the Supplementary Material, we perform a sensitivity analysis by varying *α, γ*, and 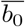, and consider as well the case of *b*_0_ depending on the perceived severity.

### 2.3 Model parameters and scenario

#### Epidemiological parameters

We consider a scenario inspired by the one unfolding in Italy starting in mid-2021, as this corresponded to the deployment of the vaccination campaign against SARS-CoV-2. We therefore use epidemiological parameters matching the characteristics of the Delta variant, which was the dominant one in Italy at that time. In particular, the parameters *ϵ, ω* and *µ* are taken from the literature [42, 43, 44, 45, 46].

The fraction of asymptomatic individuals is age-dependent and taken from [47], where we grouped the asymptomatic and pauci-symptomatic individuals, given that the latter show no clear signs allowing us to identify their disease. Individuals in pre-symptomatic and asymptomatic compartments have lower infectiousness (with respect to the symptomatic ones), quantified by the parameter *χ*, which we take from [48], in line with the value used in other models such as [17] and [49].

We tune *β* to obtain a value of *R*_0_ between 1 and 2.5 in all cases. Indeed, even if the value of *R*_0_ for COVID-19 has reached larger values during the pandemic, in particular during the first wave of 2020 [50, 51, 47], we limited our investigation to such values to take into account the restrictions adopted to mitigate the spread.

We provide in the Supplementary material the detailed computation of the formula yielding the model’s basic reproduction number *R*_0_, which we report here for convenience:

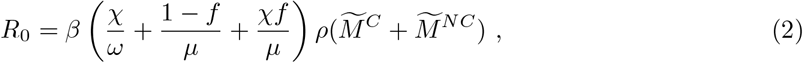

where *ρ* is the spectral radius and 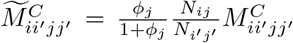, and 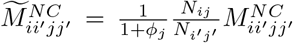 are the two contact matrices weighted by the relative population in different age and perceived severity groups. These matrices also take into account that the initial fraction of compliant individuals at the simulation’s outset, denoted by the term *ϕ*_*j*_, is not necessarily 1.

While *ϵ, ω, µ*, and *χ* are fixed, the fraction of asymptomatic individuals *f* depends on age and we obtain a different value of *R*_0_ for each age group. Moreover, the term *ϕ*_*j*_ depends on the behavioral parameters (*α, γ*, 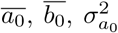, and 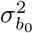), which thus also impact the value of *R*_0_. We refer to the Supplementary Material for more details.

We estimate for the average hospitalization period in ICU Δ a value of 15 days, based on data from the Centers for Disease Control and Prevention [52]. Indeed, for COVID-19 deaths, there was an average interval of approximately two to three weeks between the onset of symptoms and the occurrence of death.

The Infection Fatality Rate (*IFR*) is age-dependent and we used the values reported in [53]. The Infection ICU Ratio (*IICUR*) is obtained from [54] by multiplying the probability of hospitalization if infected and the probability of ICU if hospitalized, which are reported for each age group. On the other hand, for the Probability of Deaths among ICU (*PICUD*), which is also age-stratified, we used data from [55]. Finally, the maximum number of beds in ICU changed notably during the early phase of the pandemic in Italy, going from an initial value of around 5, 000 to over 8, 000 in the Spring of 2021. We chose to use 7, 200, an estimate of the number of beds in intensive care in Italy at the end of 2020 [56].

The parameter values and the corresponding literature sources are reported in Table 1.

**Table 1:** Parameters of the model, with their corresponding values and sources of the literature.

|  | Parameters | Symbols | Value | Source |
| --- | --- | --- | --- | --- |
| **Epidemiological** |  |  |  |  |
| | Transmission rate | $\beta$ | 0.08 | [50, 51, 47] |
| | Inverse of the latent period | $\epsilon$ | 0.25 | [42, 43, 44] |
| | Inverse of the presymptomatic period | $\omega$ | 0.56 | [42] [43, 44] |
| | Inverse of recovery period | $\mu$ | 0.2 | [45, 46] |
| | Mean days of occupancy of ICU bed | $\Delta$ | 15 | [52] |
| | Reduced infectiousness of $P$ and $A$ | $\chi$ | 0.55 | [48] |
| | Fraction of asymptomatic individuals | $f$ | age-stratified | [47] |
| | Reproductive number | $R_0$ | Computed from $\beta$ and $\mu$ | / |
|  | Infection fatality ratio | IFR | age-stratified | [53] |
|  | Infection ICU ratio | IICUR | age-stratified | [54] |
|  | Probability of death among ICU | PICUD | age-stratified | [55] |
| | Available number of ICU beds | $ICU_{max}$ | 7,200 | [56] |
| | Initial number of individuals distributed in the infected compartments | $i_0$ | 550,000 | [57, 58] |
| | Initial number of individuals in ICU compartments | $icu_0$ | 2,500 | [57, 58] |
| | Initial fraction of people in R and D compartments | $r_0, d_0$ | 0.1, 0 | [57, 59, 60] |
| **Vaccination** |  |  |  |  |
| | Rate of vaccination | $r_V$ | 0.025 | [57] |
|  | Vaccination strategy | / | Reverse order of age | [61, 62, 38] |
| | Vaccine efficacy against infection | $VE_S$ | 0.7 | [63, 64, 65] |
| | Vaccine efficacy against symptoms | $VE_{S_{ymp}}$ | 0.5 | [63, 64, 65] |
| | Vaccine efficacy against severe outcomes | $VE_D$ | 0.4 | [63, 64, 65] |
| **Behavioral** |  |  |  |  |
| | Slope of logistic rate $C \rightarrow NC$ | $\alpha$ | 10 | [66] |
| | Slope of logistic rate $NC \rightarrow C$ | $\gamma$ | 5 | [66] |
| | Midpoint of logistic rate $C \rightarrow NC$ | $a_0$ | Variable<br>Perceived severity stratified | / |
| | Midpoint of logistic rate $NC \rightarrow C$ | $b_0$ | Variable<br>Perceived severity stratified | / |

#### Initial conditions

We consider a scenario in which the vaccination campaign starts as the virus has already been able to spread among the population. In the COVID-19 pandemic indeed, the beginning of the spread can be set approximately in February 2020 but the vaccines became available at the end of December 2020. This is also in line with any scenario of a newly emerging virus for which vaccines are not immediately available. We thus do not initialize the model with all individuals in the *S* compartment, but instead, we distribute a fraction of the population in the various infected and recovered compartments, based on observations from various sources [57, 58, 59, 60].

We first distribute 550, 000 individuals, i.e., the estimated number of active cases on the first days of 2021 in Italy [58], in the infectious compartments, namely *L, P, I*, and *A*. The repartition in these compartments is based on the average period of permanence in each compartment (*ϵ*^−1^, *ω*^−1^ and *µ*^−1^, respectively) and on the fraction of asymptomatic individuals *f*. The resulting number of individuals in the *I* compartment is 143, 000, which is a middle ground between the weekly number of confirmed cases in the last weeks of 2020 (approximately 100, 000) and the biweekly one (approximately 200, 000) [57]. We also take into account that the ICU occupancy in Italy at the beginning of 2021 was around 2, 500 individuals [57]. For the initial fraction of recovered individuals we use 0.1, which corresponds to the seroprevalence obtained in independent studies in two different regions of Italy at the end of 2020 [59, 60]. Finally, even if the first wave of 2020 had already caused a considerable number of victims, we set the initial number of people in the death compartment to 0, given that their number is very small compared to the recovered individuals.

#### Vaccination

For the vaccination campaign, we used a vaccination strategy in reverse order of age which is the most effective according to the literature and was the most widely used around the world [61, 62, 38]. We consider a vaccination daily rate of 0.25% of the total population, similar to the rate of vaccination in Italy in Spring 2021 [57]. Finally, consistently with the estimated efficacy of vaccines against COVID-19, we used a vaccine efficacy against infection of *V E*_*S*_ = 70%, and we selected *V E*_*Symp*_ and *V E*_*D*_ to achieve a global vaccine efficacy around 90% [61, 62, 38]. In the Supplementary Material, we also considered scenarios with a lower vaccine efficacy.

#### Behavioral parameters

The behavioral parameters are the ones least constrained by the available data and literature. As explained above, we considered logistic functions to describe the dependency of the transition rates between compliant and non-compliant compartments on the fraction of vaccinated individuals and on the occupancy of ICU beds. We considered five different functions for the dependency of their midpoints *a*_0_ and *b*_0_ on the perceived severity. We fixed the slopes of the logistic functions to *α* = 10 and *γ* = 5. Indeed, we hypothesize that the evolution of behaviors should not have brutal threshold effects: for instance, the relaxation of behaviors during the vaccination campaign has been progressive and not triggered by a particular event [66]. The slopes we consider correspond to logistic functions that are neither too steep nor with too little variation, making them well suited to model the progressive relaxation of behaviors or their re-adoption. In the Supplementary Material, we explored the robustness of our results using other values corresponding to steeper or smoother logistic curves.

#### Numerical integration

We report in the Supplementary Material the full set of equations describing the evolution of the populations in the various compartments. We integrate these equations numerically with a temporal resolution of 1 day, using Python as a programming language and the libraries scipy, numpy, numba, and matplotlib for the visualization part.

## 3 Results

### 3.1 Perceived severity classes and age classes

Figure 4 shows the distribution of the population according to age in each perceived severity group (the distribution of perceived severity in each age group is instead shown in the Supplementary Material). The lowest perceived severity group is the least populated and displays a relatively even distribution across age groups. Groups of individuals with perceived severity 2 and 5 have similar sizes, and groups of perceived severity 3 and 4 are the most populated. As perceived severity increases, the relative contribution of the youngest age groups tends to decrease, and the contribution of the elderly increases. In particular, about 40% of the population of the groups of highest perceived severity (4 and 5) are above 60 years old, and a majority is at least 50. Overall, the majority of the elderly population is in these two groups of high perceived severity (see also Supplementary Material).

**Figure 4:**
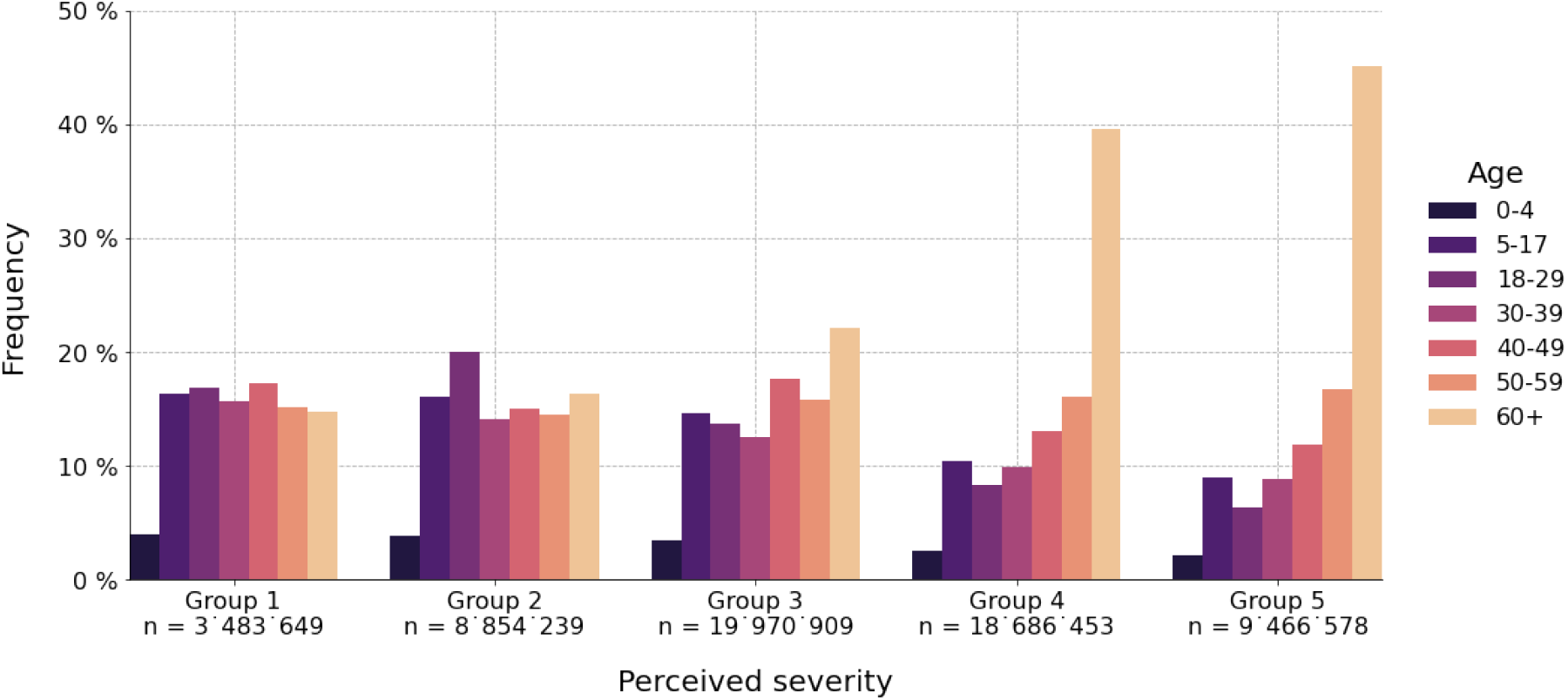
Distribution of age in each perceived severity group. We also report the total number of individuals in each group.

### 3.2 Basic reproduction number

The basic reproduction number *R*_0_ depends on the age groups and on the behavioral parameters (*α, γ*, 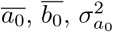, and 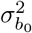), as explained in Section 2.3.

Figure 5 shows the value of *R*_0_ as a function of 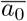 and 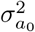, for three of the seven age groups and for the 5 different functions (see the Supplementary Material for the same figure including all age groups). All age groups present similar heatmaps, but younger ones present smaller reproduction numbers at equal 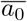 and 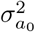 than older groups. This is mainly because the fraction of asymptomatic individuals *f* is much higher in the young population and asymptomatic individuals have a lower infectiousness given by parameter *χ*.

**Figure 5:**
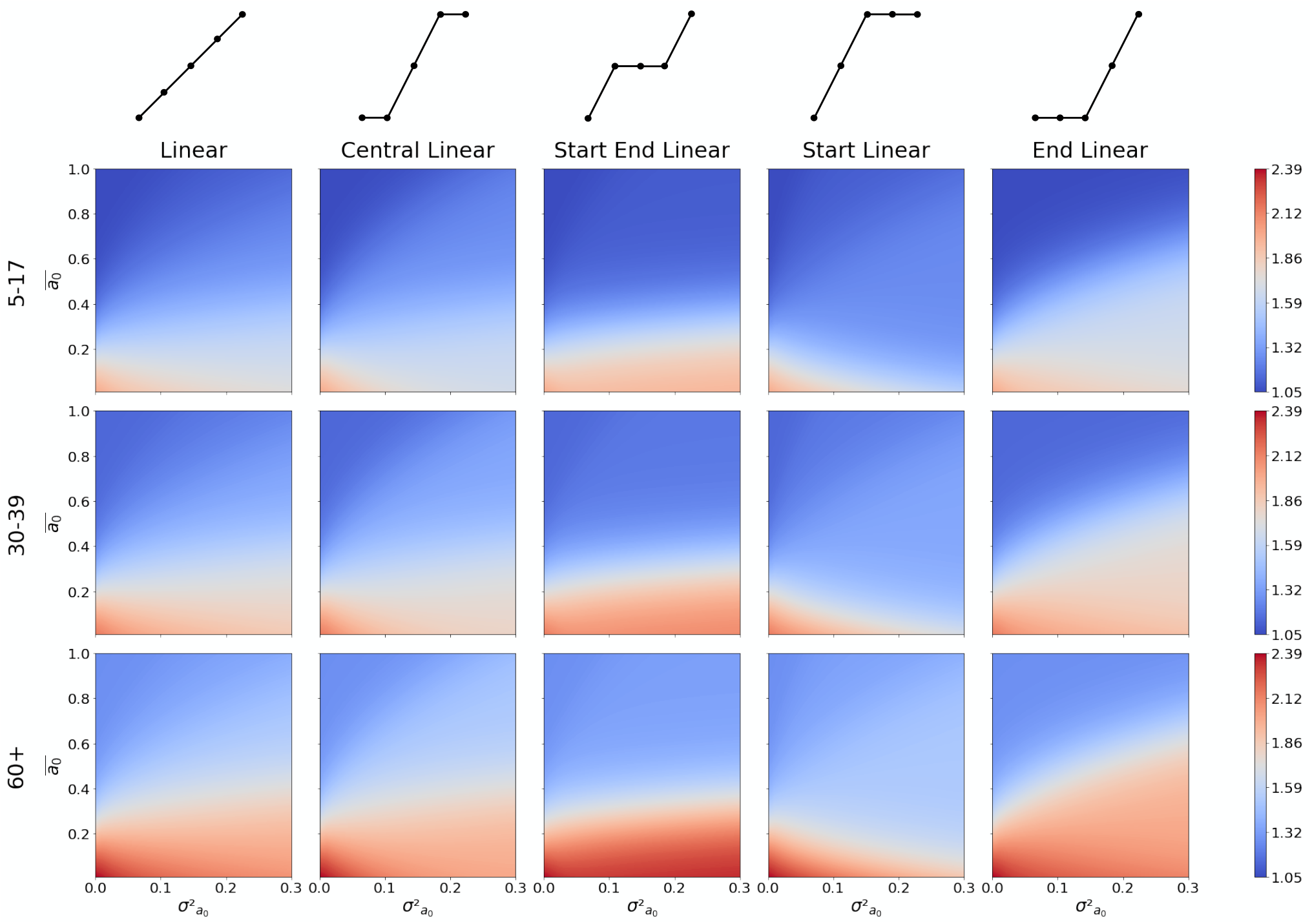
Heatmaps showing the value of *R*_0_ for three age groups (5-17, 30-39, and 60+) as a function of 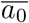 and 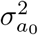. Each heatmap refers to one age group (rows) and one of the 5 functions considered (columns). Above each column is a small diagram of the function, showing how the midpoint *a*_0_ varies from small to high perceived severity groups (left to right). The other parameters used for the simulations are *α* = 10, *γ* = 5, 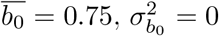. We employed a 900-value grid, with 30 values of 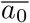 ranging from 0 to 1, and 30 values of 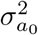 ranging from 0 to 0.3.

We first observe that *R*_0_ decreases systematically as the average midpoint 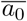 value increases. Indeed, increasing 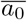 implies that the transition rates from compliant to non-compliant compartments decrease. As a result, even in the initial population, the fraction of non-compliant individuals (who have more contacts) decreases, leading to a reduction in *R*_0_.

Increasing the variance 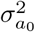 between the midpoints of different perceived severity groups at given 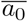 has a more contrasted outcome. It indeed introduces differences in the transition rates 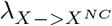 of different perceived severity groups, and thus the fractions of compliant individuals differ across these groups. As a result, groups of individuals with smaller perceived severity exhibit an initial higher fraction of non-compliance than groups with higher perceived severity. For small 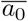, the fact that groups of high perceived severity have a decreasing fraction of non-compliant individuals as 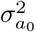 increases leads to a decrease in *R*_0_. For large 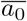 instead, the impact on *R*_0_ comes from the groups of small perceived severity, within which the fraction of non-compliant individuals increases and yields an increase in *R*_0_.

The five functions yield qualitatively similar results, with some quantitative differences due to their shapes and the different distribution of young and elderly individuals in perceived severity groups. For instance, the *R*_0_ for the function Start End Linear almost does not evolve with increased variance. This is because the three groups of intermediate perceived severity (2, 3, 4), who together represent the vast majority of the population, have the same midpoint that has only a small variation with 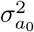. For the Start Linear function instead, *R*_0_ drops substantially when the variance increases for small 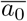, because the groups with the highest perceived severity, which also contain a large fraction of elderly, have an increasing midpoint and thus an increasing fraction of compliant individuals. Conversely, for the function End Linear *R*_0_ also decreases but remains high for small 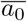 as a large fraction of the population keeps a low value of the midpoint.

### 3.3 Effects of the heterogeneity in perceived severity on the epidemic

We now turn to the role of perceived severity driving different types of behaviors on several metrics describing the outcome of the epidemic spread. Specifically, we measure as outcomes the final total number of deaths, the height of the peak of ICU occupancy (expressed as a fraction of the maximum bed capacity), and the corresponding peak date as 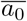 and 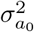 vary (fixing all other parameters and at 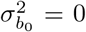). Figure 6 shows the resulting heatmaps for the five functions used to model the dependency between the midpoint and the perceived severity groups.

**Figure 6:**
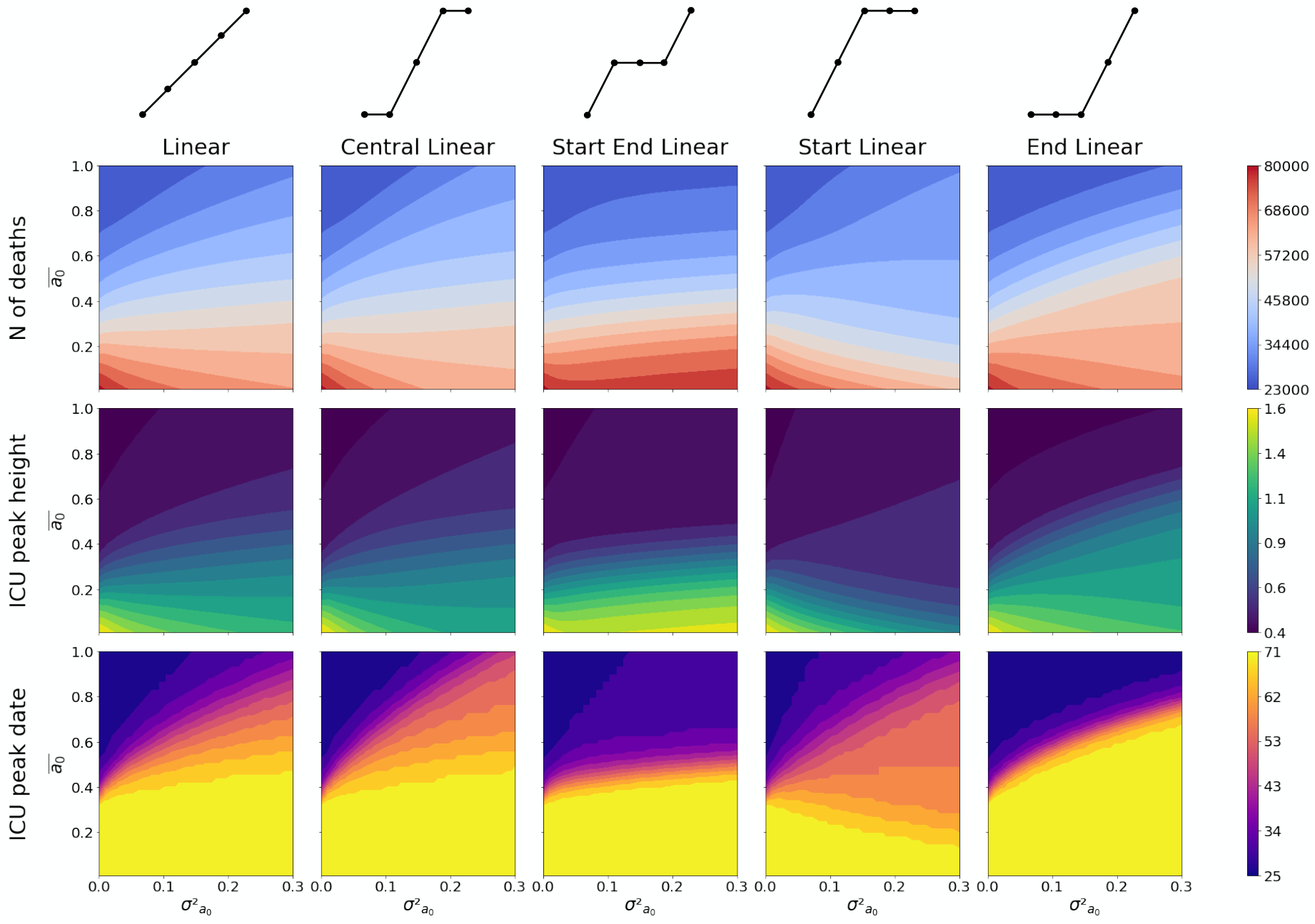
Heatmaps showing the number of deaths (first row), the height of the ICU peak (second row) and its date (third row) as a function of 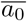 and 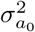. Each column corresponds to one of the five functions linking perceived severity and midpoint of the logistic curve giving the transition rate from compliant to non-compliant compartments as a function of the fraction of vaccinated individuals. Above each column is a small diagram of the function, showing how the midpoint *a*_0_ varies, going from small to high perceived severity groups (left to right). The other parameters used for the simulations are *α* = 10, *γ* = 5, 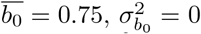. We employed a 900-value grid, with 30 values of 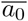 ranging from 0 to 1, and 30 values of 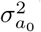 ranging from 0 to 0.3. The rugged profile of the curves related to the peak date is due to its discrete nature, with values representing the (integer) number of days after the simulation’s start in which the peak is observed.

At fixed average 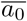, taking into account the fact that groups with different perceived severity have different behavioral parameters by increasing 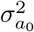 leads to two competing effects. On the one hand, groups of individuals with high perceived severity (comprising a large fraction of the elderly population) have a higher midpoint, and thus a logistic curve describing their rate of transition to non-compliant behavior that is shifted as seen in Fig. 3: as time evolves and the vaccination campaign is rolled out, this transition rate increases only when the population vaccination rate becomes rather high. Therefore, this population relaxes its behavior later with respect to the case of 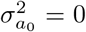, remaining compliant with fewer contacts during a longer period. This would tend to lead to a smaller impact of the epidemic as 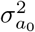 increases. On the other hand, in a symmetric fashion, groups with low perceived severity have a decreasing midpoint as 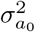 increases, and their transition rate to non-compliance increases thus earlier when the vaccination is rolled out, with respect to the case of 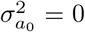 (see Fig. 3). The behavioral relaxation of these groups will then occur earlier, and the resulting increase in contacts will help the spread of the disease. This tends to increase the impact of the spread as 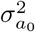increases.

Small mean values of the midpoint 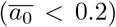 correspond to a population that is on average poorly compliant, as the transition rates to non-compliant behavior increase rapidly when the vaccination progresses. In this case, the first effect is predominant. Indeed, increasing the variance, we observe a reduction in the number of deaths and the height of ICU peak, while the peak date remains stable at around 70 days. The strongest decrease is observed with the Start Linear function, leading to approximately a 25% reduction in deaths and nearly a 40% decrease in the ICU peak. Notably, the ICU peak drops from over 1.6 times the maximum occupancy to below 1, indicating a substantial positive impact of perceived severity. With this functional shape indeed, the three groups of highest perceived severity, comprising a large part of the more vulnerable population (elderly) remain compliant for an increased time (see Fig. 3) with respect to the 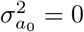 case, becoming thus less infected. The groups of lowest perceived severity become instead only slightly less compliant as their midpoint was already low. Overall, having individuals with higher perceived severity behave in a more conservative way than the ones with low perceived severity brings thus a clear global benefit.

An exception is noted for the Start End Linear function, where the decrease in deaths at low 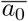 is noted only for small variance, while at larger variance values the situation tends to become worse in terms of the metrics considered. Here indeed, as seen in Fig. 3, the increase in variance leads the vast majority of the population (groups of perceived severity 2, 3, and 4, which also include a substantial number of elderly) to have a slightly lower midpoint than for 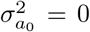 and thus relax their behavior earlier as the vaccine is rolled out, leading to more infections in these groups and thus more deaths.

For high mean values of the midpoint 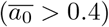, the population is on average largely compliant at the start of the simulation, and the transition rate to the non-compliant behavior increases only when the population is largely vaccinated. The impact of the spread is thus a decreasing function of this average at a given variance. In such cases, increasing 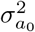 means that the groups with small perceived severity start relaxing their behavior earlier in the vaccination campaign, triggering more contacts and favoring the spread. Overall, the second effect described above prevails: even if groups with high perceived severity keep a compliant behavior longer than for 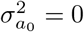, this has little impact.

Figure 6 shows a qualitative similarity of the behavior of the metrics considered for the five functions describing the dependency of the midpoint on the perceived severity. The model’s outcomes are however not strictly identical, and we provide therefore a quantitative evaluation of these differences. To this aim, we compute the Canberra distance between the heatmaps obtained with different functions, for each metric. Specifically, for each pair of functions (*f*_*i*_, *f*_*j*_) and for each metric *K*, the distance is defined by the following formula:

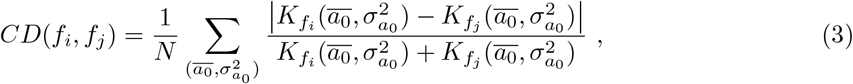

where the sum is over all values of the average 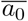 and the variance 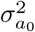 investigated. This distance is bounded between 0 and 1. Table 2 summarizes the results, showing the distances computed for the three metrics. All values are smaller than 0.1, confirming quantitatively the robustness of the model’s phenomenology with respect to the specific way in which the heterogeneities between perceived severity groups influence the relaxation rate of behaviors.

**Table 2:** Canberra distance between each pair of functions for the three metrics considered: number of deaths (a), peak height (b), and peak date (c). We used the grid of values of Figure 6 for 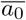 and 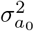. The other parameters are *α* = 10, *γ* = 5, 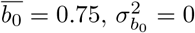.

| (a) Canberra distance – Number of deaths |  |  |  |  |  |
| --- | --- | --- | --- | --- | --- |
|  | Linear | Central Linear | Start End Linear | Start Linear | End Linear |
| Linear |  | 0.0101 | 0.0335 | 0.0379 | 0.0257 |
| Central Linear | 0.0101 |  | 0.0434 | 0.0401 | 0.0332 |
| Start End Linear | 0.0335 | 0.0434 |  | 0.0535 | 0.0399 |
| Start Linear | 0.0379 | 0.0401 | 0.0535 |  | 0.0621 |
| End Linear | 0.0257 | 0.0332 | 0.0399 | 0.0621 |  |

| (b) Canberra distance – ICU peak height |  |  |  |  |  |
| --- | --- | --- | --- | --- | --- |
|  | Linear | Central Linear | Start End Linear | Start Linear | End Linear |
| Linear |  | 0.0114 | 0.0412 | 0.0584 | 0.0367 |
| Central Linear | 0.0114 |  | 0.0525 | 0.0588 | 0.0446 |
| Start End Linear | 0.0412 | 0.0525 |  | 0.0754 | 0.0557 |
| Start Linear | 0.0584 | 0.0588 | 0.0754 |  | 0.0939 |
| End Linear | 0.0367 | 0.0446 | 0.0557 | 0.0939 |  |

Table 2: Canberra distance between each pair of functions for the three metrics considered: number of deaths (a), peak height (b), and peak date (c).
| (c) Canberra distance – Peak date |  |  |  |  |  |
| --- | --- | --- | --- | --- | --- |
|  | Linear | Central Linear | Start End Linear | Start Linear | End Linear |
| Linear |  | 0.0207 | 0.0670 | 0.0330 | 0.0381 |
| Central Linear | 0.0207 |  | 0.0870 | 0.0441 | 0.0544 |
| Start End Linear | 0.0670 | 0.0870 |  | 0.0762 | 0.0609 |
| Start Linear | 0.0330 | 0.0441 | 0.0762 |  | 0.0688 |
| End Linear | 0.0381 | 0.0544 | 0.0609 | 0.0688 |  |

#### Sensitivity analysis

To verify the robustness of the phenomenology described above, we perform some sensitivity analysis with respect to different parameter values (this is shown more in detail in the Supplementary Material). We explore in particular different slopes (*α* and *γ*) of the logistic functions, and a different value of the midpoint 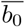. In all cases, a similar picture is obtained as in Fig. 6, with only some small quantitative differences. For instance, a smaller value of 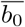 implies that the transition rate from non-compliant to compliant compartments increases more easily when the occupancy of ICU increases: the resulting better adoption of safe behaviors (with fewer contacts) leads thus to a decrease in the metrics describing the impact of the epidemic (number of deaths and ICU peak). Conversely, a higher 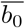 corresponds to a population remaining non-compliant for larger ICU occupancy values, resulting in a larger number of deaths and a higher ICU occupancy peak.

Reducing the slope *α* leads to logistic curves that explore a smaller range of possible values of the transition rate 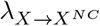 both when the vaccination progresses and when the midpoint *a*_0_ changes (see Fig. 2). As a result, increasing the variance 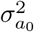 has a small effect on the considered metrics. On the contrary, a larger *α* leads to a more abrupt logistic curve and therefore to a more brutal change of 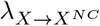 when the fraction of vaccinated individuals reaches the midpoint (see Fig. 2). Differences in the midpoint of different groups have then a stronger impact and the metrics investigated vary more strongly with the variance.

Changing the slope *γ* at fixed 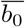 and 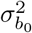 also has a small quantitative effect: small values of *γ* means that the rate of transition 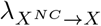 towards compliant behavior is high even for low ICU occupancy, leading thus to an overall more compliant population, a smaller number of deaths and a lower ICU peak. Conversely, a large *γ* with an abrupt logistic curve implies a low 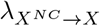 when the ICU occupancy is below *b*_0_, with therefore lower compliance at the beginning of the spread, and finally a stronger impact of the spread.

Finally, we present in the Supplementary Material an analysis of the impact of heterogeneity of the midpoint *b*_0_ of the transition from non-compliant to compliant behavior (i.e., of the impact of having 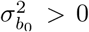). We investigate the same three metrics of the number of deaths, the ICU peak height, and the ICU peak date as a function of 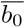 and 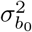, and for 5 possible functions relating perceived severity to the midpoint value. Notably, the results for the five functions are very similar, as shown by the very small values of the Canberra distances between the heatmaps describing the outcomes of the models. In almost every scenario, increasing the variance 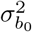 at fixed 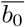 leads to an increase in the number of deaths and a later but higher peak of ICU occupancy. Indeed, the heterogeneities in the transition rate 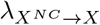 mean that groups with low perceived severity go back to being compliant only for higher ICU occupancy rates, with respect to the case of 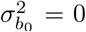, and therefore have more contacts at the beginning of the spread, helping the disease propagate. The fact that groups with high perceived severity, on the opposite, become more compliant, is not enough to compensate for this effect.

### 3.4 Dynamics of the spread

Figure 7 shows the temporal evolution of several important metrics characterizing the unfolding of the spread in the population, for a fixed average midpoint 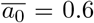, several values of the variances 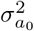, and for a linear functional form linking perceived severity and midpoint *a*_0_: the fraction of vaccinated individuals, of individuals in the *I* compartment, the cumulative fraction of cases (given by the sum of recovered individuals and deaths), of deaths and the ICU occupancy.

**Figure 7:**
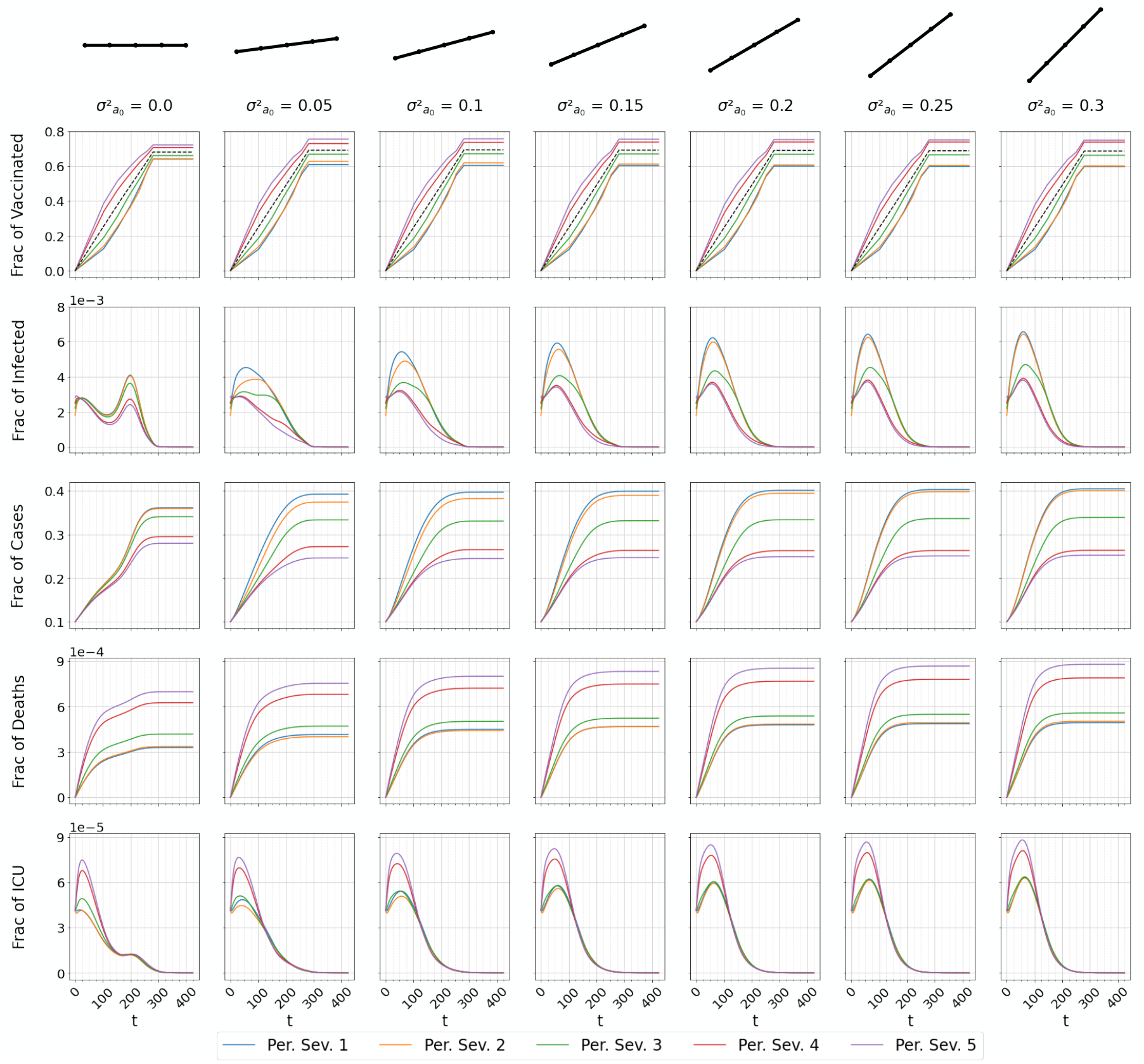
Fraction of vaccinated individuals (first row - the black dashed line reports the global fraction of vaccinated individuals in the population), infected individuals (second row), cases (third row - obtained as the sum of recovered individuals and deaths), deaths (fourth row), and individuals in ICU (fifth row) as a function of time (days), for each perceived severity group. Each column corresponds to a different value of the variance 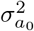, going from 0 to 0.3, with 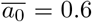. The other parameters are *α* = 10, *γ* = 5, 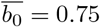, and 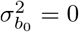.

For 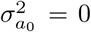 (first column), the whole population has the same rate of relaxation to the noncompliant behavior 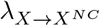. We note that the curves for the different perceived severity curves are however distinct, because of the differences in age distribution in the different groups, and of the differences in numbers of contacts and in epidemiological parameters in the different age groups. In the case considered in Fig. 7, the common midpoint 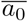 is rather large (60% of the population vaccinated). Thus, the rate of relaxation remains small for a long time and the population keeps a low number of contacts during the whole duration of the first peak of the epidemic curve. As the vaccination campaign progresses however, individuals start to have more contacts, and this triggers a second wave of infections, clearly seen as a second peak in the curves showing the evolution of the fraction of infected individuals. Interestingly however, as a large fraction of the population is then vaccinated, especially among the elderly given the vaccination strategy, this second peak has only a limited impact in terms of ICU occupancy and deaths.

For larger values of the variance, high perceived severity groups have a higher midpoint of the logistic curve; they thus remain compliant longer (until the vaccination reaches a larger fraction of the population). Low perceived severity groups instead have a lower midpoint and tend to relax their behavior sooner. As a result, the first peak of the fraction of infected individuals becomes higher, especially for the groups with low perceived severity (note that the final fraction of vaccinated individuals in groups of perceived severity 1 and 2 slightly decreases, because a larger fraction becomes infected and thus does not need vaccination). The second peak instead disappears. Overall, the early relaxation of behaviors of the groups with low perceived severity has an impact on the whole population: even the epidemic curves for the groups with high perceived severity change shape, with a higher early peak of cases and the disappearance of the second peak. The final total number of cases is larger for the low perceived severity groups (who, having a higher number of contacts, are particularly affected) and smaller for the high perceived severity groups (thanks to the disappearance of the second wave). However, as the early peak is higher and broader even for high perceived severity groups (largely comprised of elderly individuals), it impacts these groups at a time in which vaccine coverage is still limited and causes, therefore, a higher ICU peak and a higher number of deaths. Overall, the early relaxation even for only some groups of individuals leads to a worse outcome for all groups of perceived severity, including the ones who remain more compliant. This result confirms indeed from a dynamical perspective the results of Fig. 6 highlighting a worsening of the final situation at high 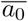 when 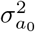increases.

At smaller values of the average midpoint, such as 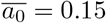, Figure 6 showed that the increase of the variance resulted in a smaller number of deaths and ICU peak heights while maintaining peak timing. We show the impact on the epidemic dynamics of a non-zero variance for 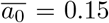 in the Supplementary Material. In scenarios without group heterogeneities 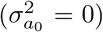, simultaneous behavioral relaxation occurs early, preventing second infection peaks. With higher variance 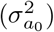, infection peaks for low perceived severity groups grow taller, as their midpoint is negative, rendering them non-compliant from the start. Conversely, higher midpoints for high perceived severity groups lead to smaller infection peaks and reduced ICU occupancy. Consequently, in such scenarios, high heterogeneities in behaviors provide increased protection to high perceived severity groups, composed of a high fraction of elderly individuals, reducing, thus, severe outcomes.

We moreover check (shown in the Supplementary Material) that the shift from an epidemic curve with two peaks to one with a single peak as the variance is increased occurs for all the five functions between perceived severity and midpoint considered in the model, albeit with some differences. In particular, for the Start End Linear function, the epidemic curves for the three groups of intermediate perceived severity (who keep a midpoint close to the average) retain two peaks. The low perceived severity group relaxes its behavior early, and has consequently a higher early peak, while the high perceived severity group, relaxing its behavior only very late, becomes more protected during the second peak. Notably, the evolution of the ICU curve shapes with the variance 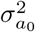 is instead the same for all functions investigated.

We also explore (shown in the Supplementary Material) different values of the slopes *α* and *γ* and of the midpoint 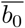. The shift from a two peaks profile to a single peak of the curve showing the fraction of infected individuals vs. time is obtained in almost every scenario analyzed, with the only exception of a very smooth *C* to *NC* transition (low *α*) or small values of 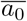.

We moreover investigate the impact of taking into account heterogeneities of perceived severity in the midpoints *b*_0_ of transition rate 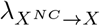, at fixed 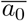 and 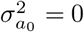. As 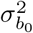 increases, we observe a very similar scenario as the one seen in Fig. 7: the increase in behavior heterogeneity leads to a higher first peak and to the disappearance of the second peak of the epidemic curve, with an overall increased pressure on the ICU occupancy and an increase in the number of deaths.

Finally, we perform a sensitivity analysis with respect to several epidemiological parameters (shown in the Supplementary Material), focusing in particular on vaccine efficacy, initial conditions, average length of the stay in ICU, and maximum number of ICU beds. We also consider a scenario with different latent and pre-symptomatic periods corresponding to the Alpha variant. We obtain similar results to the ones described above.

## 4 Discussion

The disease perception of individuals influences their adoption or disregard of preventive measures, which in turn impacts disease spread. Its significance is particularly pronounced during non-emergency times, such as a post-pandemic period, when the implementation of protective measures is solely reliant on individual choices and there are no top-down restrictions. Our study proposes a novel data-driven modeling framework that integrates disease perception, as measured by the perceived severity, as a key determinant of behavioral change. In particular, we explored a scenario with a competition between a COVID-19 wave and a vaccination campaign, where individuals possess differences in behaviors based on their perceived severity. Individuals with low perceived severity relax behaviors sooner as the vaccination campaign progresses and adopt protective measures only when the epidemiological conditions are more severe (namely, higher occupation of ICU) than individuals with high perceived severity. We leverage CoMix data for Italy [7, 33] to inform this interplay between COVID-19 dynamics, vaccination efforts, and behavioral changes driven by perceived severity: these data make it possible on the one hand to stratify the population both in age groups and in groups of perceived severity, and on the other hand to build contact matrices describing the contacts between different groups, in both situations of compliance and non-compliance to protective measures. Our work marks a twofold addition to the current literature, expanding theoretical frameworks to incorporate data-informed disease perception and investigating how behavioral variations linked to perceived severity affect disease transmission and models’ outcomes.

Results show that behavioral heterogeneities influenced by perceived severity have a substantial impact on the evolution and outcome of the epidemic. These heterogeneities generate two opposing effects. On the one hand, the longer adoption of protective measures by high perceived severity groups (comprising a high proportion of elderly individuals) resulted in higher protection for those individuals. On the other hand, virus spread was facilitated by low perceived severity groups relaxing behaviors more easily. The prevailing effect depended on the overall behavior of the population. In populations that were overall less compliant on average, characterized by high numbers of deaths and ICU peaks, increasing behavioral heterogeneities led to a reduction in these metrics. Conversely, in populations that were on average more compliant, lower severe outcomes were observed, but increasing heterogeneities resulted in an increase in deaths and ICU occupancy. Epidemiological curves gave more insight into this phenomenology. Indeed, when differences in behaviors among groups were not taken into account, we observed a double peak in the evolution of the fraction of infected: the second peak was due to the contemporary relaxation of behaviors by the whole population when the vaccination campaign reached a large enough fraction of the population. Thanks to the high immunization provided by the vaccine, this second peak had small consequences in terms of ICU occupancy and deaths. On the contrary, an increase in the heterogeneities among perceived severity groups caused the disappearance of the second infection peak in favor of a higher first peak for the whole population, resulting in more deaths and ICU hospitalizations, due to the absence of widespread vaccine protection at the time of this first peak. Additionally, our simulations revealed that the specific way we modeled the dependency between behavior relaxation and perceived severity had a small impact on crucial metrics such as the number of deaths and the height of the ICU peak. The sensitivity analysis reported in the Supplementary Material confirmed the robustness of our results. Modifying key epidemiological parameters provided similar pictures, with behavioral heterogeneities consistently impacting metrics and epidemic peaks in the same way across the majority of analyzed scenarios.

Our study comes with several limitations. First of all, we considered a very simple vaccination mechanism. We included in the model a single dose instead of two or more, we assumed a fully working vaccine from the beginning, with no waning phenomenon. Furthermore, despite making extensive use of data and basing our choices on evidence from literature, our model uses several assumptions for which it is not possible to perform a quantitative validation; among those, the mechanism of behavioral change and the expression of the transition rates between compliant and non-compliant compartments based on perceived severity. Additionally, this theoretical framework is intended not for making specific predictions on disease spreading, but rather for conducting comparative analyses of hypothetical scenarios.

Our work has on the other hand some important research and public health implications. Our modeling framework is a data-driven step towards a more comprehensive understanding of how disease perception, particularly perceived severity, can impact the complex dynamics of disease spreading. The strong impact of differences in disease perception on the model’s outcome highlights the importance of taking such heterogeneities into account in models aiming to capture the dynamics of infectious disease transmission and calls for more extensive, continuative, and comprehensive data collections to help uncover which aspects of behavior are most influential. In particular, this study adds insights to the relatively limited empirical research in this area, setting the stage for further exploration and broader understanding necessary to better grasp human adaptive behaviors, especially during non-emergency times. It also paves the way for creating additional data collections drawing on individuals’ personal experiences and perceived risks to help study individual and collective protective strategies. From the perspective of public health, a data-informed identification of the principal factors that drive changes in behavior provides new ways for predicting these shifts and creating more effective communication strategies to reduce transmission among individuals. For instance, our model’s results highlight how the relaxation of behaviors by a limited fraction of the population, who experience a low perceived severity, can negatively impact other groups of the population even if those continue to adopt a self-protective behavior. Communication strategies should thus raise awareness of the global benefits of protective behaviors especially in those groups of the population who are less likely to be affected severely by the disease, to highlight the benefits for at-risk populations.

## Data Availability

All data produced in the present study are available upon reasonable request to the authors

## Acknowledgments

DP and ADG acknowledge support by the VERDI project (101045989), funded by the European Union. Views and opinions expressed in this article are however those of the author(s) only and do not necessarily reflect those of the European Union or the Health and Digital Executive Agency. Neither the European Union nor the granting authority can be held responsible for them. AB acknowledges support from the Agence Nationale de la Recherche (ANR) project DATAREDUX (ANR-19-CE46-0008). All the authors are grateful to Dr Michele Tizzani for support with CoMix data and useful discussions.

## References

[1] N. Ferguson, “Capturing human behaviour,” Nature, vol. 446, no. 7137, pp. 733–733, 2007.

[2] A. Vespignani, “Predicting the behavior of techno-social systems,” Science, vol. 325, no. 5939, pp. 425–428, 2009.

[3] N. Perra, D. Balcan, B. Gonçalves, and A. Vespignani, “Towards a characterization of behavior-disease models,” PloS one, vol. 6, no. 8, p. e23084, 2011.

[4] S. Funk, M. Salathé, and V. A. Jansen, “Modelling the influence of human behaviour on the spread of infectious diseases: a review,” Journal of the Royal Society Interface, vol. 7, no. 50, pp. 1247–1256, 2010.

[5] F. Verelst, L. Willem, and P. Beutels, “Behavioural change models for infectious disease transmission: a systematic review (2010–2015),” Journal of The Royal Society Interface, vol. 13, no. 125, p. 20160820, 2016.

[6] N. Perra, “Non-pharmaceutical interventions during the covid-19 pandemic: A review,” Physics Reports, vol. 913, pp. 1–52, 2021.

[7] F. Verelst, L. Hermans, S. Vercruysse, A. Gimma, P. Coletti, J. A. Backer, K. L. Wong, J. Wambua, K. van Zandvoort, L. Willem, et al., “Socrates-comix: a platform for timely and open-source contact mixing data during and in between covid-19 surges and interventions in over 20 european countries,” BMC medicine, vol. 19, no. 1, pp. 1–7, 2021.

[8] J. A. Salomon, A. Reinhart, A. Bilinski, E. J. Chua, W. La Motte-Kerr, M. M. Rönn, M. B. Reitsma, K. A. Morris, S. LaRocca, T. H. Farag, et al., “The us covid-19 trends and impact survey: Continuous real-time measurement of covid-19 symptoms, risks, protective behaviors, testing, and vaccination,” Proceedings of the National Academy of Sciences, vol. 118, no. 51, p. e2111454118, 2021.

[9] Google LLC, “Google COVID-19 Community Mobility Reports.” https://www.google.com/covid19/mobility/. Accessed: 20/09/23.

[10] T. Hale, N. Angrist, B. Kira, A. Petherick, T. Phillips, and S. Webster, “Variation in government responses to covid-19,” 2020.

[11] E. Del Fava, J. Cimentada, D. Perrotta, A. Grow, F. Rampazzo, S. Gil-Clavel, and E. Zagheni, “Differential impact of physical distancing strategies on social contacts relevant for the spread of sars-cov-2: evidence from a cross-national online survey, march–april 2020,” BMJ open, vol. 11, no. 10, p. e050651, 2021.

[12] U. Basellini, D. Alburez-Gutierrez, E. Del Fava, D. Perrotta, M. Bonetti, C. G. Camarda, and E. Zagheni, “Linking excess mortality to mobility data during the first wave of covid-19 in england and wales,” SSM-Population Health, vol. 14, p. 100799, 2021.

[13] Y. Ge, W.-B. Zhang, H. Liu, C. W. Ruktanonchai, M. Hu, X. Wu, Y. Song, N. W. Ruktanonchai, W. Yan, E. Cleary, et al., “Impacts of worldwide individual non-pharmaceutical interventions on covid-19 transmission across waves and space,” International Journal of Applied Earth Observation and Geoinformation, vol. 106, p. 102649, 2022.

[14] J. Vlachos, E. Hertegård, and H. B. Svaleryd, “The effects of school closures on sars-cov-2 among parents and teachers,” Proceedings of the National Academy of Sciences, vol. 118, no. 9, p. e2020834118, 2021.

[15] T. Hale, N. Angrist, A. J. Hale, B. Kira, S. Majumdar, A. Petherick, T. Phillips, D. Sridhar,R. N. Thompson, S. Webster, et al., “Government responses and covid-19 deaths: Global evidence across multiple pandemic waves,” PLoS One, vol. 16, no. 7, p. e0253116, 2021.

[16] M. Galanti, S. Pei, T. K. Yamana, F. J. Angulo, A. Charos, D. L. Swerdlow, and J. Shaman, “Social distancing remains key during vaccinations,” Science, vol. 371, no. 6528, pp. 473–474, 2021.

[17] N. Gozzi, P. Bajardi, and N. Perra, “The importance of non-pharmaceutical interventions during the covid-19 vaccine rollout,” PLoS computational biology, vol. 17, no. 9, p. e1009346, 2021.

[18] C. N. Ngonghala and A. B. Gumel, “Mathematical assessment of the role of vaccination against covid-19 in the united states,” in Mathematical Modelling, Simulations, and AI for Emergent Pandemic Diseases, pp. 221–249, Elsevier, 2023.

[19] C. N. Ngonghala, H. B. Taboe, S. Safdar, and A. B. Gumel, “Unraveling the dynamics of the omicron and delta variants of the 2019 coronavirus in the presence of vaccination, mask usage, and antiviral treatment,” Applied mathematical modelling, vol. 114, pp. 447–465, 2023.

[20] I. M. Rosenstock, “The health belief model and preventive health behavior,” Health education monographs, vol. 2, no. 4, pp. 354–386, 1974.

[21] G. M. Hochbaum, Public participation in medical screening programs: A socio-psychological study. sNo. 572, US Department of Health, Education, and Welfare, Public Health Service …, 1958.

[22] J. Hayden, Introduction to health behavior theory. Jones & Bartlett Learning, 2022.

[23] H. Seale, A. E. Heywood, J. Leask, M. Sheel, S. Thomas, D. N. Durrheim, K. Bolsewicz, and R. Kaur, “Covid-19 is rapidly changing: Examining public perceptions and behaviors in response to this evolving pandemic,” PloS one, vol. 15, no. 6, p. e0235112, 2020.

[24] W. B. De Bruin and D. Bennett, “Relationships between initial covid-19 risk perceptions and protective health behaviors: a national survey,” American Journal of Preventive Medicine, vol. 59, no. 2, pp. 157–167, 2020.

[25] N. T. Brewer, N. D. Weinstein, C. L. Cuite, and J. E. Herrington, “Risk perceptions and their relation to risk behavior,” Annals of behavioral medicine, vol. 27, pp. 125–130, 2004.

[26] N. D. Weinstein, A. Kwitel, K. D. McCaul, R. E. Magnan, M. Gerrard, and F. X. Gibbons, “Risk perceptions: assessment and relationship to influenza vaccination.,” Health Psychology, vol. 26, no. 2, p. 146, 2007.

[27] R. A. Ferrer and W. M. Klein, “Risk perceptions and health behavior,” Current opinion in psychology, vol. 5, pp. 85–89, 2015.

[28] Z. Vally, “Public perceptions, anxiety and the perceived efficacy of health-protective behaviours to mitigate the spread of the sars-cov-2/covid-19 pandemic,” Public health, vol. 187, pp. 67–73, 2020.

[29] S. Dryhurst, C. R. Schneider, J. Kerr, A. L. Freeman, G. Recchia, A. M. Van Der Bles,D. Spiegelhalter, and S. Van Der Linden, “Risk perceptions of covid-19 around the world,” Journal of risk research, vol. 23, no. 7-8, pp. 994–1006, 2020.

[30] C. R. Schneider, S. Dryhurst, J. Kerr, A. L. Freeman, G. Recchia, D. Spiegelhalter, and S. van der Linden, “Covid-19 risk perception: a longitudinal analysis of its predictors and associations with health protective behaviours in the united kingdom,” Journal of Risk Research, vol. 24, no. 3-4, pp. 294–313, 2021.

[31] M. Siegrist, L. Luchsinger, and A. Bearth, “The impact of trust and risk perception on the acceptance of measures to reduce covid-19 cases,” Risk Analysis, vol. 41, no. 5, pp. 787–800, 2021.

[32] A. Kaim, M. Siman-Tov, E. Jaffe, and B. Adini, “Factors that enhance or impede compliance of the public with governmental regulation of lockdown during covid-19 in israel,” International Journal of Disaster Risk Reduction, vol. 66, p. 102596, 2021.

[33] J. Wambua, N. Loedy, C. I. Jarvis, K. L. M. Wong, C. Faes, R. Grah, B. Prasse, F. Sandmann,R. Niehus, H. Johnson, W. Edmunds, P. Beutels, N. Hens, and P. Coletti, “The influence of covid-19 risk perception and vaccination status on the number of social contacts across europe: insights from the comix study.,” BMC Public Health, vol. 23, p. 1350, 2023.

[34] “Italian ministerial decree (DPCM) of 03/11/2020.” https://www.salute.gov.it/portale/nuovocoronavirus/dettaglioNotizieNuovoCoronavirus.jsp?lingua=english&menu=notizie&p=dalministero&id=5154.

[35] “United Nations, Department of Economic and Social Affairs, Population Division. World Population Prospects: The 2019 Revision; 2020..” https://population.un.org/wpp/Download/Metadata/Documentation/.

[36] M. Tizzani, A. De Gaetano, C. I. Jarvis, A. Gimma, K. Wong, W. J. Edmunds, P. Beutels, N. Hens, P. Coletti, and D. Paolotti, “Impact of tiered measures on social contact and mixing patterns of in italy during the second wave of covid-19,” BMC Public Health, vol. 23, no. 1, p. 906, 2023.

[37] E. Commodari and V. L. La Rosa, “Adolescents in quarantine during covid-19 pandemic in italy: perceived health risk, beliefs, psychological experiences and expectations for the future,” Frontiers in psychology, vol. 11, p. 559951, 2020.

[38] L. Matrajt, J. Eaton, T. Leung, and E. R. Brown, “Vaccine optimization for covid-19: Who to vaccinate first?,” Science Advances, vol. 7, no. 6, p. eabf1374, 2021.

[39] L. Matrajt, J. Eaton, T. Leung, D. Dimitrov, J. T. Schiffer, D. A. Swan, and H. Janes, “Optimizing vaccine allocation for covid-19 vaccines shows the potential role of single-dose vaccination,” Nature communications, vol. 12, no. 1, p. 3449, 2021.

[40] J. Wambua, L. Hermans, P. Coletti, F. Verelst, L. Willem, C. I. Jarvis, A. Gimma, K. L. Wong,A. Lajot, S. Demarest, et al., “The influence of risk perceptions on close contact frequency during the sars-cov-2 pandemic,” Scientific reports, vol. 12, no. 1, p. 5192, 2022.

[41] A. Gimma, J. D. Munday, K. L. Wong, P. Coletti, K. van Zandvoort, K. Prem, C. C.-. working group, P. Klepac, G. J. Rubin, S. Funk, et al., “Changes in social contacts in england duringthe covid-19 pandemic between march 2020 and march 2021 as measured by the comix survey: A repeated cross-sectional study,” PLoS medicine, vol. 19, no. 3, p. e1003907, 2022.

[42] S. A. Lauer, K. H. Grantz, Q. Bi, F. K. Jones, Q. Zheng, H. R. Meredith, A. S. Azman,N. G. Reich, and J. Lessler, “The incubation period of coronavirus disease 2019 (covid-19) from publicly reported confirmed cases: estimation and application,” Annals of internal medicine, vol. 172, no. 9, pp. 577–582, 2020.

[43] L. Ferretti, C. Wymant, M. Kendall, L. Zhao, A. Nurtay, L. Abeler-Dörner, M. Parker, D. Bonsall, and C. Fraser, “Quantifying sars-cov-2 transmission suggests epidemic control with digital contact tracing,” science, vol. 368, no. 6491, p. eabb6936, 2020.

[44] M. Kang, H. Xin, J. Yuan, S. T. Ali, Z. Liang, J. Zhang, T. Hu, E. H. Lau, Y. Zhang, M. Zhang, et al., “Transmission dynamics and epidemiological characteristics of sars-cov-2 delta variant infections in guangdong, china, may to june 2021,” Eurosurveillance, vol. 27, no. 10, p. 2100815, 2022.

[45] “COVID-19 Omicron variant infectious period and transmission from people with asymptomatic compared with symptomatic infection: a rapid review.” https://assets.publishing.service.gov.uk/government/uploads/system/uploads/attachment_data/file/1145484/COVID-19-infectiousness-_asymptomatic-transmission.pdf.

[46] S. M. Kissler, C. Tedijanto, E. Goldstein, Y. H. Grad, and M. Lipsitch, “Projecting the transmission dynamics of sars-cov-2 through the postpandemic period,” Science, vol. 368, no. 6493, pp. 860–868, 2020.

[47] F. Riccardo, M. Ajelli, X. D. Andrianou, A. Bella, M. Del Manso, M. Fabiani, S. Bellino,S. Boros, A. M. Urdiales, V. Marziano, et al., “Epidemiological characteristics of covid-19 cases and estimates of the reproductive numbers 1 month into the epidemic, italy, 28 january to 31 march 2020,” Eurosurveillance, vol. 25, no. 49, p. 2000790, 2020.

[48] R. Li, S. Pei, B. Chen, Y. Song, T. Zhang, W. Yang, and J. Shaman, “Substantial undocumented infection facilitates the rapid dissemination of novel coronavirus (sars-cov-2),” Science, vol. 368, no. 6490, pp. 489–493, 2020.

[49] E. Colosi, G. Bassignana, D. A. Contreras, C. Poirier, P.-Y. Böelle, S. Cauchemez, Y. Yazdanpanah, B. Lina, A. Fontanet, A. Barrat, et al., “Screening and vaccination against covid-19 to minimise school closure: a modelling study,” The Lancet Infectious Diseases, vol. 22, no. 7, pp. 977–989, 2022.

[50] Y. Liu, A. A. Gayle, A. Wilder-Smith, and J. Rocklöv, “The reproductive number of covid-19 is higher compared to sars coronavirus,” Journal of travel medicine, 2020.

[51] D. Cereda, M. Manica, M. Tirani, F. Rovida, V. Demicheli, M. Ajelli, P. Poletti, F. Trentini,G. Guzzetta, V. Marziano, et al., “The early phase of the covid-19 epidemic in lombardy, italy,” Epidemics, vol. 37, p. 100528, 2021.

[52] “Centers for Disease Control and Prevention: COVID-19 Pandemic Planning Scenarios.” https://www.cdc.gov/coronavirus/2019-ncov/hcp/planning-scenarios.html#table-1.

[53] R. Verity, L. C. Okell, I. Dorigatti, P. Winskill, C. Whittaker, N. Imai, G. Cuomo-Dannenburg,H. Thompson, P. G. Walker, H. Fu, et al., “Estimates of the severity of coronavirus disease 2019: a model-based analysis,” The Lancet infectious diseases, vol. 20, no. 6, pp. 669–677, 2020.

[54] H. Salje, C. Tran Kiem, N. Lefrancq, N. Courtejoie, P. Bosetti, J. Paireau, A. Andronico, N. Hozé, J. Richet, C.-L. Dubost, et al., “Estimating the burden of sars-cov-2 in france,” Science, vol. 369, no. 6500, pp. 208–211, 2020.

[55] P. D. W. Garcia, T. Fumeaux, P. Guerci, D. M. Heuberger, J. Montomoli, F. Roche-Campo,R. A. Schuepbach, M. P. Hilty, M. A. Farias, A. Margarit, et al., “Prognostic factors associated with mortality risk and disease progression in 639 critically ill patients with covid-19 in europe: Initial report of the international risc-19-icu prospective observational cohort,” E Clinical Medicine, vol. 25, 2020.

[56] M. Giancotti, “Responses of italian public hospitals to covid-19 pandemic: analysis of supply and demand of hospital icu beds,” in Medical Sciences Forum, vol. 4, p. 16, MDPI, 2021.

[57] E. Mathieu, H. Ritchie, L. Rodés-Guirao, C. Appel, C. Giattino, J. Hasell, B. Macdonald,S. Dattani, D. Beltekian, E. Ortiz-Ospina, and M. Roser, “Coronavirus pandemic (covid-19),” Our World in Data, 2020. https://ourworldindata.org/coronavirus.

[58] C. U. Worldometer, “Cases and deaths from COVID-19 virus pandemic for Italy.” https://www.worldometers.info/coronavirus/country/italy.

[59] S. Paduano, P. Galante, N. Berselli, L. Ugolotti, A. Modenese, A. Poggi, M. Malavolti, S. Turchi,I. Marchesi, R. Vivoli, et al., “Seroprevalence survey of anti-sars-cov-2 antibodies in a population of emilia-romagna region, northern italy,” International Journal of Environmental Research and Public Health, vol. 19, no. 13, p. 7882, 2022.

[60] S. Marchi, C. Coppola, P. Piu, L. Benincasa, F. Dapporto, A. Manenti, S. Viviani, E. Montomoli, and C. M. Trombetta, “Sars-cov-2 epidemiological trend before vaccination era: a seroprevalence study in apulia, southern italy, in 2020,” Journal of Public Health, pp. 1–6, 2023.

[61] W. Wang, Q. Wu, J. Yang, K. Dong, X. Chen, X. Bai, X. Chen, Z. Chen, C. Viboud, M. Ajelli, et al., “Global, regional, and national estimates of target population sizes for covid-19 vaccination: descriptive study,” bmj, vol. 371, 2020.

[62] K. M. Bubar, K. Reinholt, S. M. Kissler, M. Lipsitch, S. Cobey, Y. H. Grad, and D. B. Larremore, “Model-informed covid-19 vaccine prioritization strategies by age and serostatus,” Science, vol. 371, no. 6352, pp. 916–921, 2021.

[63] J. L. Bernal, N. Andrews, C. Gower, C. Robertson, J. Stowe, E. Tessier, R. Simmons, S. Cottrell,R. Roberts, M. O’Doherty, et al., “Effectiveness of the pfizer-biontech and oxford-astrazeneca vaccines on covid-19 related symptoms, hospital admissions, and mortality in older adults in england: test negative case-control study,” bmj, vol. 373, 2021.

[64] K. B. Pouwels, E. Pritchard, P. C. Matthews, N. Stoesser, D. W. Eyre, K.-D. Vihta, T. House,J. Hay, J. I. Bell, J. N. Newton, et al., “Effect of delta variant on viral burden and vaccine effectiveness against new sars-cov-2 infections in the uk,” Nature medicine, vol. 27, no. 12, pp. 2127–2135, 2021.

[65] C. Zheng, W. Shao, X. Chen, B. Zhang, G. Wang, and W. Zhang, “Real-world effectiveness of covid-19 vaccines: a literature review and meta-analysis,” International Journal of Infectious Diseases, vol. 114, pp. 252–260, 2022.

[66] A. De Gaetano, P. Bajardi, N. Gozzi, N. Perra, D. Perrotta, and D. Paolotti, “Behavioral changes associated with covid-19 vaccination: Cross-national online survey,” Journal of Medical Internet Research, vol. 25, p. e47563, 2023.

