## Supplementary Material for "Modeling the interplay between disease spread, behaviors, and disease perception with a data-driven approach"

**Bibliography** **50**

### S1 Data and model

#### S1.1 Perceived severity distribution

As described in the main text, we use data from the CoMix surveys [1] to stratify the population considered in the model, according to age and to their perceived severity. The categorization into five perceived severity groups relies on participants' responses to the statement "Coronavirus would be a serious illness for me", rated on a scale from 1 (lowest) to 5 (highest). In the main text, we showed the age distribution in each perceived severity groups. For completeness we present in Figure S1 the perceived severity distribution across different age groups.

As individuals from age groups 0-4 and 5-17 were not surveyed, we assume that children's perceptions align with their parents' perceived severity and behaviors. We thus aggregated responses from participants aged 20 to 50, to infer the distribution of these two age groups. This assumption finds support in the study by [2], revealing a perceived severity distribution among adolescents (13-20 years old) closely resembling our aggregation of participants aged 20 to 50.

We observe in Figure S1 a general trend of increasing perceived severity with age. The 60+ age group comprises a larger proportion in severity groups 4 or 5 compared to other age groups. Adults below 50 years old exhibit a larger proportion in severity group 3 than in other perceived severity groups.

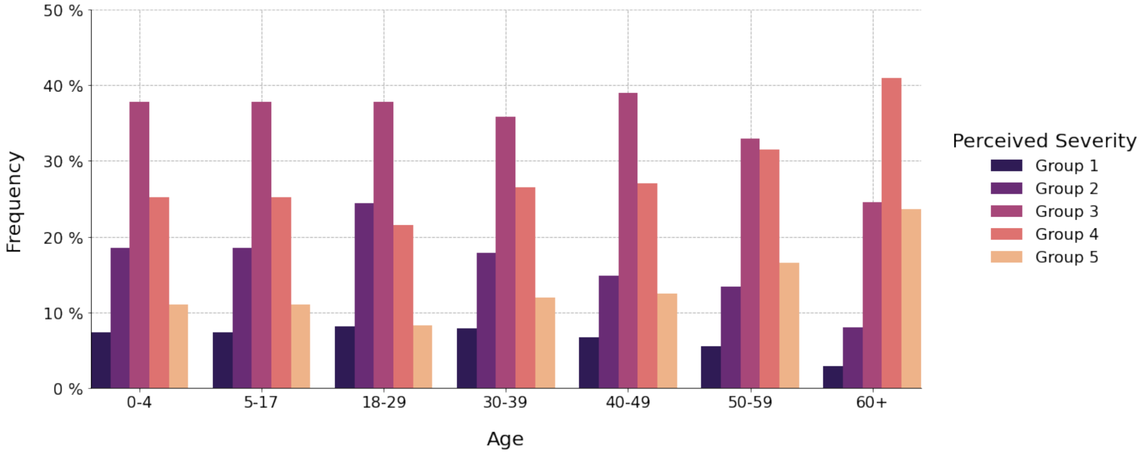

Figure S1: Distribution of perceived severity across the age groups.

#### S1.2 Equations of the model

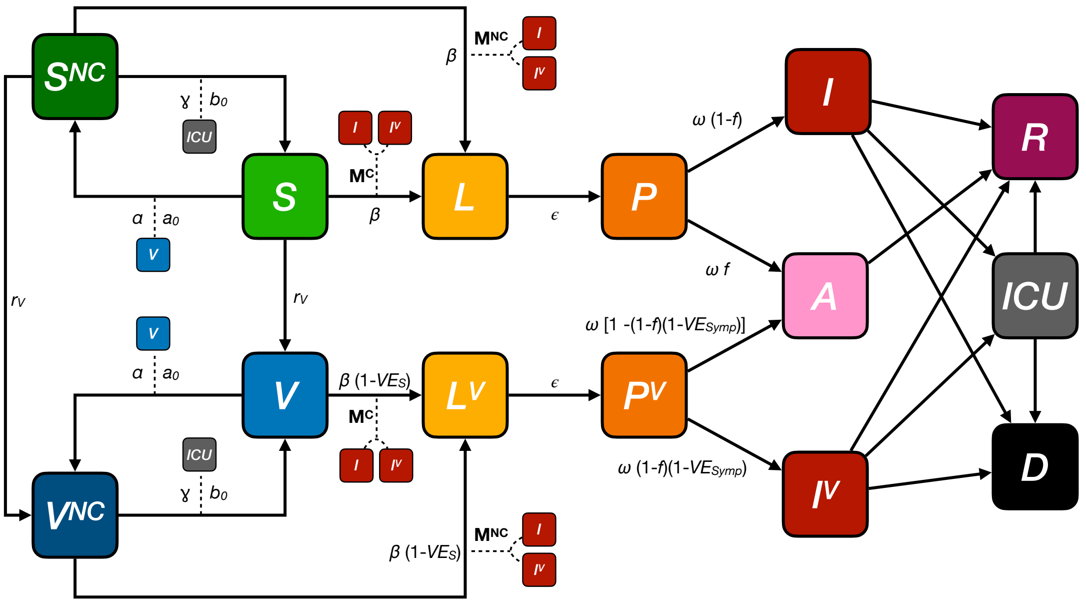

Figure S2: Model diagram. The model is an extension of a standard SLIR (Susceptible-Latent-Infected-Recovered) with the addition of individuals who are pre-symptomatic ($P$) and asymptomatic ($A$) and also individuals in Intensive Care Units ($ICU$), and individuals who die ($D$). Furthermore, we introduced an additional series of compartments for vaccinated individuals ($V$, $L^V$, $P^V$, $I^V$) and two compartments ($S^{NC}$ and $V^{NC}$) for individuals that relax their protective behaviors. Those individuals have a higher risk of infection, modeled using a contact matrix $M^{NC}$ with a greater number of contacts than the one used for compliant compartments ($S$ and $V$), $M^C$. Both matrices $M^C$ and $M^{NC}$ capture contact patterns of the winter period 2020-2021 in Italy, for regions with, respectively, high and small stringent restrictions.

We report for self-consistency the sketch of the model in Figure S2, giving the various model compartments and the possible transitions between these compartments. The evolution of the populations in each compartment, stratified by age group (indicated by the subscript $k$, going from 1 to the number $K$ or age groups) and perceived severity group (subscript $p$, from 1 to the number $P = 5$ of perceived severity groups), are determined by the deterministic equations reported in Equation 1.

$$
\begin{aligned}
\frac{dS_{kp}}{dt} &= -\lambda^{C}_{kp}S_{kp} - \frac{1}{1+\exp\left[-\alpha\left(v_t - a_{0_p}\right)\right]}S_{kp} - r_v S_{kp} + \frac{1}{1+\exp\left[-\gamma\left(\frac{ICU_t}{ICU_{max}} - b_{0_p}\right)\right]}S^{NC}_{kp} \\
\frac{dS^{NC}_{kp}}{dt} &= -\lambda^{NC}_{kp}S^{NC}_{kp} + \frac{1}{1+\exp\left[-\alpha\left(v_t - a_{0_p}\right)\right]}S_{kp} - r_v S^{NC}_{kp} - \frac{1}{1+\exp\left[-\gamma\left(\frac{ICU_t}{ICU_{max}} - b_{0_p}\right)\right]}S^{NC}_{kp} \\
\frac{dV_{kp}}{dt} &= -(1-VE_S)\lambda^{C}_{kp}V_{kp} - \frac{1}{1+\exp\left[-\alpha\left(v_t - a_{0_p}\right)\right]}V_{kp} + r_v S_{kp} + \frac{1}{1+\exp\left[-\gamma\left(\frac{ICU_t}{ICU_{max}} - b_{0_p}\right)\right]}V^{NC}_{kp} \\
\frac{dV^{NC}_{kp}}{dt} &= -(1-VE_S)\lambda^{NC}_{kp}V^{NC}_{kp} + \frac{1}{1+\exp\left[-\alpha\left(v_t - a_{0_p}\right)\right]}V_{kp} + r_v V^{NC}_{kp} - \frac{1}{1+\exp\left[-\gamma\left(\frac{ICU_t}{ICU_{max}} - b_{0_p}\right)\right]}V^{NC}_{kp} \\
\frac{dL_{kp}}{dt} &= \lambda^{C}_{kp}S_{kp} + \lambda^{NC}_{kp}S^{NC}_{kp} - \epsilon L_{kp} \\
\frac{dP_{kp}}{dt} &= \epsilon L_{kp} - \omega P_{kp} \\
\frac{dI_{kp}}{dt} &= \omega(1-f_k)P_{kp} - \mu I_{kp} \\
\frac{dL^{V}_{kp}}{dt} &= +(1-VE_S)\lambda^{C}_{kp}V_{kp} + (1-VE_S)\lambda^{NC}_{kp}V^{NC}_{kp} - \epsilon L^{V}_{kp} \\
\frac{dP^{V}_{kp}}{dt} &= \epsilon L^{V}_{kp} - \omega P^{V}_{kp} \\
\frac{dI^{V}_{kp}}{dt} &= \omega(1-f_k)P^{V}_{kp} - \mu I^{V}_{kp} \\
\frac{dA_{kp}}{dt} &= \omega f P_{kp} + \omega f P^{V}_{kp} - \mu A_{kp} \\
\frac{dR_{kp}}{dt} &= \mu(1 - IICUR_k - \lambda^{deaths}_k)I_{kp} + \frac{1}{\Delta}(1-PICUD_k)ICU_{kp} + \mu[1-(IICUR_k + \lambda^{deaths}_k)(1-VE_D)]I^{V}_{kp} \\
\frac{dICU_{kp}}{dt} &= \mu IICUR_k I_{kp} + \mu IICUR_k(1-VE_D)I^{V}_{kp} - \frac{1}{\Delta}PICUD_k ICU_{kp} + \frac{1}{\Delta}(1-PICUD_k)ICU_{kp} \\
\frac{dD_{kp}}{dt} &= \mu\lambda^{deaths}_k I_{kp} + \mu\lambda^{deaths}_k(1-VE_D)I^{V}_{kp} + \frac{1}{\Delta}PICUD_k ICU_{kp}
\end{aligned} \tag{1}
$$

where

$$
\lambda^{C}_{kp} = \beta\sum_{k'=1}^{K}\sum_{p'=1}^{P} M^{C}_{kk'pp'}\frac{I_{k'p'} + I^{V}_{k'p'} + \chi(P_{kp} + P^{V}_{kp} + A_{kp})}{N_{k'p'}}
$$

$$
\lambda^{NC}_{kp} = \beta\sum_{k'=1}^{K}\sum_{p'=1}^{P} M^{NC}_{kk'pp'}\frac{I_{k'p'} + I^{V}_{k'p'} + \chi(P_{kp} + P^{V}_{kp} + A_{kp})}{N_{k'p'}}
$$

are the force of infection for, respectively, compliant and non compliant individuals in age group $k$ and perceived severity group $p$, and $\mu\lambda^{deaths}_k$ is the rate of infected individuals directly transitioning to the death compartment in age group $k$.

We report in the main text the sources from the literature for the various parameters entering in Eqs 1. Here we detail how we can deduce, from the parameters found in the literature, the transition rates between the infected compartments ($I$, $I^V$) and the compartments of recovered ($R$), ICU ($ICU$) and death ($D$). Indeed, the quantities found in the literature are typically: the Infectious Fatality Rate $IFR$, the Infection ICU Ratio $IICUR$, and the Probability of deaths among ICU $PICUD$.

First, $IICUR$ represents the conditional probability to be recovered in ICU if infected, $P(ICU|I)$. The transition rate from the compartment $I$ to $ICU$ is thus simply $IICUR \times \mu$.

Second, $PICUD$ corresponds to the conditional probability to die if recovered in ICU, $P(D|ICU)$ and the rate from $ICU$ to $D$ is given by the inverse of the mean number of days of occupancy of ICU bed $\frac{1}{\Delta}$ multiplied by this conditional probability. The rate from $ICU$ to $R$ is instead expressed as $\frac{1}{\Delta}$ $P(R|ICU)$. Given that individuals in ICU either recover or die, we have $P(R|ICU)+P(D|ICU) = 1$ and, thus, $P(R|ICU) = 1 - P(D|ICU) = 1 - PICUD$.

We finally need to compute the transition rates for the direct transition from $I$ to $D$ and from $I$ to $R$ (in both cases without going through the $ICU$ compartment). We have access to the Infectious Fatality Rate $IFR$, which represents the probability of dying if infected $P(D|I)$ and can be expanded as follow:

$$IFR = P(D|I) = P(D|ICU)P(ICU|I) + P(D|\neg ICU)P(\neg ICU|I) \ .$$

The first term of the right habd side is $PICUD \times IICUR$ and the second is the probability to die directly when infected, without going through ICU. Calling this probability $\lambda^{deaths} = P(D|I, \neg ICU) = P(D|\neg ICU)P(\neg ICU|I)$, we have thus

$$\lambda^{deaths} = IFR - P(D|ICU)P(ICU|I) = IFR - PICUD \cdot IICUR$$

and the rate of direct transition from $I$ to $D$ is $\mu\lambda^{deaths}$.

For the direct transition from $I$ to $R$ without passing from the ICU compartment, we simply use $P(R|I, \neg ICU) + P(D|I, \neg ICU) + P(ICU|I) = 1$. Using the previous results, we obtain the transition rate for the direct transition from $I$ to $R$ as $\mu \times (1 - IICUR - \lambda^{deaths})$.

The rate of transitions for vaccinated individuals in the $I^V$ compartment are obtained in the same way, with the exception that $IICUR$ and $IFR$ are both reduced by a factor $(1 - VE_D)$ to take into account the efficacy of the vaccine.

#### S1.3 Computation of $R_0$

**Compartments to consider** We compute the value of $R_0$ in the model using the Next Generation Matrix Method. In order to apply it, we need to consider all the equations that regulate the compartments related to the disease, i.e., $L$, $P$, $I$, $A$. However, we leave out the equations and the terms for the vaccinated compartments ($L^V$, $P^V$, $I^V$) because their contribution to the basic reproductive number is of the second order and, thus, become negligible in the Next Generation Matrix Method. Therefore, we are considering a total of $4(K \cdot P)$ equations, where $K$ is the number of age groups and $P$ is the number of perceived severity groups.

$$\begin{aligned}
\frac{dL_{kp}}{dt} &= \beta\lambda^C_{kp}S_{kp} + \beta\lambda^{NC}_{kp}S^{NC}_{kp} - \epsilon L_{kp} \\
\frac{dP_{kp}}{dt} &= \epsilon L_{kp} - \omega P_{kp} \\
\frac{dI_{kp}}{dt} &= \omega(1-f)P_{kp} - \mu I_{kp} \\
\frac{dA_{kp}}{dt} &= \omega f P_{kp} + \omega f P^V_{kp} - \mu A_{kp}
\end{aligned} \tag{2}$$

where $\lambda_{kp}^{C} = \sum_{k'=1}^{K} \sum_{p'=1}^{P} M_{kk'pp'}^{C} \frac{I_{k'p'}+\chi(P_{kp}+A_{kp})}{N_{k'p'}}$ is the force of infection for compliant individuals in age group $k$ and perceived severity group $p$ and $\lambda_{kp}^{NC} = \sum_{k'=1}^{K} \sum_{p'=1}^{P} M_{kk'pp'}^{NC} \frac{I_{k'p'}+\chi(P_{kp}+A_{kp})}{N_{k'p'}}$ is the force of infection for non compliant individuals in age group $k$ and perceived severity group $p$. These equations can be written in matrix notation as:

$$\begin{bmatrix} d_tL_{11} \\ \vdots \\ d_tL_{KP} \\ d_tP_{11} \\ \vdots \\ d_tP_{KP} \\ d_tI_{11} \\ \vdots \\ d_tI_{KP} \\ d_tA_{11} \\ \vdots \\ d_tA_{KP} \end{bmatrix} = \begin{bmatrix} \beta S_{11}\lambda_{11}^{C} + \beta S_{11}^{NC}\lambda_{11}^{NC} \\ \vdots \\ \beta S_{KP}\lambda_{KP}^{C} + \beta S_{KP}^{NC}\lambda_{KP}^{NC} \\ 0 \\ \vdots \\ 0 \\ 0 \\ \vdots \\ 0 \\ 0 \\ \vdots \\ 0 \end{bmatrix} - \begin{bmatrix} \epsilon L_{11} \\ \vdots \\ \epsilon L_{KP} \\ \omega P_{11} - \epsilon L_{11} \\ \vdots \\ \omega P_{KP} - \epsilon L_{KP} \\ \mu I_{11} - \omega(1-f)P_{11} \\ \vdots \\ \mu I_{KP} - \omega(1-f)P_{KP} \\ \mu A_{11} - \omega f P_{11} \\ \vdots \\ \mu A_{KP} - \omega f P_{KP} \end{bmatrix} \tag{3}$$

By denoting the terms of the left hand side of the equation as $d_t\theta_i$, with $i$ going from 1 to $4(K \cdot P)$ and by renaming the two vectors in the right hand side as $T$ and $B$, respectively, we obtain:

$$\begin{bmatrix} d_t\theta_1 \\ \vdots \\ d_t\theta_{KP} \\ d_t\theta_{KP+1} \\ \vdots \\ d_t\theta_{2KP} \\ d_t\theta_{2KP+1} \\ \vdots \\ d_t\theta_{3KP} \\ d_t\theta_{3KP+1} \\ \vdots \\ d_t\theta_{4KP} \end{bmatrix} = \begin{bmatrix} T_1 \\ \vdots \\ T_{KP} \\ 0 \\ \vdots \\ 0 \\ 0 \\ \vdots \\ 0 \\ 0 \\ \vdots \\ 0 \end{bmatrix} - \begin{bmatrix} B_1 \\ \vdots \\ B_{KP} \\ B_{KP+1} \\ \vdots \\ B_{2KP} \\ B_{2KP+1} \\ \vdots \\ B_{3KP} \\ B_{3KP+1} \\ \vdots \\ B_{4KP} \end{bmatrix} . \tag{4}$$

**Disease free equilibrium** In our model, we defined the rate of transition between compliant and non compliant compartments as $\lambda_{X \to X^{NC}} = \frac{1}{1+\exp\left[-\alpha\left(v_t - a_{0_p}\right)\right]}$ with $X = [S, V]$, and the rate between non-compliant and compliant compartment as $\lambda_{X^{NC} \to X} = \frac{1}{1+\exp\left[-\gamma\left(\frac{ICU_t}{ICU_{max}} - b_{0_p}\right)\right]}$ with $X = [S, V]$. At the beginning of the simulations, the fraction of compliant and non-compliant individuals is given by an equilibrium between these two transitions in each age and perceived severity group (for the susceptible population - there are no vaccinated at the initial time):

$$S_{kp}\lambda_{X \to X^{NC}}(0) = S_{kp}^{NC}\lambda_{X^{NC} \to X}(0) \ . \tag{5}$$

At the beginning, we can assume that the number of recovered is much smaller than the total population ($R_k << N_k$), i.e., that all individuals are in susceptible compartments $S_{kp}^{NC} = N_{kp} - S_{kp}$. Therefore, Equation 5 can be rewritten as:

$$S_{kp}\frac{1}{1+\exp\left(\alpha a_{0_p}\right)} = (N_{kp} - S_{kp})\frac{1}{1+\exp\left(\gamma b_{0_p}\right)} \tag{6}$$

and we obtain

$$S_{kp} = (N_{kp} - S_{kp})\frac{1+\exp\left(\alpha a_{0_p}\right)}{1+\exp\left(\gamma b_{0_p}\right)} = (N_{kp} - S_{kp})\phi_p \tag{7}$$

where we called $\phi_p = (1+\exp\left(\alpha a_{0_p}\right))/(1+\exp\left(\gamma b_{0_p}\right))$ the ratio between the two rates at time 0 for perceived severity $p$. We deduce

$$S_{kp} = N_{kp}\frac{\phi_p}{1+\phi_p} \qquad\qquad S_{kp}^{NC} = N_{kp}\frac{1}{1+\phi_p}\ . \tag{8}$$

The disease free equilibrium (DFE) for age group $k$ and perceived severity group $p$ can thus be defined as:

$$(S_{kp}, S_{kp}^{NC}, L_{kp}, P_{kp}, I_{kp}, A_{kp}) = \left(N_{kp}\frac{\phi_p}{1+\phi_p}, N_{kp}\frac{1}{1+\phi_p}, 0, 0, 0, 0\right)\ . \tag{9}$$

**Computing $R_0$** Let us now define the two matrices $F$ and $V$ as follows: $F_{ij} = \frac{dT_i}{d\theta_j}|_{DFE}$ e $V_{ij} = \frac{dB_i}{d\theta_j}|_{DFE}$, where $i, j = 1, \cdots, 4(K \times P)$. These matrices can be written as:

$$F = \begin{bmatrix} 0 & \cdots & 0 & \beta\chi\Psi_{1111} & \cdots & \beta\chi\Psi_{1K1P} & \beta\Psi_{1111} & \cdots & \beta\Psi_{1K1P} & \beta\chi\Psi_{1111} & \cdots & \beta\chi\Psi_{1K1P} \\ \vdots & \ddots & \vdots & \vdots & \ddots & \vdots & \vdots & \ddots & \vdots & \vdots & \ddots & \vdots \\ 0 & \cdots & 0 & \beta\chi\Psi_{K1P1} & \cdots & \beta\chi\Psi_{KKPP} & \beta\Psi_{K1P1} & \cdots & \beta\Psi_{KKPP} & \beta\chi\Psi_{K1P1} & \cdots & \beta\chi\Psi_{KKPP} \\ 0 & \cdots & 0 & 0 & \cdots & 0 & 0 & \cdots & 0 & 0 & \cdots & 0 \\ \vdots & \ddots & \vdots & \vdots & \ddots & \vdots & \vdots & \ddots & \vdots & \vdots & \ddots & \vdots \\ 0 & \cdots & 0 & 0 & \cdots & 0 & 0 & \cdots & 0 & 0 & \cdots & 0 \end{bmatrix} \tag{10}$$

where $\Psi_{ii'jj'} = \frac{N_{ij}[\frac{\phi_j}{1+\phi_j}M_{ii'jj'}^{C} + \frac{1}{1+\phi_j}M_{ii'jj'}^{NC}]}{N_{i'j'}}$

$$V = \begin{bmatrix} \epsilon & \cdots & 0 & 0 & \cdots & 0 & 0 & \cdots & 0 & 0 & \cdots & 0 \\ \vdots & \ddots & \vdots & \vdots & \ddots & \vdots & \vdots & \ddots & \vdots & \vdots & \ddots & \vdots \\ 0 & \cdots & \epsilon & 0 & \cdots & 0 & 0 & \cdots & 0 & 0 & \cdots & 0 \\ -\epsilon & \cdots & 0 & \omega & \cdots & 0 & 0 & \cdots & 0 & 0 & \cdots & 0 \\ \vdots & \ddots & \vdots & \vdots & \ddots & \vdots & \vdots & \ddots & \vdots & \vdots & \ddots & \vdots \\ 0 & \cdots & -\epsilon & 0 & \cdots & \omega & 0 & \cdots & 0 & 0 & \cdots & 0 \\ 0 & \cdots & 0 & -\omega(1-f) & \cdots & 0 & \mu & \cdots & 0 & 0 & \cdots & 0 \\ \vdots & \ddots & \vdots & \vdots & \ddots & \vdots & \vdots & \ddots & \vdots & \vdots & \ddots & \vdots \\ 0 & \cdots & 0 & 0 & \cdots & -\omega(1-f) & 0 & \cdots & \mu & 0 & \cdots & 0 \\ 0 & \cdots & 0 & -\omega f & \cdots & 0 & 0 & \cdots & 0 & \mu & \cdots & 0 \\ \vdots & \ddots & \vdots & \vdots & \ddots & \vdots & \vdots & \ddots & \vdots & \vdots & \ddots & \vdots \\ 0 & \cdots & 0 & 0 & \cdots & -\omega f & 0 & \cdots & 0 & 0 & \cdots & \mu \end{bmatrix} \tag{11}$$

The basic reproduction number is defined as $R_0 = \rho(FV^{-1})$, where $\rho(\cdot)$ indicates the spectral radius. Both $V$ and $F$ can be written in blocks of dimension $(K \cdot P) \times (K \cdot P)$:

$$F = \begin{bmatrix} 0 & \beta\chi(\widetilde{M}^C + \widetilde{M}^{NC}) & \beta(\widetilde{M}^C + \widetilde{M}^{NC}) & \beta\chi(\widetilde{M}^C + \widetilde{M}^{NC}) \\ 0 & 0 & 0 & 0 \\ 0 & 0 & 0 & 0 \\ 0 & 0 & 0 & 0 \end{bmatrix} = \beta(\widetilde{M}^C + \widetilde{M}^{NC}) \begin{bmatrix} 0 & \chi\mathbb{1} & \mathbb{1} & \chi\mathbb{1} \\ 0 & 0 & 0 & 0 \\ 0 & 0 & 0 & 0 \\ 0 & 0 & 0 & 0 \end{bmatrix} \tag{12}$$

$$V = \begin{bmatrix} H & 0 & 0 & 0 \\ J & I & 0 & 0 \\ 0 & L & N & 0 \\ 0 & P & 0 & R \end{bmatrix} \tag{13}$$

All the block components of $V$ are diagonal matrices, and $\widetilde{M}^C$ and $\widetilde{M}^{NC}$ are the contacts matrices weighted by the relative populations of compliant and non-compliant individuals in different age groups and perceived severity groups at the beginning of the simulations (i.e. $\widetilde{M}^C_{ii'jj'} = \frac{\phi_j}{1+\phi_j}\frac{N_{ij}}{N_{i'j'}}M^C_{ii'jj'}$ and $\widetilde{M}^{NC}_{ii'jj'} = \frac{1}{1+\phi_j}\frac{N_{ij}}{N_{i'j'}}M^{NC}_{ii'jj'}$).

Let us first compute $V^{-1}$. The inverse of a block matrix $\begin{bmatrix} X & 0 \\ Y & Z \end{bmatrix}$ can be written as $\begin{bmatrix} X^{-1} & 0 \\ -Z^{-1}YX^{-1} & Z^{-1} \end{bmatrix}$ where, in the case of our $V$ matrix, $X = \begin{bmatrix} H & 0 \\ J & I \end{bmatrix}$, $Y = \begin{bmatrix} 0 & L \\ 0 & P \end{bmatrix}$, and $Z = \begin{bmatrix} N & 0 \\ 0 & R \end{bmatrix}$. We can use the same formula giving the inverse of a 2x2 block matrix to compute the inverse of $X$ and $Z$

$$X^{-1} = \begin{bmatrix} H & 0 \\ J & I \end{bmatrix}^{-1} = \begin{bmatrix} H^{-1} & 0 \\ -I^{-1}JH^{-1} & I^{-1} \end{bmatrix} \tag{14}$$

$$Z^{-1} = \begin{bmatrix} N & 0 \\ 0 & R \end{bmatrix}^{-1} \begin{bmatrix} N^{-1} & 0 \\ 0 & R^{-1} \end{bmatrix}, \tag{15}$$

from which we obtain:

$$\begin{aligned} -Z^{-1}YX^{-1} &= -\begin{bmatrix} N & 0 \\ 0 & R \end{bmatrix}^{-1} \begin{bmatrix} 0 & L \\ 0 & P \end{bmatrix} \begin{bmatrix} H & 0 \\ J & I \end{bmatrix}^{-1} = \\ &= -\begin{bmatrix} N^{-1} & 0 \\ 0 & R^{-1} \end{bmatrix} \begin{bmatrix} 0 & L \\ 0 & P \end{bmatrix} \begin{bmatrix} H^{-1} & 0 \\ -I^{-1}JH^{-1} & I^{-1} \end{bmatrix} = \\ &= -\begin{bmatrix} 0 & N^{-1}L \\ 0 & R^{-1}P \end{bmatrix} \begin{bmatrix} H^{-1} & 0 \\ -I^{-1}JH^{-1} & I^{-1} \end{bmatrix} = \\ &= \begin{bmatrix} N^{-1}LI^{-1}JH^{-1} & -N^{-1}LI^{-1} \\ R^{-1}PI^{-1}JH^{-1} & -R^{-1}PI^{-1} \end{bmatrix} \end{aligned} \tag{16}$$

Substituting all the expressions above in $V^{-1}$ leads to:

$$V^{-1} = \begin{bmatrix} H^{-1} & 0 & 0 & 0 \\ -I^{-1}JH^{-1} & I^{-1} & 0 & 0 \\ N^{-1}LI^{-1}JH^{-1} & -N^{-1}LI^{-1} & N^{-1} & 0 \\ R^{-1}PI^{-1}JH^{-1} & -R^{-1}PI^{-1} & 0 & R^{-1} \end{bmatrix} \tag{17}$$

The next step consists in computing the product $FV^{-1}$:

$$FV^{-1} = \beta(\widetilde{M}^C + \widetilde{M}^{NC}) \begin{bmatrix} 0 & \chi\mathbb{1} & \mathbb{1} & \chi\mathbb{1} \\ 0 & 0 & 0 & 0 \\ 0 & 0 & 0 & 0 \\ 0 & 0 & 0 & 0 \end{bmatrix} \begin{bmatrix} H^{-1} & 0 & 0 & 0 \\ -I^{-1}JH^{-1} & I^{-1} & 0 & 0 \\ N^{-1}LI^{-1}JH^{-1} & -N^{-1}LI^{-1} & N^{-1} & 0 \\ R^{-1}PI^{-1}JH^{-1} & -R^{-1}PI^{-1} & 0 & R^{-1} \end{bmatrix} =$$

$$= \beta(\widetilde{M}^C + \widetilde{M}^{NC}) \begin{bmatrix} (-\chi\mathbb{1} + N^{-1}L + \chi R^{-1}P)I^{-1}JH^{-1} & (\chi\mathbb{1} - +N^{-1}L - \chi R^{-1}P)I^{-1} & N^{-1} & \chi R^{-1} \\ 0 & 0 & 0 & 0 \\ 0 & 0 & 0 & 0 \\ 0 & 0 & 0 & 0 \end{bmatrix} \tag{18}$$

Finally, we are left with finding the spectral radius of $FV^{-1}$ (i.e., its largest eigenvalue). The eigenvalue problem can be written as $\det(FV^{-1}$ - $\lambda\mathbb{1}) = 0$. Given the structure of $FV^{-1}$, and since we are interested in non-trivial solutions ($\lambda \neq 0$), the problem reduces to:

$$det\left[\beta\left(\widetilde{M}^C + \widetilde{M}^{NC}\right)(-\chi\mathbb{1} + N^{-1}L + \chi R^{-1}P)I^{-1}JH^{-1} - \lambda\mathbb{1}\right] = 0 \tag{19}$$

Since $N$, $L$, $R$, $P$, $I$, $J$, and $H$ are all diagonal we can compute the inverses and products and simplify the expression to:

$$det\left[\beta\left(\frac{\chi}{\omega} + \frac{1-f}{\mu} + \frac{\chi f}{\mu}\right)\left(\widetilde{M}^C + \widetilde{M}^{NC}\right) - \lambda\mathbb{1}\right] = 0 \tag{20}$$

Therefore, finding the spectral radius of $FV^{-1}$ is equivalent to solving the eigenvalue problem for $\beta\left(\frac{\chi}{\omega} + \frac{1-f}{\mu} + \frac{\chi f}{\mu}\right)\left(\widetilde{M}^C + \widetilde{M}^{NC}\right)$ and taking the largest eigenvalue. We finally get:

$$R_0 = \beta\left(\frac{\chi}{\omega} + \frac{1-f}{\mu} + \frac{\chi f}{\mu}\right)\rho\left(\widetilde{M}^C + \widetilde{M}^{NC}\right) \; .$$

#### S1.4 Relation between the midpoint values $a_{0_i}$ of the logistic functions of each perceived severity group as a function of the mean $\overline{a_0}$ and variance $\sigma^2_{a_0}$

As explained in the main text, the transition rates between the compliant and non-compliant compartments are given in the model by a logistic function of either the fraction of vaccinated or the ICU relative occupancy. The values of the midpoints of these logistic functions determine whether these rates become rapidly high when the fraction of vaccination increases (for $\lambda_{X\to X^{NC}}$) or when the ICU fill up (for $\lambda_{X^{NC}\to X}$). In the paper, we have explored the impact of a situation in which different perceived severity groups have different midpoints values. In particular, we have considered, at given weighted average of $\overline{a_0}$, the impact of introducing a non-zero weighted variance $\sigma^2_{a_0} > 0$ (mean and variance need to be weighted by the relative population in each perceived severity group, to obtain a global population average and variance).

For $\sigma^2_{a_0} > 0$, groups with higher perceived severity have a higher value for the midpoint $a_0$ of $\lambda_{X\to X^{NC}}$ (hence relaxing their behavior less easily). Symmetrically, for $\sigma^2_{b_0} > 0$ they have a lower value for the midpoint $b_0$ of $\lambda_{X^{NC}\to X}$ (they return to a compliant behaviour more easily). In the main text, we considered five different functions to model the growth of $a_0$ (or the decrease of $b_0$) as the perceived severity increases. Here we illustrate the calculations to obtain the five values of $a_0$ (one for each perceived severity group) at given weighted mean $\overline{a_0}$ and weighted variance $\sigma^2_{a_0}$, for the linear function. Similar calculations can be done to obtain the parameters for the other functions.

We indicate our five perceived severity groups with the index $i$, which goes from 0 (lowest perceived severity group) to 4 (highest perceived severity group), and with $a_{0_i}$ the corresponding parameter that we want to compute. The population of perceived severity group $i$ is $n_i$ and the total population is $N$. If we use a linear function between $i$ and $a_{0_i}$, we have:

$$a_{0_i} = mi + q \tag{21}$$

where $m$ and $q$ are, respectively, the slope and the intercept of the linear function. The average $\overline{a_0}$ can be written as:

$$\overline{a_0} = \frac{\sum_j n_j a_{0_j}}{\sum_j n_j} = \frac{\sum_j n_j (mj+q)}{\sum_j n_j} = \frac{m\sum_j n_j j + q\sum_j n_j}{\sum_j n_j} = \frac{m\sum_j n_j j + qN}{N} \ . \tag{22}$$

From this equation we obtain $q = \overline{a_0} - m\frac{\sum_j n_j j}{N}$ and by substituting it in the formula of $a_{0_i}$ we have:

$$a_{0_i} = \overline{a_0} + m\left(i - \frac{\sum_j n_j j}{N}\right) = \overline{a_0} + m\left(i - \frac{M}{N}\right) \ , \tag{23}$$

where we used $M = \sum_j n_j j$ for simplicity of notation.

To obtain $m$, we now write the expression of $\sigma^2_{a_0}$:

$$\begin{aligned}\sigma^2_{a_0} &= \frac{\sum_j n_j\left(a_{0_j} - \overline{a_0}\right)^2}{\sum_j n_j} = \frac{\sum_j n_j\left[\overline{a_0} + m\left(j - \frac{M}{N}\right) - \overline{a_0}\right]^2}{N} = \frac{m^2\sum_j n_j (j - \frac{M}{N})^2}{N} \\ &= \frac{m^2\sum_j n_j\left(j^2 + \frac{M^2}{N^2} - 2j\frac{M}{N}\right)}{N} = \frac{m^2\left(\sum_j n_j j^2 + \frac{M^2}{N^2}\sum_j n_j - 2\frac{M}{N}\sum_j n_j j\right)}{N} \ .\end{aligned} \tag{24}$$

If we call $D = \sum_j n_j j^2$, we obtain:

$$\sigma^2_{a_0} = \frac{m^2\left(D + \frac{M^2}{N^2}N - 2\frac{M}{N}M\right)}{N} = \frac{m^2\left(D + \frac{M^2}{N} - 2\frac{M^2}{N}\right)}{N} = \frac{m^2\left(D - \frac{M^2}{N}\right)}{N} = \frac{m^2}{N^2}\left(ND - M^2\right) \ . \tag{25}$$

We obtain $m = \sqrt{\frac{N^2\sigma^2}{ND-M^2}}$ ($m$ being positive as $a_{0_i}$ increases with $i$). By substituting it in the formula of $a_{0_i}$ we have:

$$a_{0_i} = \overline{a_0} + \sqrt{\frac{N^2\sigma_{a_0}^2}{ND - M^2}}\left(i - \frac{M}{N}\right) \tag{26}$$

An analogous computation yields for $b_{0_i}$, which decreases with $i$:

$$b_{0_i} = \overline{b_0} - \sqrt{\frac{N^2\sigma_{b_0}^2}{ND - M^2}}\left(i - \frac{M}{N}\right) \tag{27}$$

Similar computations can be done for the other four functions we investigated in the main text.

### S2 $R_0$ for different age groups

$R_0$ depends on several epidemiological parameters, such as $\beta$, $\mu$, $\omega$, $\chi$ and $f$, which are obtained from literature. While the first four parameters are uniform across the population, the fraction $f$ of asymptomatic individuals is age dependent and, thus, $R_0$ will be different for each age group. We also note that $R_0$ depends on the behavioral parameters through its dependency in $\phi_j$.

In the figure 5 of the main text we showed the differences in $R_0$ for three different age groups: 5-17, 30-39, 60+. In Figure S3 we expand these results, showing the value of $R_0$ as a function of $\overline{a_0}$ and $\sigma_{a_0}^2$, for the five different functions (columns) and all the 7 different age groups (rows). The trends observed when increasing the variance are the same for each age group. However, the values of $R_0$ are smaller for younger age groups than for older ones.

In Figure S4 we moreover show the value of $R_0$ for the age group 60+ as a function of $\overline{a_0}$ and $\sigma_{a_0}^2$, for three different slopes $\alpha$ and for the 5 different functions. As discussed in the main text, $R_0$ decreases with increasing variance for small $\overline{a_0}$ and increases for high $\overline{a_0}$. These trends are more pronounced for high values of the slope $\alpha$. In contrast, smoother slopes, such as $\alpha = 4$, lead to a lesser impact of the midpoint value on the logistic curves, so that increasing the variance causes smaller variations in $R_0$ than for the other two values of $\alpha$ illustrated in the figure.

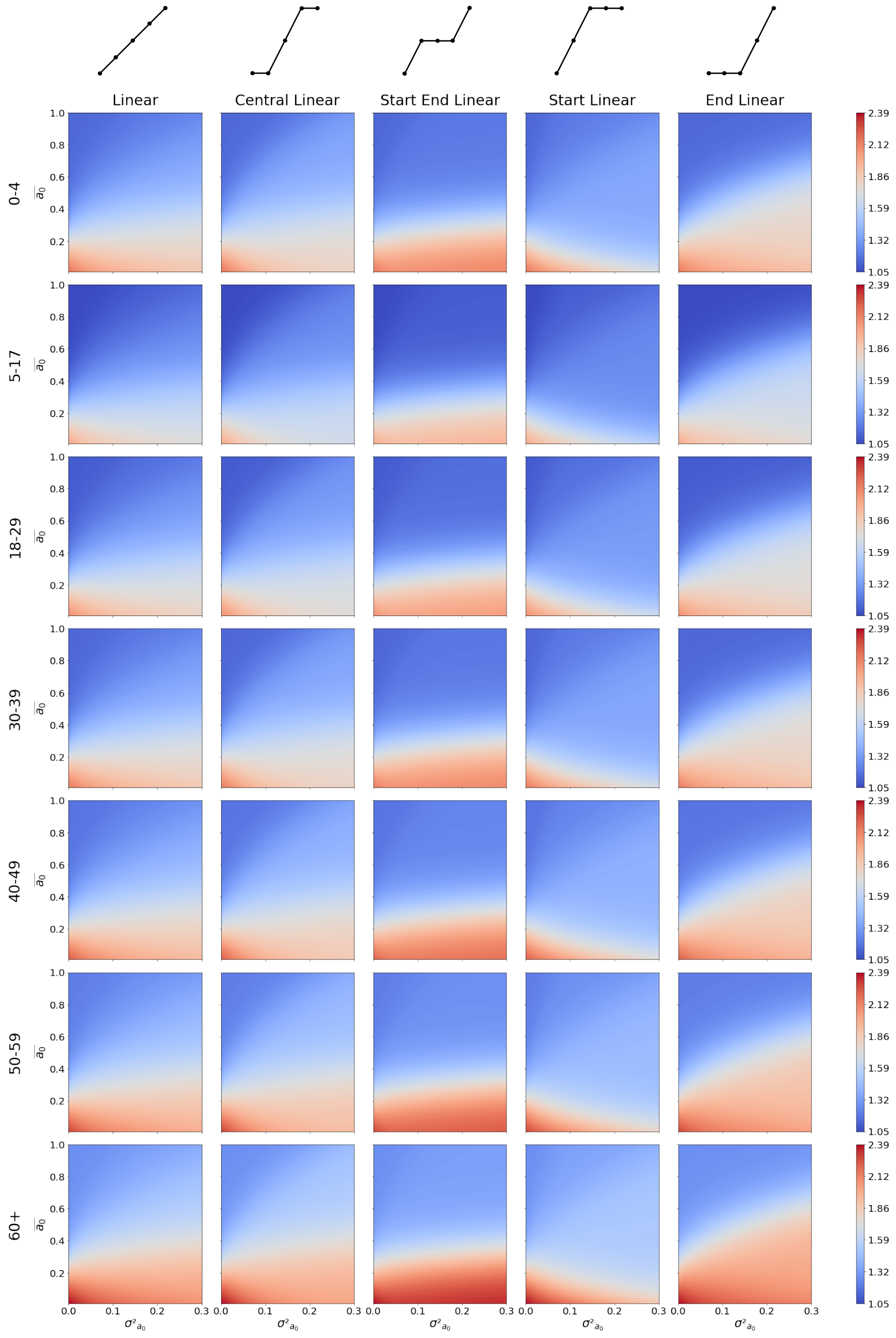

Figure S3: Heatmaps showing the value of $R_0$ as a function of $\overline{a_0}$ and $\sigma^2_{a_0}$. Each heatmap refers to one of the 7 age groups (rows) and one of the 5 functions considered (columns). Above each column is a small diagram of the function, showing how the midpoint $a_0$ varies from small to high perceived severity groups (left to right). The other parameters used for the simulations are $\alpha = 10$, $\gamma = 5$, $\overline{b_0} = 0.75$, $\sigma^2_{b_0} = 0$. We employed a 900-value grid, with 30 values of $\overline{a_0}$ ranging from 0 to 1, and 30 values of $\sigma^2_{a_0}$ ranging from 0 to 0.3.

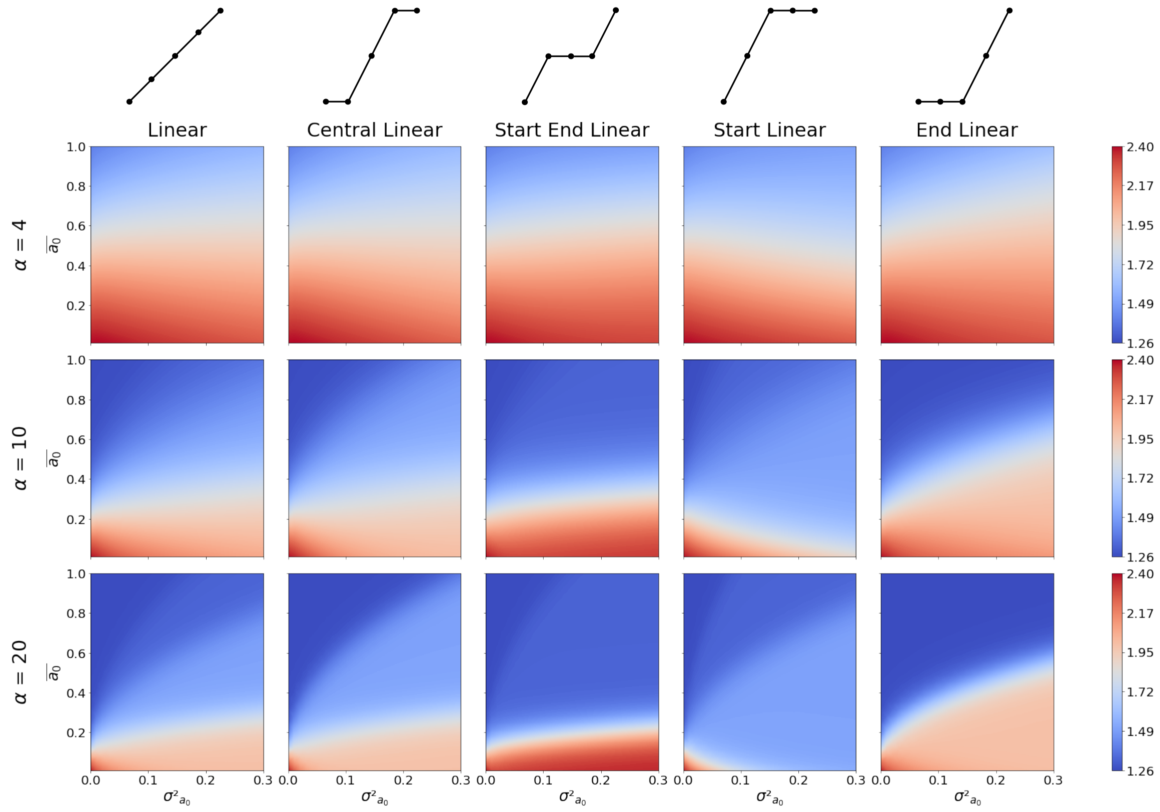

Figure S4: Heatmaps showing the value of $R_0$ for the age group 60+ as a function of $\overline{a_0}$ and $\sigma^2_{a_0}$. Each heatmap refers to one value of the slope $\alpha$ (rows) and one of the 5 functions considered (columns). Above each column is a small diagram of the function, showing how the midpoint $a_0$ varies from small to high perceived severity groups (left to right). The other parameters used for the simulations are $\alpha = 10$, $\gamma = 5$, $\overline{b_0} = 0.75$, $\sigma^2_{b_0} = 0$. We employed a 900-value grid, with 30 values of $\overline{a_0}$ ranging from 0 to 1, and 30 values of $\sigma^2_{a_0}$ ranging from 0 to 0.3.

#### S3 Sensitivity Analysis - Behavioral parameters

In the main article, we presented the three metrics (number of deaths, ICU peak height, and ICU peak date) as a function of the mean value of the midpoints $\overline{a_0}$ and of their variance $\sigma^2_{a_0}$, at fixed values of the other parameters. In particular we have set $\alpha = 10$, $\gamma = 5$ for the slopes of the logistic functions, and $\overline{b_0} = 0.75$ and $\sigma^2_{b_0} = 0$ for the midpoint of the logistic function giving the rate for transition from the non-compliant to the compliant compartments.

Here, we first present the epidemiological curves obtained (i) with the same parameters as in the main text, but for the five different functions linking perceived severity and midpoint $a_0$, and (ii) for a different value of $\overline{a_0}$ than in the main text. We then perform a sensitivity analysis with respect to these choices: we consider different values of $\overline{b_0}$ (still for $\sigma^2_{b_0} = 0$), and of the slopes $\alpha$ and $\gamma$. We finally consider the effect of having the midpoint $b_0$ depending on the perceived severity: on the one hand, at fixed $\overline{a_0}$ and with $\sigma^2_{a_0} = 0$ ($a_0$ independent from the perceived severity), and on the other hand when both $a_0$ and $b_0$ depend on perceived severity (both $\sigma^2_{a_0} > 0$ and $\sigma^2_{b_0} > 0$).

##### S3.1 Epidemiological curves with $\overline{a_0} = 0.6$, for all five functions

In the main article, we showed that the differences between the five functions are small when considering the three metrics analyzed, which are all related to severe disease outcomes (number of deaths or height and date of the ICU peak). We here compare in Figures S5 and S6 the impact of an increase in the variance $\sigma^2_{a_0}$ on the temporal evolution of the fraction of infected and of the ICU occupancy, for the five functions, at fixed $\overline{a_0} = 0.6$. The change in the shape of the epidemic curve as the variance increases, from two peaks to a single one, is overall similar in all cases, with however some distinctions.

The more similar cases in Figure S5 are the ones of the Linear and Central Linear functions. The main difference is that for the Central Linear function, the two highest perceived severity groups share the same midpoint value (the same holds for the two lowest perceived severity groups), so that their epidemic curves are very close.

For the Start Linear function, a slight bump remains at the time of the second peak for high values of $\sigma^2_{a_0}$. This can be attributed to the fact that the three highest perceived severity groups share the same midpoint, slightly larger than $\overline{a_0}$ (see Fig. 3 of the main text). Consequently, the relaxation towards non-compliant behavior occurs for an important fraction of the population in a simultaneous manner towards the end of the vaccination campaign. This has an overall small but still noticeable impact on the epidemic curves.

For the End Linear function. high perceived severity groups have large values of the midpoint, while the three groups with small or medium perceived severity (forming thus a large part of the population) share a midpoint slightly smaller than $\overline{a_0}$. Consequently, a large increase in variance is required to observe the early behavioral relaxation, and the shift from two peaks to a single one occurs at larger variance than for the linear function.

Finally, the Start End Linear function exhibits the most distinct trend, as the second peak never entirely disappears, even for high $\sigma^2_{a_0}$. This is due to the fact that the three groups with perceived severity 2, 3 or 4 share the same midpoint value, which remain close to $\overline{a_0}$. Therefore, the increase in $\sigma^2_{a_0}$ does not lead to an important change for these three groups, and only the groups with highest and lowest perceived severity have a change in their dynamics.

Interestingly, while the different functions used lead thus to some dynamical differences when considering the temporal evolution of the fraction of infected, Figure S6 shows a very strong robustness in how the curves of the ICU occupancy depend on the variance, for the five functions considered.

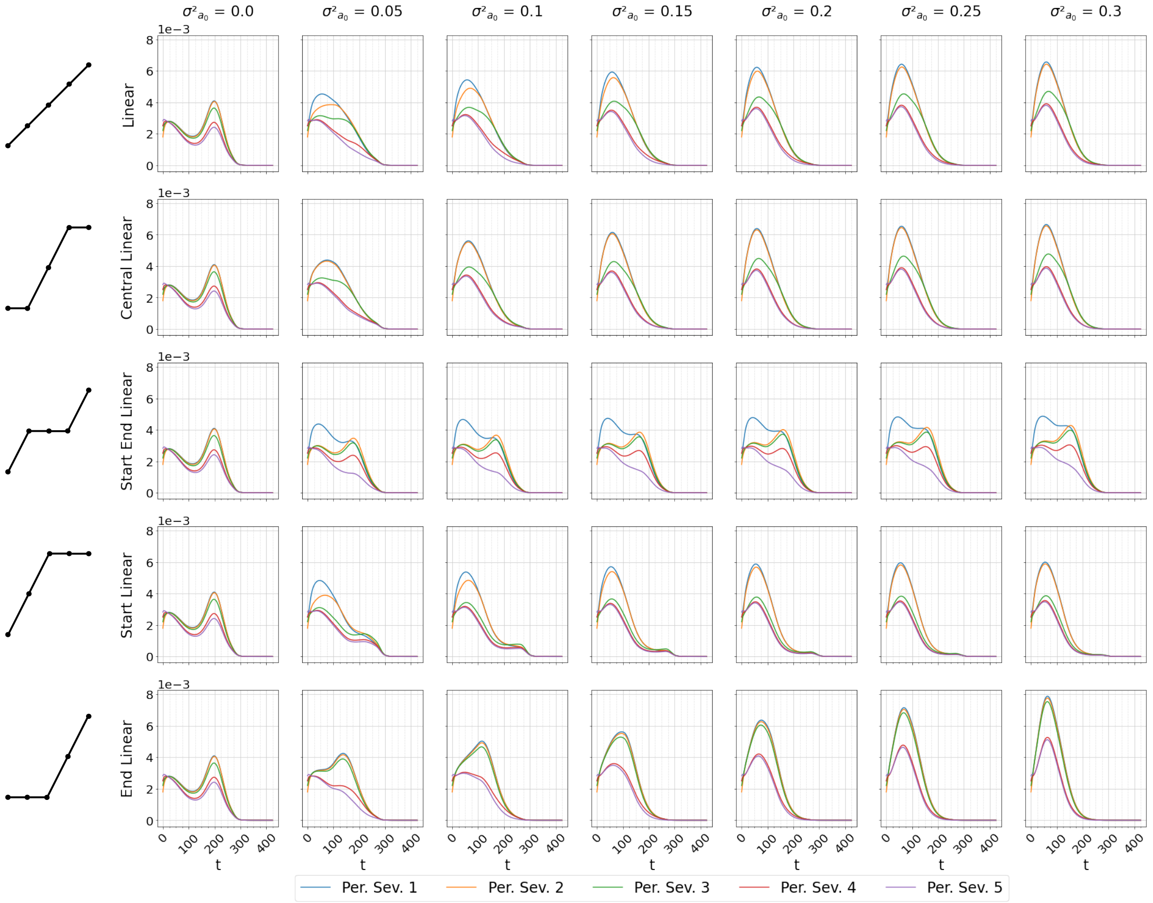

Figure S5: Fraction of infected individuals as a function of time (days) for each perceived severity group. Each row represents one of the 5 functions considered, while each column corresponds to a different value of the variance $\sigma^2_{a_0}$, going from 0 to 0.3. The other parameters used for the simulations are $\alpha = 10$, $\gamma = 5$, $\overline{a_0} = 0.6$, $\overline{b_0} = 0.75$, and $\sigma^2_{b_0} = 0$.

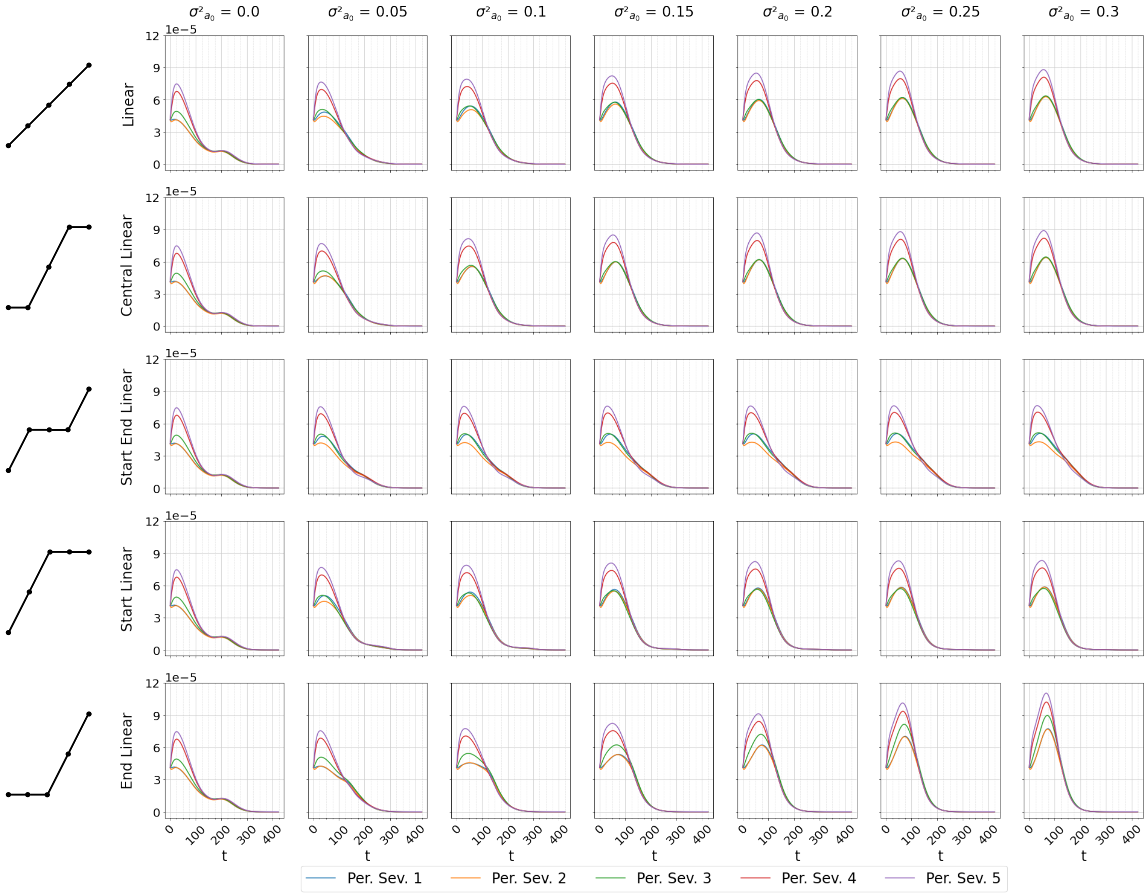

Figure S6: Fraction of individuals in ICU as a function of time (days) for each perceived severity group. Each row represents one of the 5 functions considered, while each column corresponds to a different value of the variance $\sigma^2_{a_0}$, going from 0 to 0.3. The other parameters used for the simulations are $\alpha = 10$, $\gamma = 5$, $\overline{a_0} = 0.6$, $\overline{b_0} = 0.75$, and $\sigma^2_{b_0} = 0$.

#### S3.2 Epidemiological curves with $\overline{a_0} = 0.15$

The epidemiological curves presented in the main article were obtained with a high mean value of the midpoint $\overline{a_0} = 0.6$, using a linear function. In that scenario, increasing the variance $\sigma^2_{a_0}$ revealed a trend where the earlier relaxation of low perceived severity groups facilitated disease spread, outweighing any additional protection afforded to high perceived severity groups due to their prolonged compliance. This led to the disappearance of the second peak in the temporal evolution of the fraction of infected, accompanied by a rise in the height of the initial peak, resulting overall in increased mortality and ICU admissions (as the first peak occurs when the fraction of vaccinated is small).

For a small mean value of the midpoint $\overline{a_0} = 0.15$, Figure 6 of the main text shows that an increase in heterogeneity (in $\sigma^2_{a_0}$) led to a reduction in mortality rates and of the height of the ICU peak. To further explore this point, we show the epidemiological curves with $\overline{a_0} = 0.15$ in Figure S7. When there is no effect of the perceived severity on the transition rates (i.e., $\sigma^2_{a_0} = 0$), the transition rate to non-compliant compartments increase early for all groups (i.e., when a relatively small fraction of the population is vaccinated), leading to early, much higher peaks than for $\overline{a_0} = 0.6$, and more deaths. The second peak observed for $\overline{a_0} = 0.6$ is instead not present (as the population has already relaxed their behavior). If the variance $\sigma^2_{a_0}$ increases, the low perceived severity groups relax their behaviour even earlier (these entire groups become mostly non-compliant already at the start of the simulation), and their infection peak height increases. However, the groups with high perceived severity have then a larger value of the midpoint and thus tend to relax their behaviour later. This leads to a smaller peak for these groups and a reduced ICU occupancy. As these groups are largely composed of elderly individuals, this has a positive impact in terms of reducing the overall number of deaths. Overall, if the population has an average tendency to relax their behaviour early, the heterogeneity between groups leads to a better self-protection of the more vulnerable groups, thereby reducing the global impact in terms of severe outcomes and deaths. In terms of the dynamics of the epidemics, the increase in heterogeneity does not impact the shape of the epidemiological curves but only impacts the heights of the peaks.

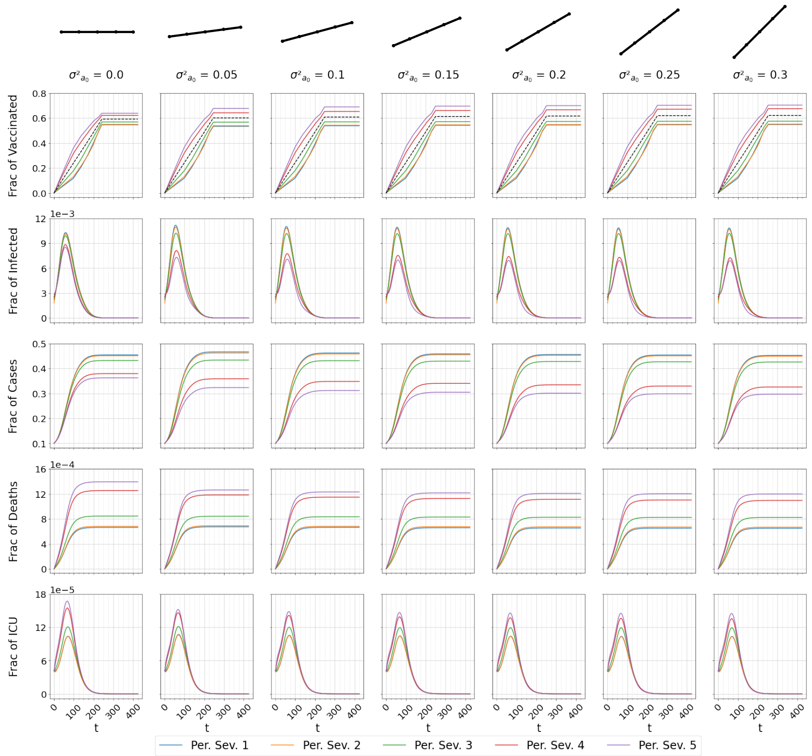

Figure S7: Fraction of vaccinated individuals (first row - the black dashed line reports the global fraction of vaccinated individuals in the population), infected individuals (second row), cases (third row - obtained as the sum of recovered individuals and deaths), deaths (fourth row) and individuals in ICU (fifth row) as a function of time (days), for each perceived severity group. Each column corresponds to a different value of the variance $\sigma^2_{a_0}$, going from 0 to 0.3, with $\overline{a_0} = 0.15$. The other parameters are $\alpha = 10$, $\gamma = 5$, $\overline{b_0} = 0.75$, and $\sigma^2_{b_0} = 0$.

##### S3.3 Changing the average midpoint $\overline{b_0}$, at $\sigma^2_{b_0} = 0$.

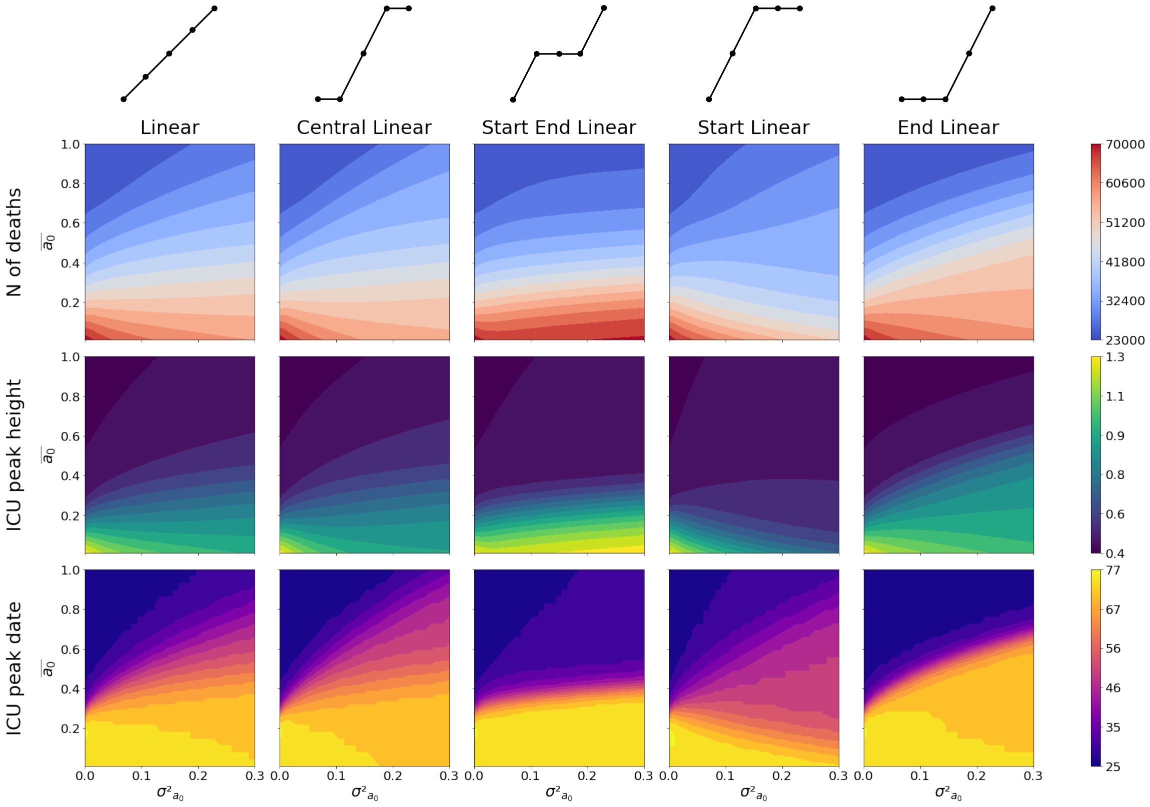

Figure S8: Heatmaps showing the number of deaths (first row), the height of the ICU peak (second row) and its date (third row) as a function of $\overline{a_0}$ and $\sigma^2_{a_0}$ with $\overline{b_0} = 0.5$ and $\sigma^2_{b_0} = 0$. Each column corresponds to one of the five functions linking perceived severity and midpoint of the logistic curve giving the transition rate from compliant to non-compliant compartments as a function of the fraction of vaccinated individuals. Above each column is a small diagram of the function, showing how the midpoint $a_0$ varies, going from small to high perceived severity groups (left to right). The other parameters used for the simulations are $\alpha = 10$ and $\gamma = 5$. We employed a 900-value grid, with 30 values of $\overline{a_0}$ ranging from 0 to 1, and 30 values of $\sigma^2_{a_0}$ ranging from 0 to 0.3. The rugged profile of the curves related to the peak date is due to its discrete nature, with values representing the (integer) number of days after the simulation's start in which the peak is observed.

Figure S8 shows the values of the three metrics as a function of $\overline{a_0}$ and $\sigma^2_{a_0}$ (similarly to Fig. 6 of the main text), for a smaller value of $\overline{b_0}$ (the midpoint of the logistic function concerning the transition rate from the non-compliant to the compliant compartments): $\overline{b_0} = 0.5$, with still $\sigma^2_{b_0} = 0$. A smaller $\overline{b_0}$ means that the increase of the transition rate to compliant compartments increases at a smaller occupancy of ICU. As a result, we observe the same overall phenomenology as in the main text, but with smaller values of the number of deaths and of the ICU peak height.

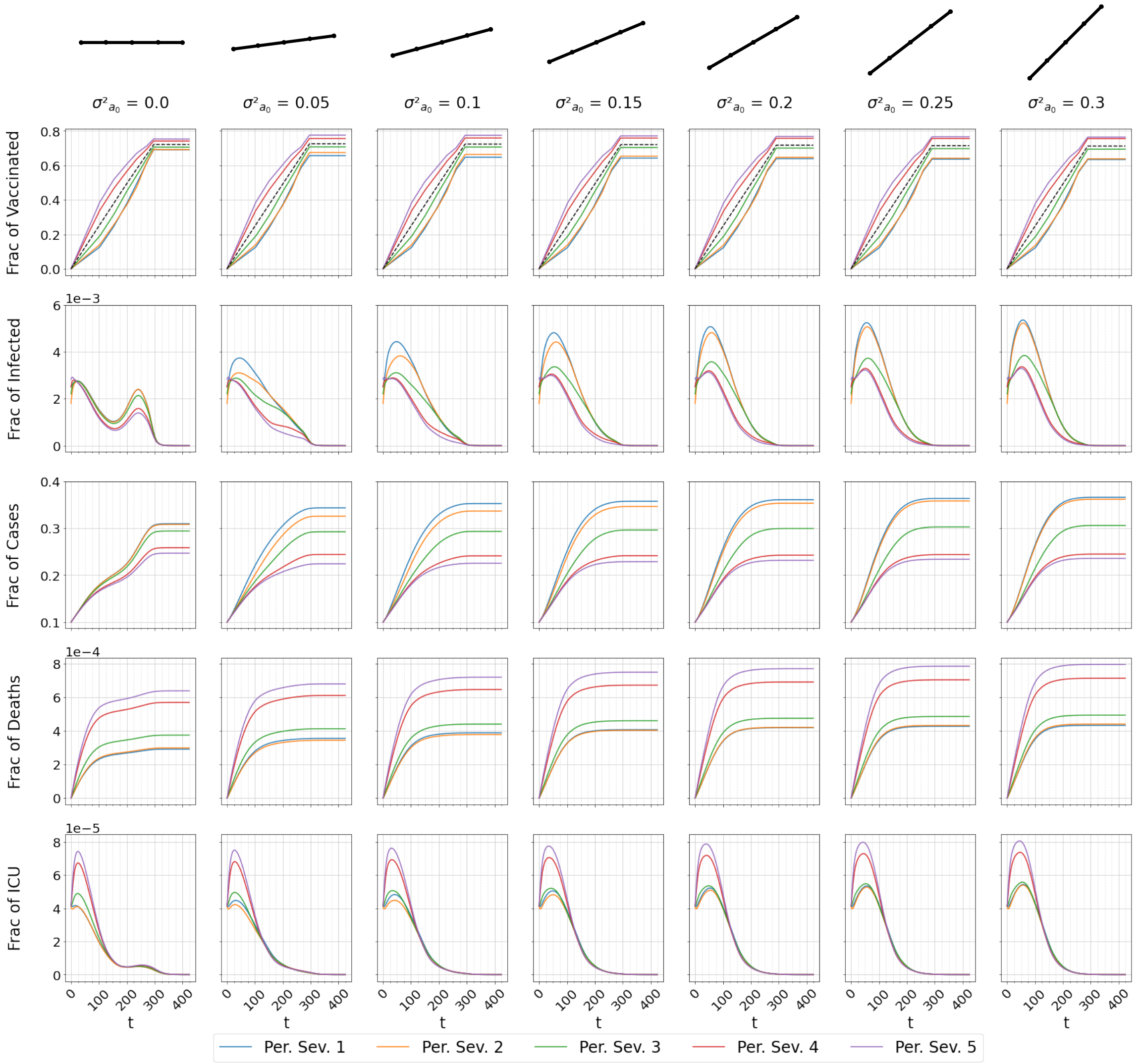

Figure S9: Fraction of vaccinated individuals (first row - the black dashed line reports the global fraction of vaccinated individuals in the population), infected individuals (second row), cases (third row - obtained as the sum of recovered individuals and deaths), deaths (fourth row) and individuals in ICU (fifth row) as a function of time (days), for each perceived severity group. Each column corresponds to a different value of the variance $\sigma^2_{a_0}$, going from 0 to 0.3, with $\overline{b_0} = 0.5$ and $\sigma^2_{b_0} = 0$. The other parameters are $\alpha = 10$, $\gamma = 5$, and $\overline{a_0} = 0.6$.

Figure S9 shows the epidemiological curves for $\overline{b_0} = 0.5$ and $\sigma^2_{b_0} = 0$, for $\overline{a_0} = 0.6$ and several values of $\sigma^2_{a_0}$ (similarly to Figure 7 of the main text). Here also we have a similar behaviour as for $\overline{b_0} = 0.75$, with slightly lower peaks.

Figures S10 and S11 show the results obtained with $\overline{b_0} = 1$ and $\sigma^2_{b_0} = 0$. The transition rates to compliant compartments increases then only slowly as the ICU fill up, resulting in higher peaks and higher final number of deaths, which can reach, respectively, a maximum of 1.9 times the maximum ICU capacity and 900,000 deaths. On the other hand however, the phenomenological picture as $\overline{a_0}$ and $\sigma^2_{a_0}$ vary is unchanged.

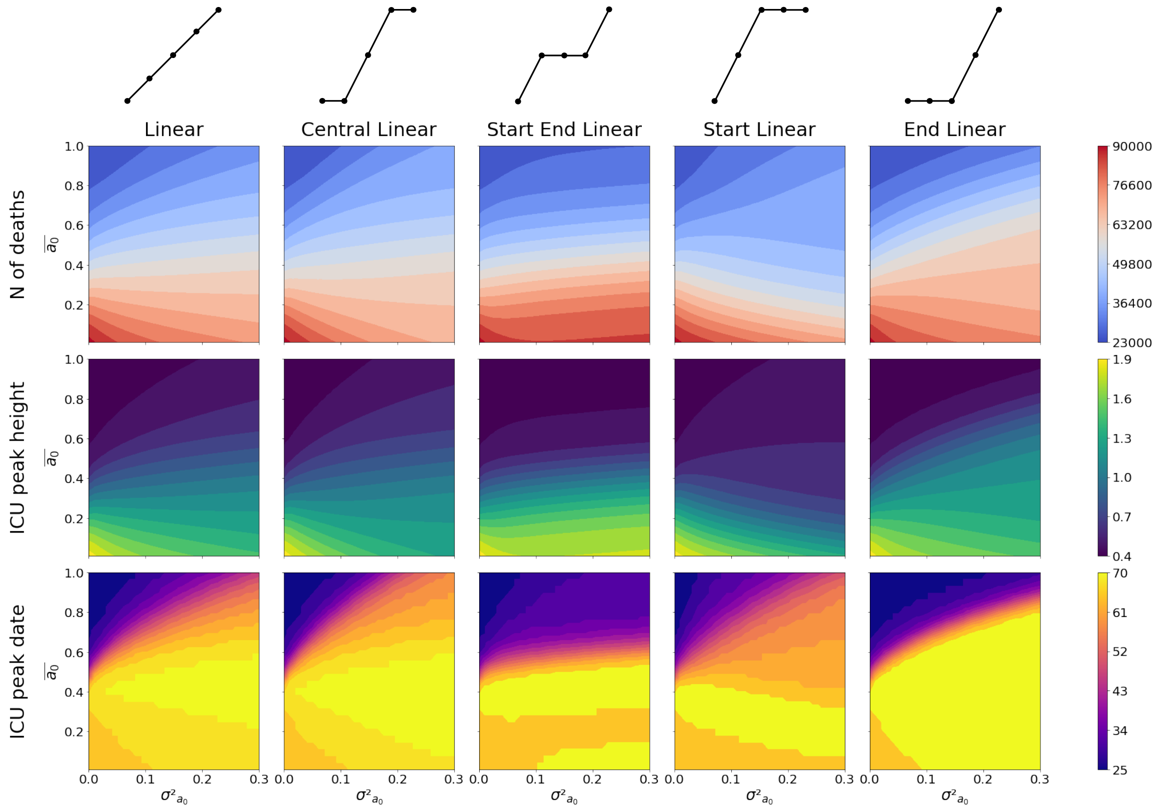

Figure S10: Heatmaps showing the number of deaths (first row), the height of the ICU peak (second row) and its date (third row) as a function of $\overline{a_0}$ and $\sigma^2_{a_0}$ with $\overline{b_0} = 1$ and $\sigma^2_{b_0} = 0$. Each column corresponds to one of the five functions linking perceived severity and midpoint of the logistic curve giving the transition rate from compliant to non-compliant compartments as a function of the fraction of vaccinated individuals. Above each column is a small diagram of the function, showing how the midpoint $a_0$ varies, going from small to high perceived severity groups (left to right). The other parameters used for the simulations are $\alpha = 10$ and $\gamma = 5$. We employed a 900-value grid, with 30 values of $\overline{a_0}$ ranging from 0 to 1, and 30 values of $\sigma^2_{a_0}$ ranging from 0 to 0.3. The rugged profile of the curves related to the peak date is due to its discrete nature, with values representing the (integer) number of days after the simulation's start in which the peak is observed.

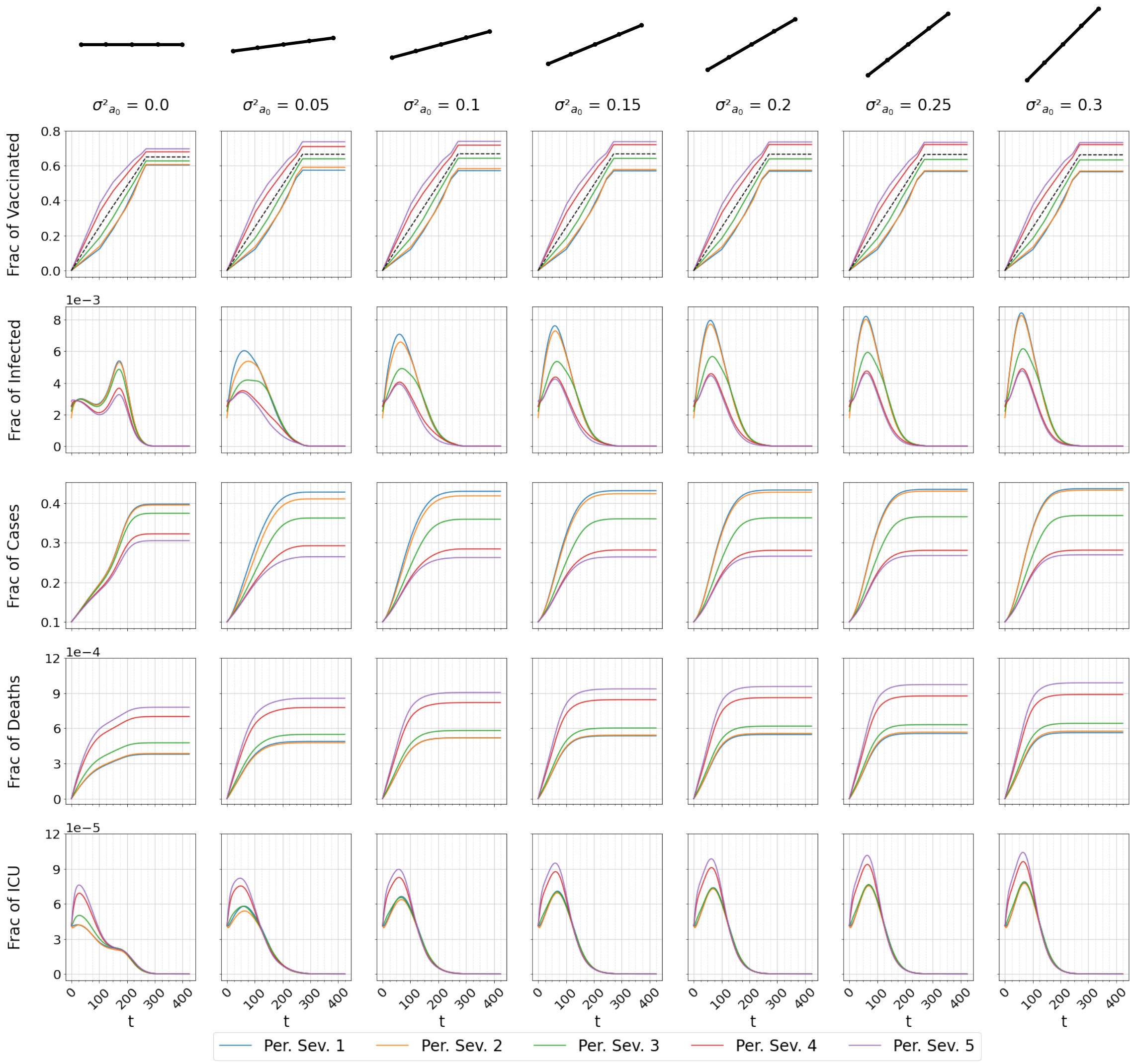

Figure S11: Fraction of vaccinated individuals (first row - the black dashed line reports the global fraction of vaccinated individuals in the population), infected individuals (second row), cases (third row - obtained as the sum of recovered individuals and deaths), deaths (fourth row) and individuals in ICU (fifth row) as a function of time (days), for each perceived severity group. Each column corresponds to a different value of the variance $\sigma^2_{a_0}$, going from 0 to 0.3, with $\overline{b_0} = 1$ and $\sigma^2_{b_0} = 0$. The other parameters are $\alpha = 10$, $\gamma = 5$, and $\overline{a_0} = 0.6$.

#### S3.4 Changing the slope of the logistic curves

We have used $\alpha = 10$ and $\gamma = 5$ as slopes of the logistic curves in the main text, in order to emulate a progressive relaxation of behaviors with the progress of the vaccination campaign (and also a re-adoption with the growth in ICU occupancy). Here, we consider different values for these slopes.

##### S3.4.1 Changing $\alpha$

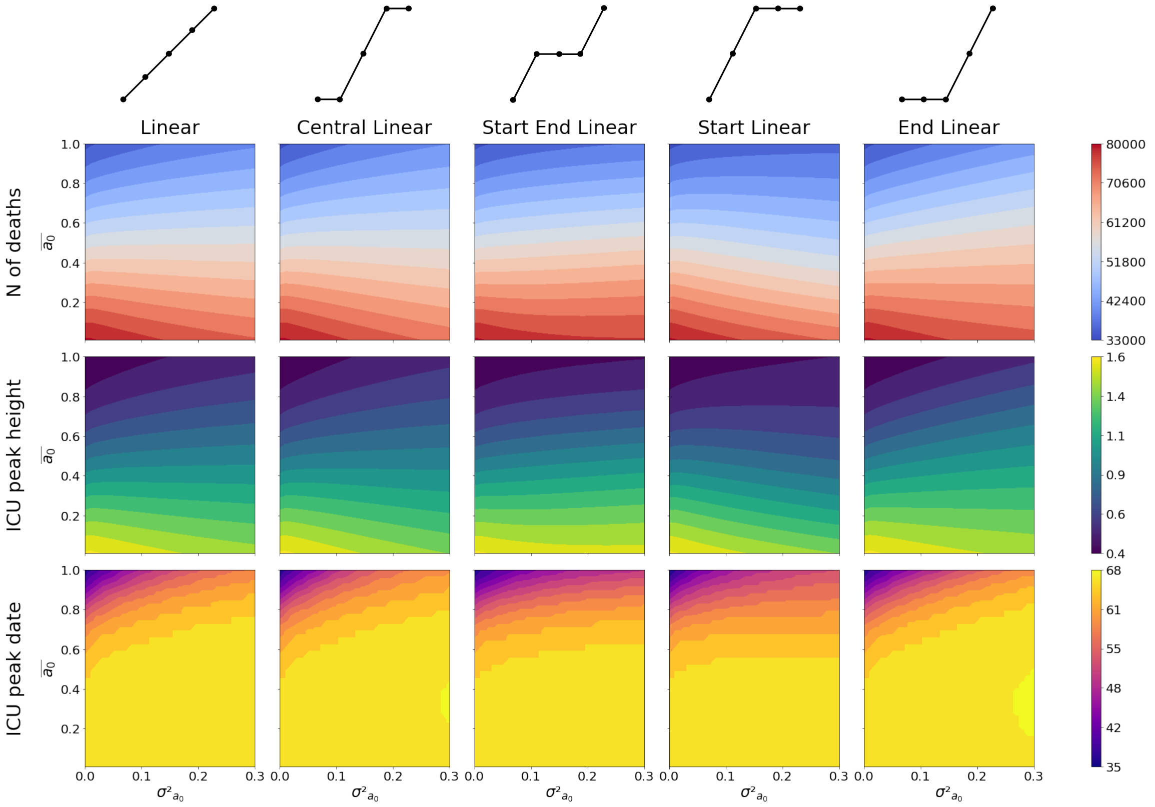

Figure S12: Heatmaps showing the number of deaths (first row), the height of the ICU peak (second row) and its date (third row) as a function of $\overline{a_0}$ and $\sigma^2_{a_0}$ with $\alpha = 4$. Each column corresponds to one of the five functions linking perceived severity and midpoint of the logistic curve giving the transition rate from compliant to non-compliant compartments as a function of the fraction of vaccinated individuals. Above each column is a small diagram of the function, showing how the midpoint $a_0$ varies, going from small to high perceived severity groups (left to right). The other parameters used for the simulations are $\gamma = 5$, $\overline{b_0} = 0.75$, and $\sigma^2_{b_0} = 0$. We employed a 900-value grid, with 30 values of $\overline{a_0}$ ranging from 0 to 1, and 30 values of $\sigma^2_{a_0}$ ranging from 0 to 0.3. The rugged profile of the curves related to the peak date is due to its discrete nature, with values representing the (integer) number of days after the simulation's start in which the peak is observed.

Figures S12 and S13 show the results for a smoother logistic curve, with $\alpha = 4$. In this case (see Figure 2 of the main text), the transition rate to the non-compliant compartment is larger even with no vaccinated individuals, and grows more gradually as the fraction of vaccinated increases. In such a case, a change in the midpoint value has only a limited impact on the value of the transition rate, at given fraction of vaccinated. Thus, increasing the variance also has a smaller impact on the metrics considered and on the dynamics of the epidemics with respect to the case investigated in the main text.

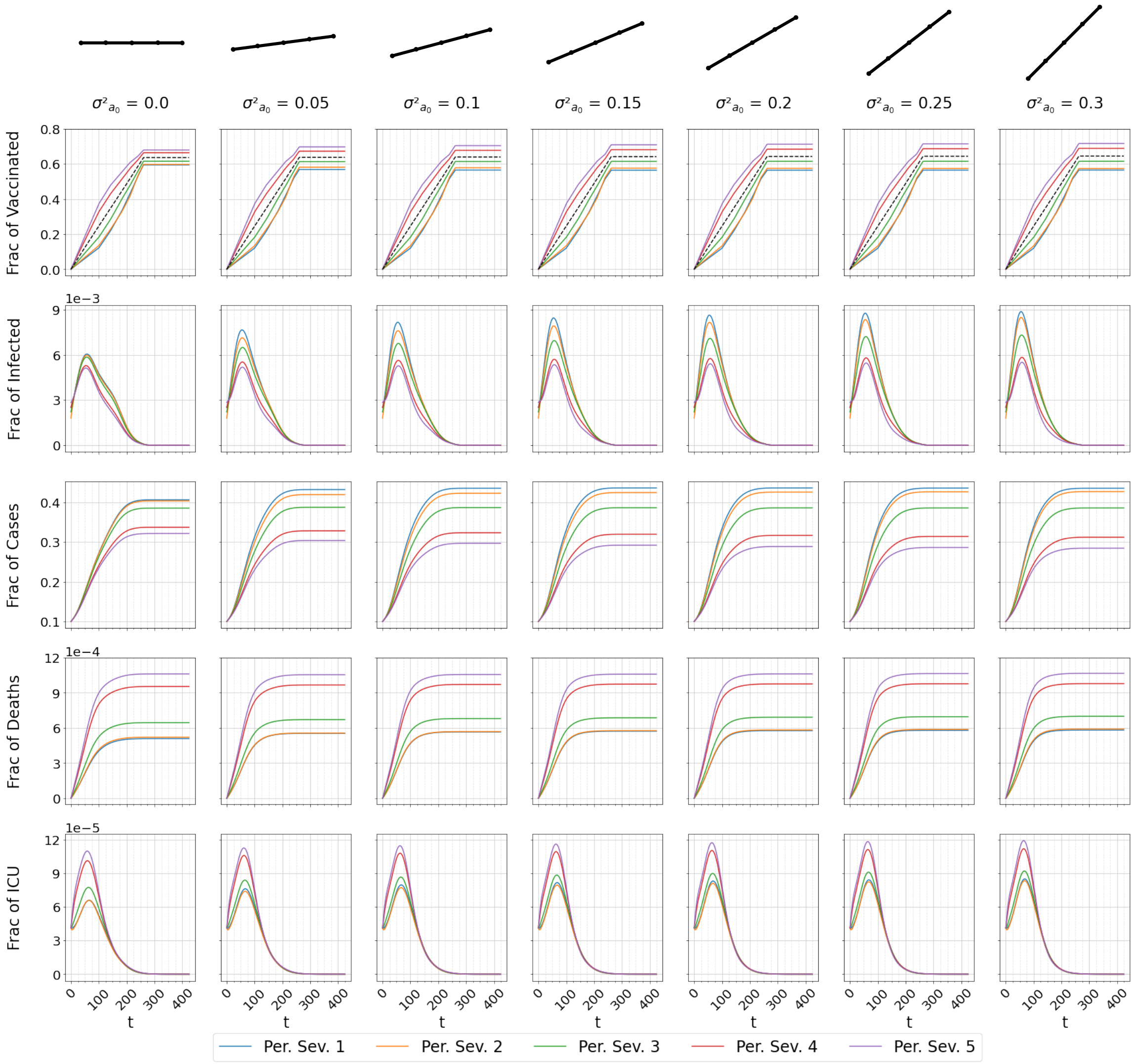

Figure S13: Fraction of vaccinated individuals (first row - the black dashed line reports the global fraction of vaccinated individuals in the population), infected individuals (second row), cases (third row - obtained as the sum of recovered individuals and deaths), deaths (fourth row) and individuals in ICU (fifth row) as a function of time (days), for each perceived severity group. Each column corresponds to a different value of the variance $\sigma^2_{a_0}$, going from 0 to 0.3, with $\alpha = 4$. The other parameters are $\gamma = 5$, $\overline{a_0} = 0.6$, $\overline{b_0} = 0.75$, and $\sigma^2_{b_0} = 0$.

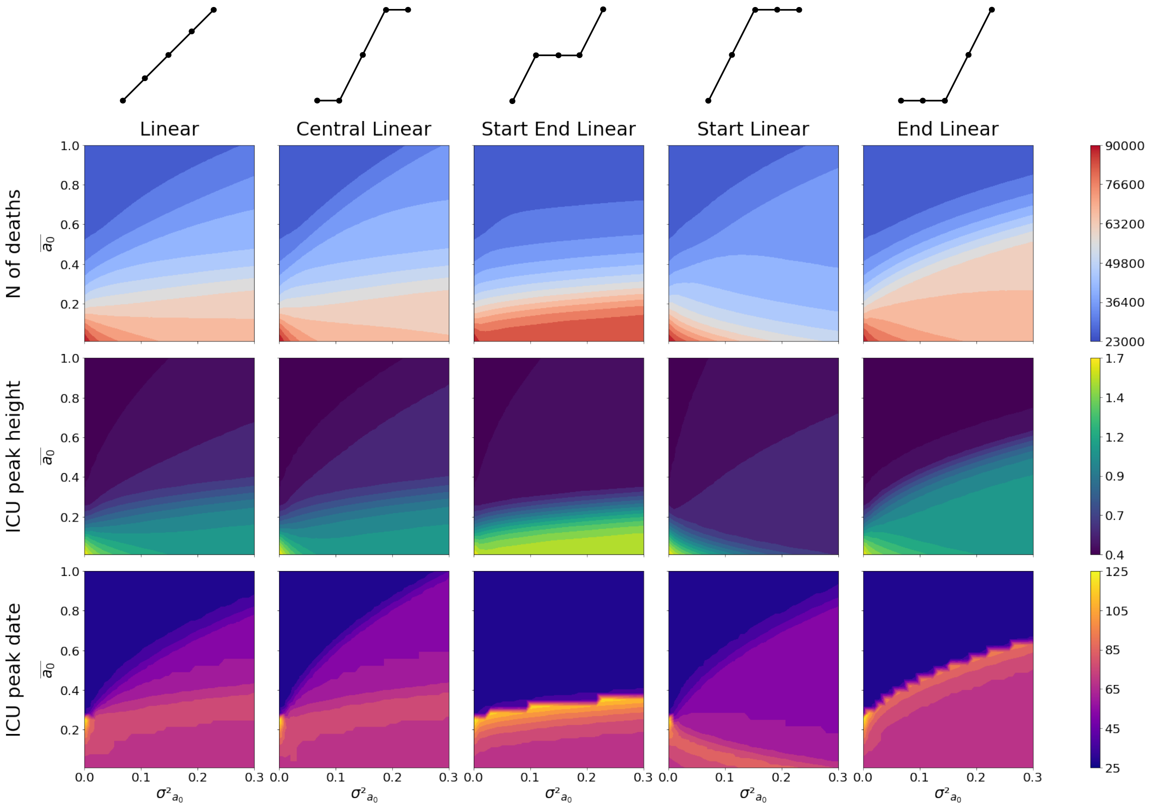

Figure S14: Heatmaps showing the number of deaths (first row), the height of the ICU peak (second row) and its date (third row) as a function of $\overline{a_0}$ and $\sigma^2_{a_0}$ with $\alpha = 20$. Each column corresponds to one of the five functions linking perceived severity and midpoint of the logistic curve giving the transition rate from compliant to non-compliant compartments as a function of the fraction of vaccinated individuals. Above each column is a small diagram of the function, showing how the midpoint $a_0$ varies, going from small to high perceived severity groups (left to right). The other parameters used for the simulations are $\gamma = 5$, $\overline{b_0} = 0.75$, and $\sigma^2_{b_0} = 0$. We employed a 900-value grid, with 30 values of $\overline{a_0 a_0}$ and ranging from 0 to 1, and 30 values of $\sigma^2_{a_0}$ ranging from 0 to 0.3. The rugged profile of the curves related to the peak date is due to its discrete nature, with values representing the (integer) number of days after the simulation's start in which the peak is observed.

For high values of the slope $\alpha$ on the contrary, the transition rate evolves much faster when the fraction of vaccinated is near the midpoint. Vice versa, away from the midpoint an increase in the fraction of vaccinated individuals does not change much the value of the transition rate from compliant to non-compliant behaviors (see Figure 2 of the main text). The impact on the ICU peak height and on the final number of deaths is limited (Figure S14 with $\alpha = 20$). A stronger effect is seen in the peak date and in the temporal evolution of the fraction of infected (Figure S15). In particular, the second peak for $\sigma^2_{a_0} = 0$ is much narrower with respect to the case of $\alpha = 10$, as the increase in the transition rate towards non-compliant compartments, and thus the relaxation of behaviors, takes place over a shorter timescale. This second peak can even become higher than the first. Despite these differences, we observe also here the same phenomenology as before when the parameters $\overline{a_0}$ and $\sigma^2_{a_0}$ vary, in particular with the disappearance of the second peak for $\sigma^2_{a_0} > 0$.

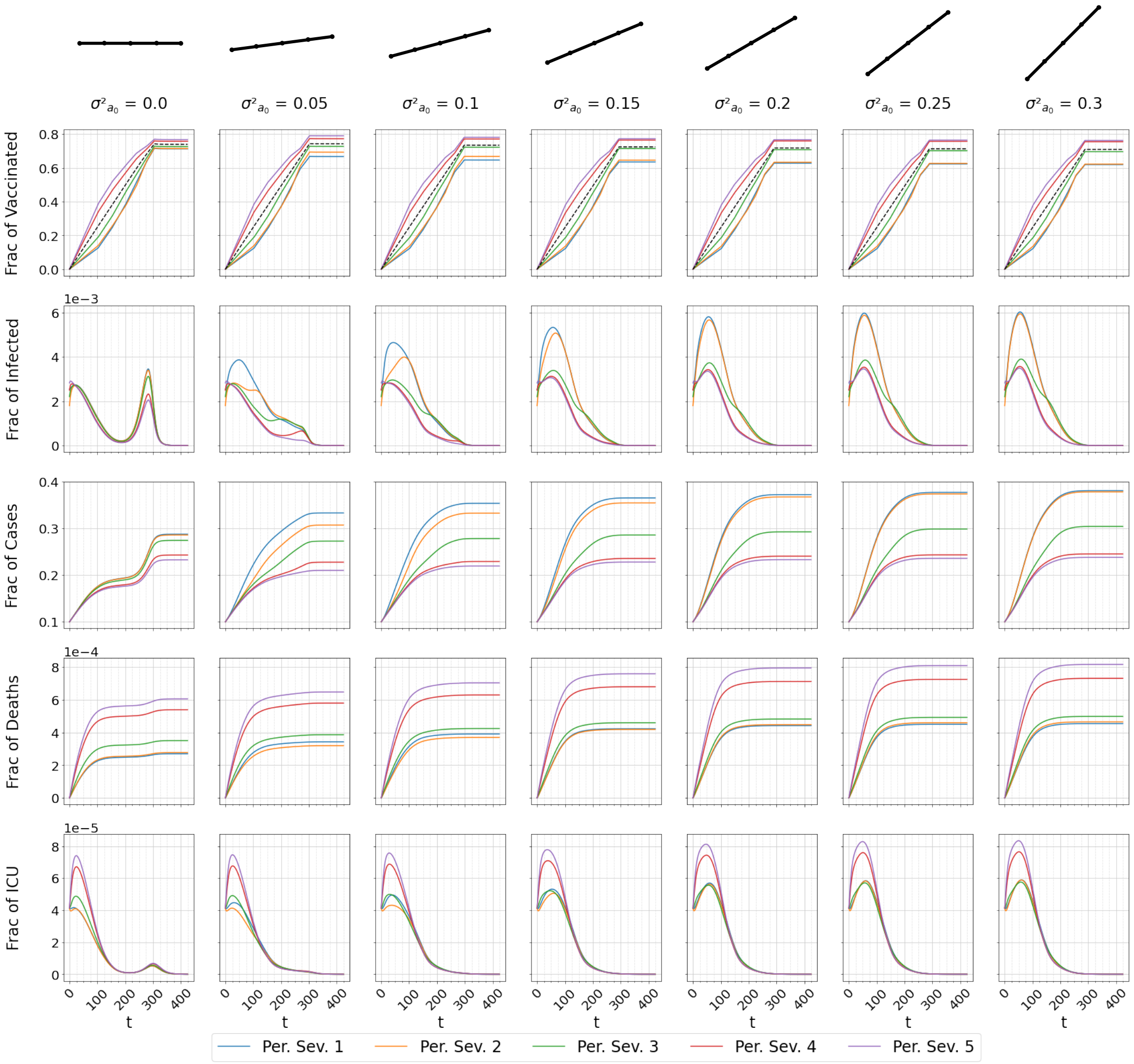

Figure S15: Fraction of vaccinated individuals (first row - the black dashed line reports the global fraction of vaccinated individuals in the population), infected individuals (second row), cases (third row - obtained as the sum of recovered individuals and deaths), deaths (fourth row) and individuals in ICU (fifth row) as a function of time (days), for each perceived severity group. Each column corresponds to a different value of the variance $\sigma^2_{a_0}$, going from 0 to 0.3, with $\alpha = 20$. The other parameters are $\gamma = 5$, $\overline{a_0} = 0.6$, $\overline{b_0} = 0.75$, and $\sigma^2_{b_0} = 0$.

##### S3.4.2 Changing $\gamma$

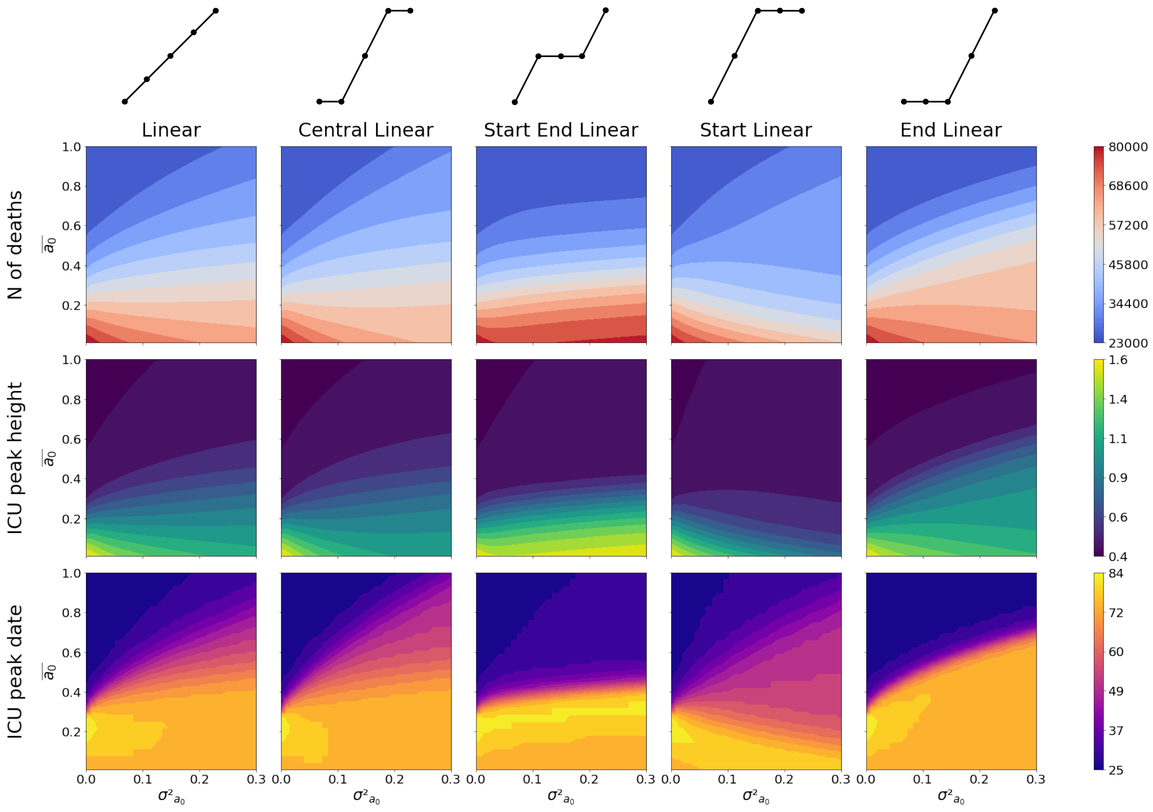

Figure S16: Heatmaps showing the number of deaths (first row), the height of the ICU peak (second row) and its date (third row) as a function of $\overline{a_0}$ and $\sigma^2_{a_0}$ with $\gamma = 2$. Each column corresponds to one of the five functions linking perceived severity and midpoint of the logistic curve giving the transition rate from compliant to non-compliant compartments as a function of the fraction of vaccinated individuals. Above each column is a small diagram of the function, showing how the midpoint $a_0$ varies, going from small to high perceived severity groups (left to right). The other parameters used for the simulations are $\alpha = 10$, $\overline{b_0} = 0.75$, and $\sigma^2_{b_0} = 0$. We employed a 900-value grid, with 30 values of $\overline{a_0}$ ranging from 0 to 1, and 30 values of $\sigma^2_{a_0}$ ranging from 0 to 0.3. The rugged profile of the curves related to the peak date is due to its discrete nature, with values representing the (integer) number of days after the simulation's start in which the peak is observed.

Figures S16 and S17 show the results for a small value of the slope $\gamma = 2$. In this case, the transition rate to compliant compartments is high even when the ICU are empty, and this rate has a weak dependency on the ICU occupancy. As a result, the global impact of the spread is smaller than in the case considered in the main text, at given $(\overline{a_0}, \sigma^2_{a_0})$. Moreover, the second peak of infected when $\sigma^2_{a_0} = 0$ is very small if compared to the ones obtained in the main text, and the resulting bump in the curve of the ICU occcupancy vs time is also very small.

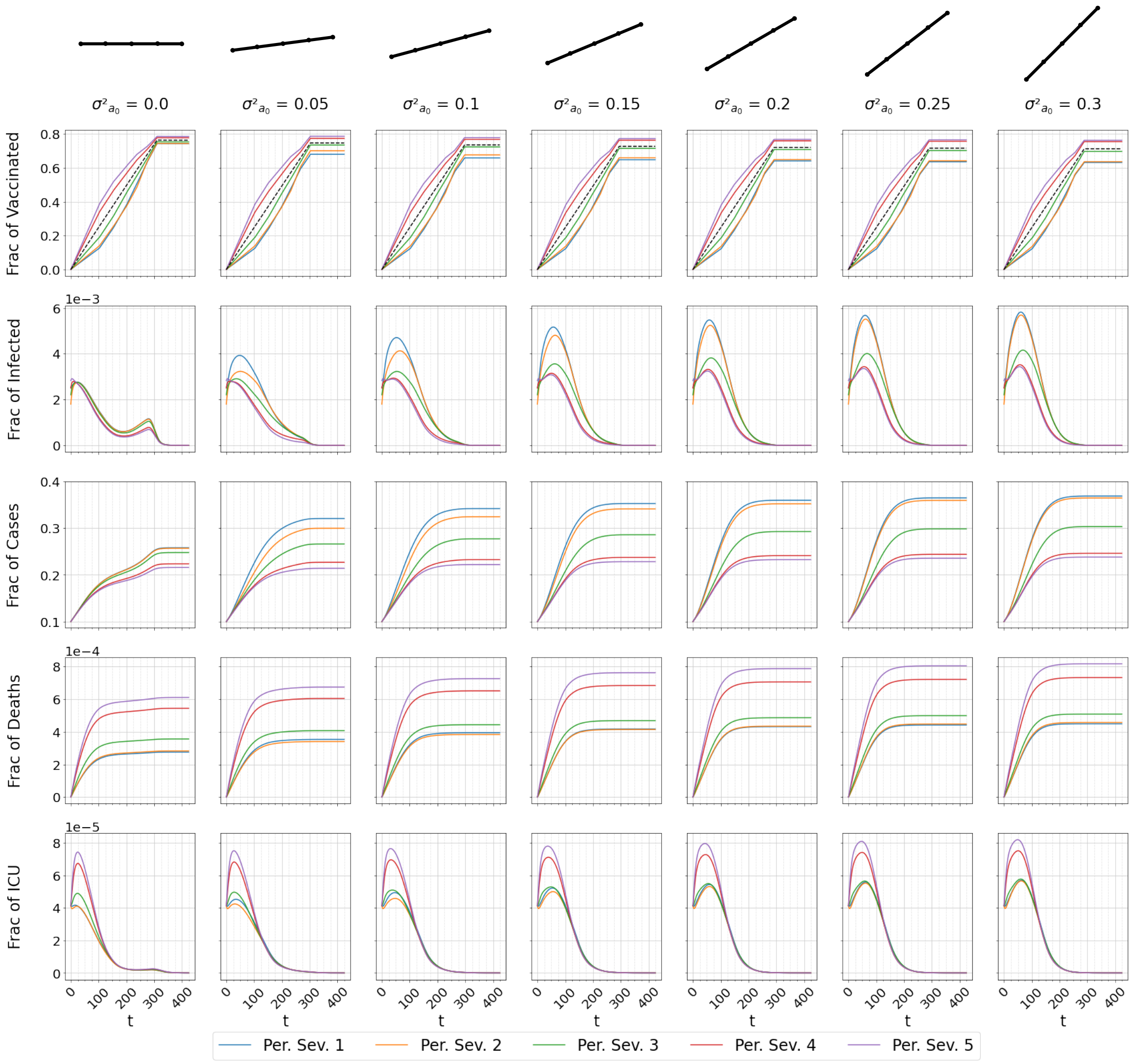

Figure S17: Fraction of vaccinated individuals (first row - the black dashed line reports the global fraction of vaccinated individuals in the population), infected individuals (second row), cases (third row - obtained as the sum of recovered individuals and deaths), deaths (fourth row) and individuals in ICU (fifth row) as a function of time (days), for each perceived severity group. Each column corresponds to a different value of the variance $\sigma^2_{a_0}$, going from 0 to 0.3, with $\gamma = 2$. The other parameters are $\alpha = 10$, $\overline{a_0} = 0.6$, $\overline{b_0} = 0.75$, and $\sigma^2_{b_0} = 0$.

Figures S18 and S19 show the case of a high slope $\gamma = 15$. The transition rate to compliant compartments is then very small until the ICU occupancy reaches a midpoint, so that a large part of the population remains non-compliant, leading to higher ICU peak and a larger number of deaths than for smaller $\gamma$. The overall picture when $\overline{a_0}$ and $\sigma^2_{a_0}$ remains similar.

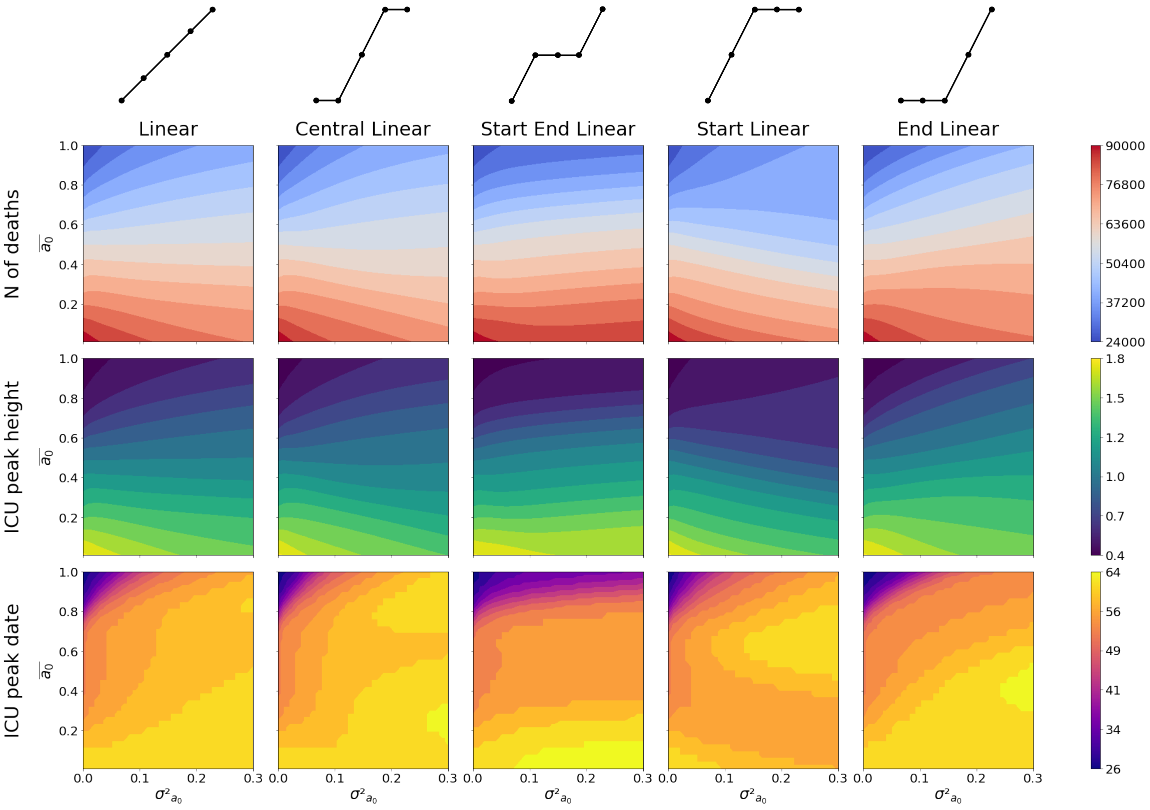

Figure S18: Heatmaps showing the number of deaths (first row), the height of the ICU peak (second row) and its date (third row) as a function of $\overline{a_0}$ and $\sigma^2_{a_0}$ with $\gamma = 15$. Each column corresponds to one of the five functions linking perceived severity and midpoint of the logistic curve giving the transition rate from compliant to non-compliant compartments as a function of the fraction of vaccinated individuals. Above each column is a small diagram of the function, showing how the midpoint $a_0$ varies, going from small to high perceived severity groups (left to right). The other parameters used for the simulations are $\alpha = 10$, $\overline{b_0} = 0.75$, and $\sigma^2_{b_0} = 0$. We employed a 900-value grid, with 30 values of $\overline{a_0}$ ranging from 0 to 1, and 30 values of $\sigma^2_{a_0}$ ranging from 0 to 0.3. The rugged profile of the curves related to the peak date is due to its discrete nature, with values representing the (integer) number of days after the simulation's start in which the peak is observed.

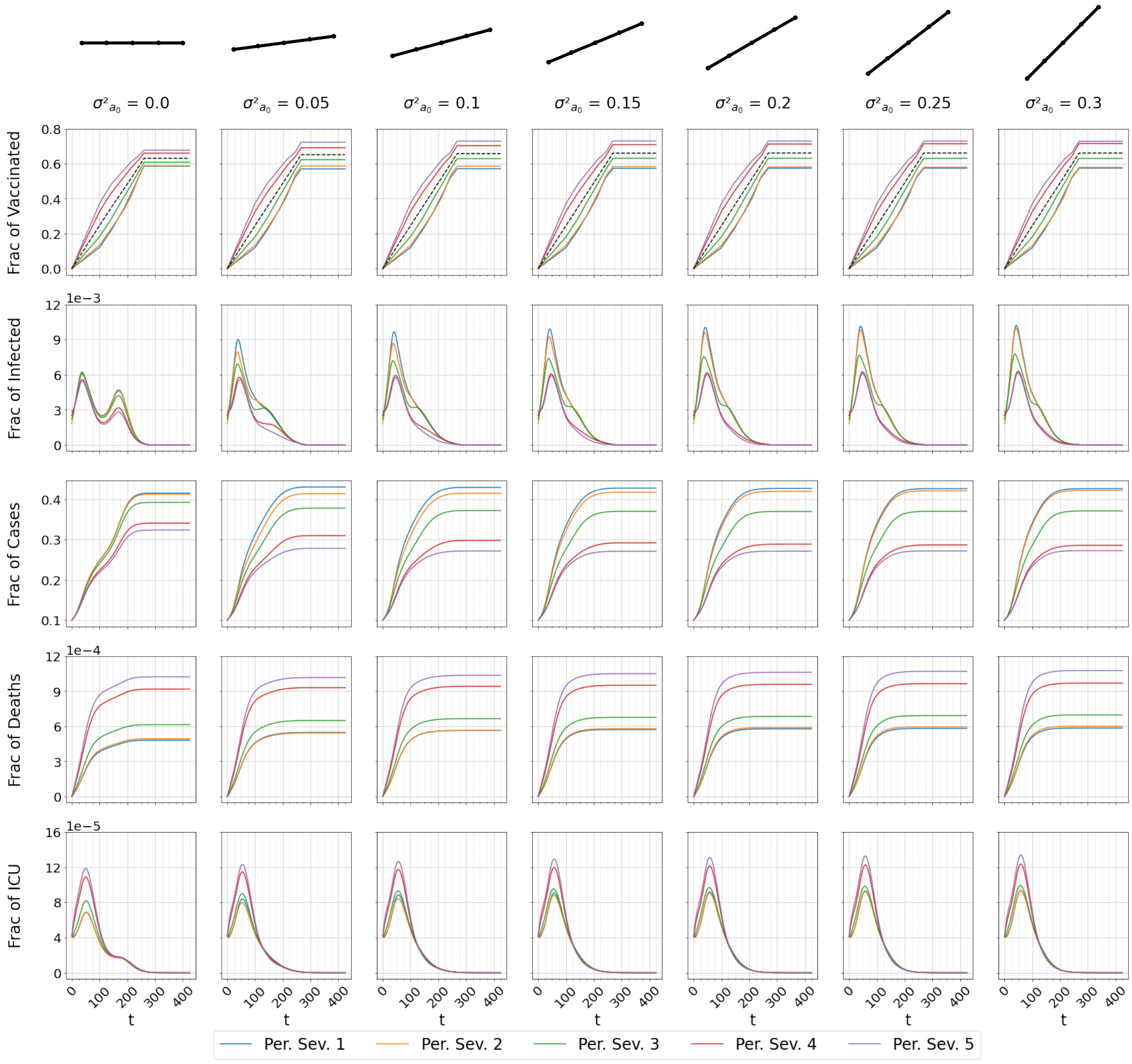

Figure S19: Fraction of vaccinated individuals (first row - the black dashed line reports the global fraction of vaccinated individuals in the population), infected individuals (second row), cases (third row - obtained as the sum of recovered individuals and deaths), deaths (fourth row) and individuals in ICU (fifth row) as a function of time (days), for each perceived severity group. Each column corresponds to a different value of the variance $\sigma^2_{a_0}$, going from 0 to 0.3, with $\gamma = 15$. The other parameters are $\alpha = 10$, $\overline{a_0} = 0.6$, $\overline{b_0} = 0.75$, and $\sigma^2_{b_0} = 0$.

#### S3.5 Transition from non-compliant to compliant compartments depending on perceived severity

In this section, we explore a scenario where the logistic curve giving the transition rate from non-compliant to compliant parameters, ruled by the parameters $\gamma$ and $b_0$, depends on perceived severity. Therefore, we fix the variance $\sigma^2_{a_0}$ to 0 and we analyze the effect of introducing heterogeneities in the re-adoption of protective measures by different perceived severity groups, by varying $\overline{b_0}$ and $\sigma^2_{b_0}$.

In Figure S20 we show the results for the three metrics as a function of $\overline{b_0}$ and $\sigma^2_{b_0}$ for the five functions, fixing the slopes $\alpha = 10$, $\gamma = 5$ and the midpoint of the logistic curve describing the transition rate from C to NC to $\overline{a_0} = 0.6$. The heatmaps are very similar for all five functions. Increasing the mean value of the midpoint $\overline{b_0}$ results in a higher number of deaths and a later ICU peak. This is due to the fact that, if $\overline{b_0}$ increases, the rate of re-adoption of a compliant behavior increases only for larger ICU occupancy: the non-compliant behavior is thus favored, with more contacts and an increased propagation. Increasing the variance $\sigma^2_{b_0}$ leads to increased heterogeneities among perceived severity groups, resulting in higher numbers of deaths and a delayed peak, for all $\overline{b_0}$. As for the increase in $\sigma^2_{a_0}$, the increase in $\sigma^2_{b_0}$ has in fact a priori two opposite effects: here groups with higher perceived severity tend to revert more easily to a compliant behavior, while the groups with lower perceived severity maintain a non-compliant behavior even at high full ICU occupancy. Overall, for this value of $\overline{a_0} = 0.6$, the main effect is that the non-compliance of these latter groups facilitates initial virus spread, leading to a higher initial peak and the disappearance of the second peak, as seen in the epidemiological curves in Figure S21.

Finally, we quantify the differences between the heatmaps shown in Figure S21 through their Canberra distances, given in Table S1. As in the case studied in the main text, we obtain very small values (i.e. $< 0.1$).

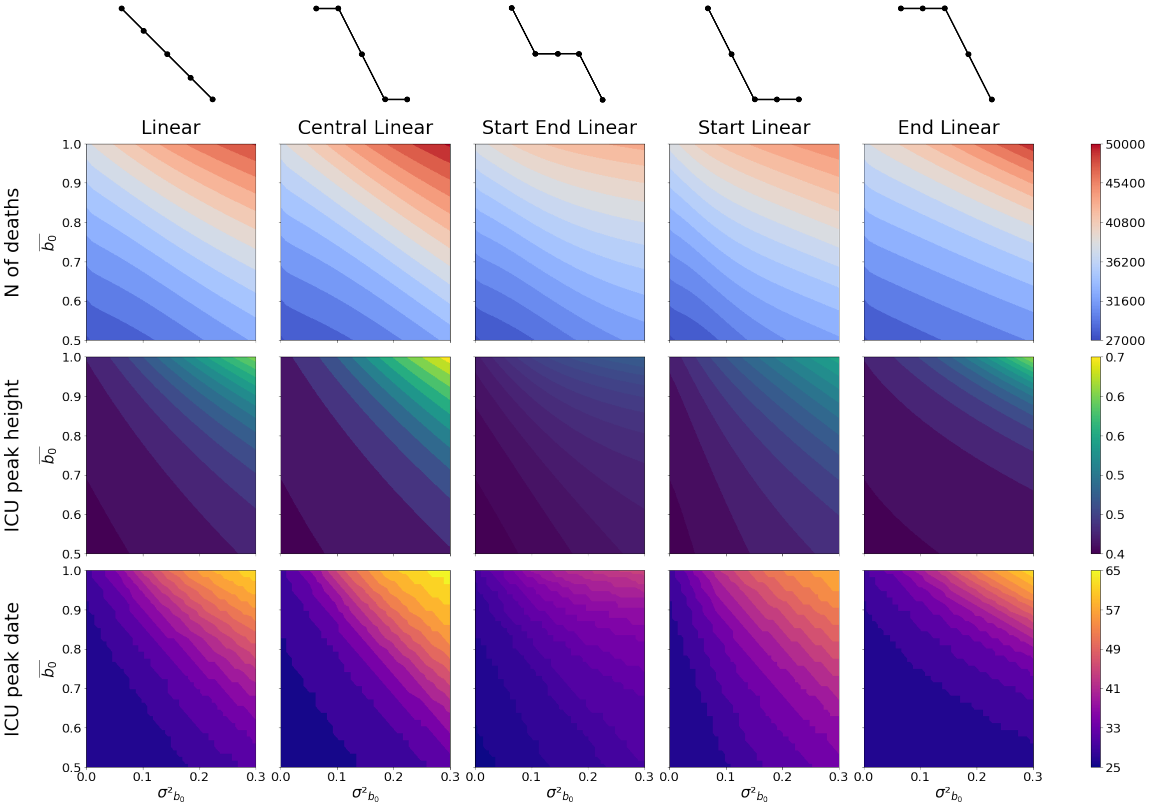

Figure S20: Heatmaps showing the number of deaths (first row), the height of the ICU peak (second row) and its date (third row) as a function of $\overline{b_0}$ and $\sigma^2_{b_0}$ with $\overline{a_0} = 0.6$ and $\sigma^2_{a_0} = 0$. Each column corresponds to one of the five functions linking perceived severity and midpoint of the logistic curve giving the transition rate from compliant to non-compliant compartments as a function of the fraction of vaccinated individuals. Above each column is a small diagram of the function, showing how the midpoint $a_0$ varies, going from small to high perceived severity groups (left to right). The other parameters used for the simulations are $\alpha = 10$ and $\gamma = 5$. We employed a 900-value grid, with 30 values of $\overline{b_0}$ ranging from 0.5 to 1, and 30 values of $\sigma^2_{b_0}$ ranging from 0 to 0.3. The rugged profile of the curves related to the peak date is due to its discrete nature, with values representing the (integer) number of days after the simulation's start in which the peak is observed.

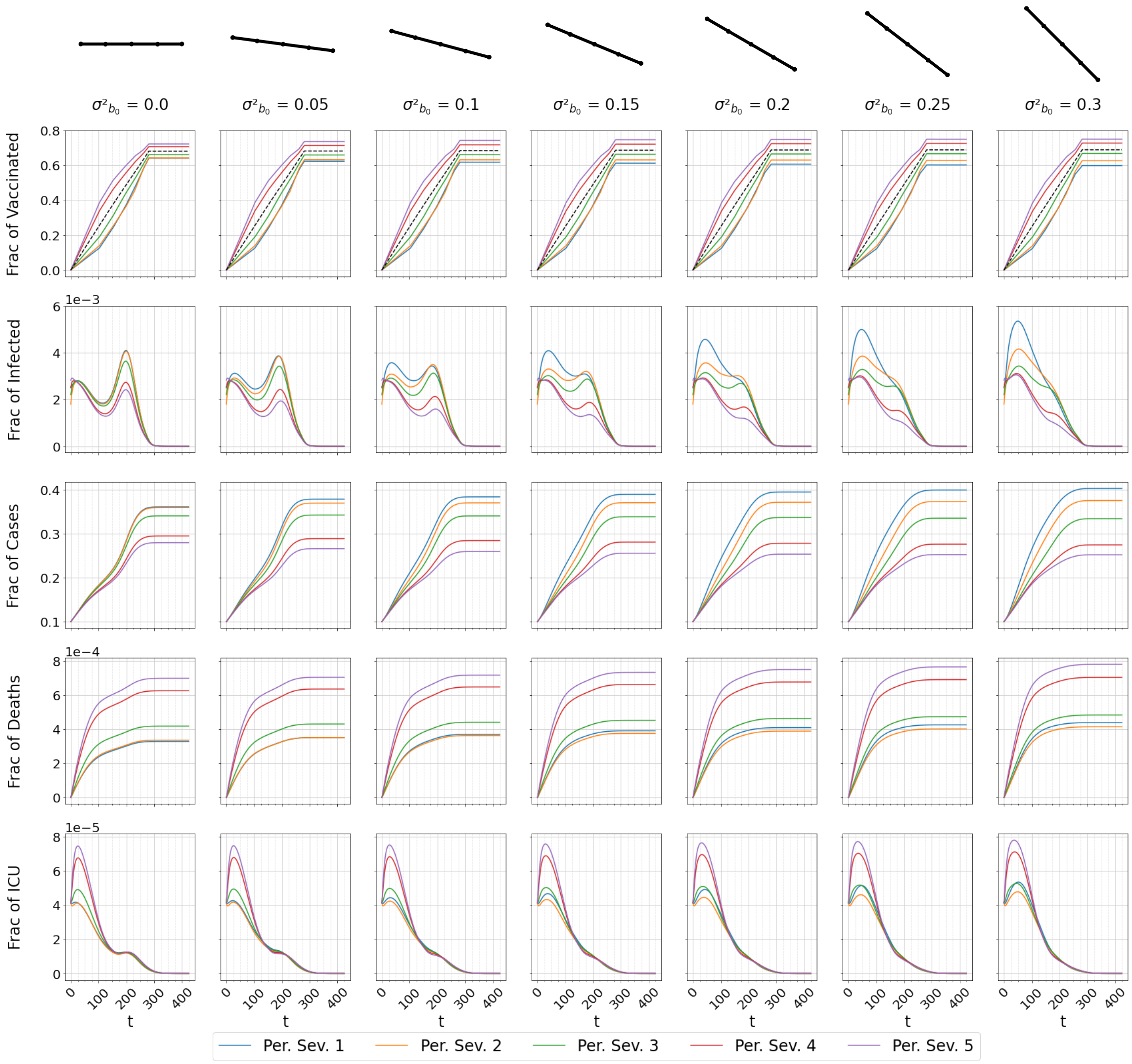

Figure S21: Fraction of vaccinated individuals (first row - the black dashed line reports the global fraction of vaccinated individuals in the population), infected individuals (second row), cases (third row - obtained as the sum of recovered individuals and deaths), deaths (fourth row) and individuals in ICU (fifth row) as a function of time (days), for each perceived severity group. Each column corresponds to a different value of the variance $\sigma^2_{b_0}$, going from 0 to 0.3, with $\overline{a_0} = 0.6$ and $\sigma^2_{a_0} = 0$. The other parameters are $\alpha = 10$, $\gamma = 5$, $\overline{b_0} = 0.8$.

(a) Canberra distance - Number of deaths

| | Linear | Central Linear | Start End Linear | Start Linear | End Linear |
|---|---|---|---|---|---|
| Linear | | 0.0038 | 0.0091 | 0.0050 | 0.0078 |
| Central Linear | 0.0038 | | 0.0126 | 0.0086 | 0.0112 |
| Start End Linear | 0.0091 | 0.0126 | | 0.0073 | 0.0058 |
| Start Linear | 0.0050 | 0.0086 | 0.0073 | | 0.0083 |
| End Linear | 0.0078 | 0.0112 | 0.0058 | 0.0083 | |

(b) Canberra distance - Peak height

| | Linear | Central Linear | Start End Linear | Start Linear | End Linear |
|---|---|---|---|---|---|
| Linear | | 0.0050 | 0.0089 | 0.0045 | 0.0070 |
| Central Linear | 0.0050 | | 0.0139 | 0.0075 | 0.0120 |
| Start End Linear | 0.0089 | 0.0139 | | 0.0108 | 0.0054 |
| Start Linear | 0.0045 | 0.0075 | 0.0108 | | 0.0097 |
| End Linear | 0.0070 | 0.0120 | 0.0054 | 0.0097 | |

(c) Canberra distance - Peak date

| | Linear | Central Linear | Start End Linear | Start Linear | End Linear |
|---|---|---|---|---|---|
| Linear | | 0.0177 | 0.0466 | 0.0254 | 0.0441 |
| Central Linear | 0.0177 | | 0.0639 | 0.0289 | 0.0613 |
| Start End Linear | 0.0466 | 0.0639 | | 0.0669 | 0.0274 |
| Start Linear | 0.0254 | 0.0289 | 0.0669 | | 0.0651 |
| End Linear | 0.0441 | 0.0613 | 0.0274 | 0.0651 | |

Table S1: Canberra distance among each pair of functions for the three metrics considered: number of deaths (a), peak height (b) and peak date (c). We used the grid of values of Figure S20 for $\overline{b_0}$ and $\sigma^2_{b_0}$. The other parameters are $\alpha = 10$, $\gamma = 5$, $\overline{a_0} = 0.6$, $\sigma^2_{a_0} = 0$.

We present in Figure S22 the heatmaps for a smaller midpoint $\overline{a_0} = 0.3$. In this case, the number of deaths and the ICU peak height are larger, as the transition rate to non-compliant compartments increases earlier when the fraction of population that is vaccinated increases. However, the overall trend remains similar: an increase in the variance or mean value of $\overline{b_0}$ leads to higher number of deaths and ICU peak. An exception is observed with the Start Linear function, where a very high midpoint and increased variance initially result in reduced deaths and ICU occupancy, followed by an eventual increase. Regarding the peak date, there is a noticeable difference in the level curves for small and high values of $\overline{b_0}$. Specifically, for $\overline{b_0}$ values below 0.75, increasing the variance delays the peak, as observed in Figure S20 with $\overline{a_0} = 0.6$. However, for $\overline{b_0}$ values above 0.75, three of the five functions anticipate the peak with increased variance, while the other two show either fluctuations around a value (Start End Linear) or an anticipation of the peak only for higher values of $\overline{b_0}$. For completeness, in Figure S23, we provide the epidemiological curves with $\overline{a_0} = 0.3$ and $\overline{b_0} = 0.8$. The earlier relaxation of behaviors due to the small value of $\overline{a_0}$ causes the presence of a single peak also with $\sigma^2_{b_0} = 0$. An increase in the variance leads to shorter but higher peaks, both for the infections and the ICU occupancy. The Canberra Distances among the pairs of functions take very small values as in the previous case.

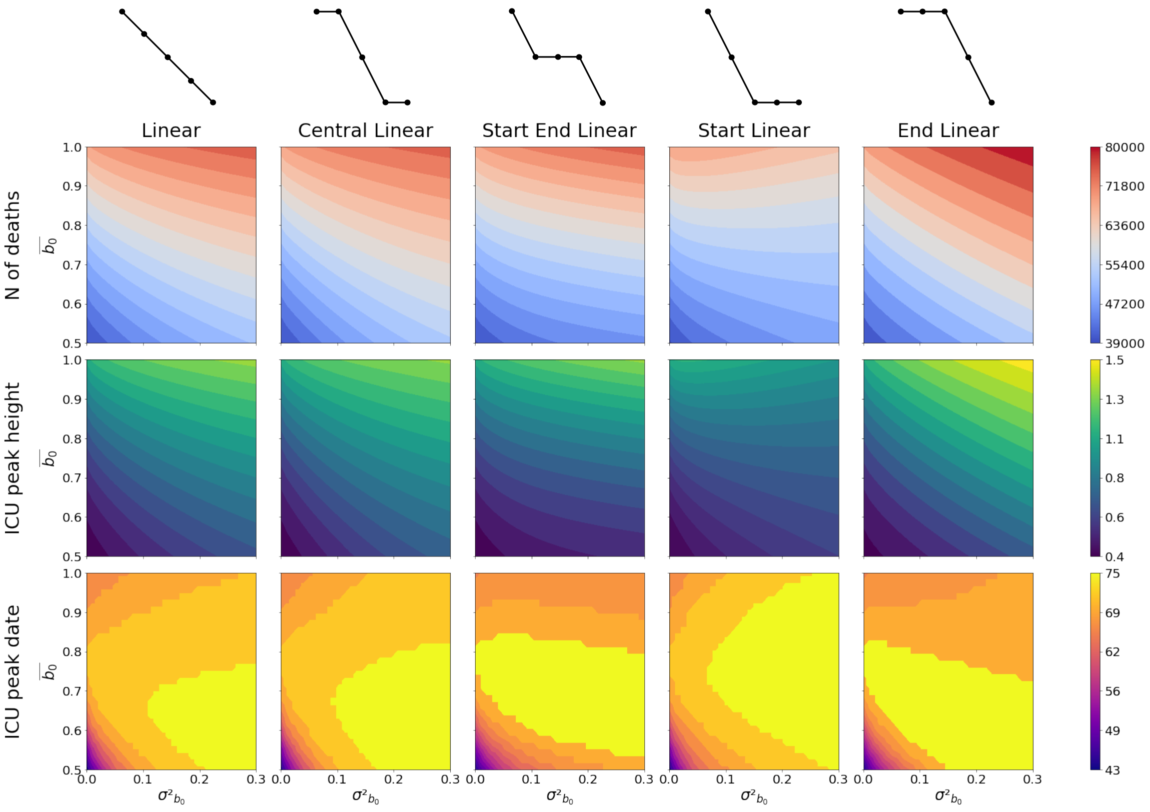

Figure S22: Heatmaps showing the number of deaths (first row), the height of the ICU peak (second row) and its date (third row) as a function of $\overline{b_0}$ and $\sigma^2_{b_0}$ with $\overline{a_0} = 0.3$ and $\sigma^2_{a_0} = 0$. Each column corresponds to one of the five functions linking perceived severity and midpoint of the logistic curve giving the transition rate from compliant to non-compliant compartments as a function of the fraction of vaccinated individuals. Above each column is a small diagram of the function, showing how the midpoint $a_0$ varies, going from small to high perceived severity groups (left to right). The other parameters used for the simulations are $\alpha = 10$ and $\gamma = 5$. We employed a 900-value grid, with 30 values of $\overline{b_0}$ ranging from 0.5 to 1, and 30 values of $\sigma^2_{b_0}$ ranging from 0 to 0.3. The rugged profile of the curves related to the peak date is due to its discrete nature, with values representing the (integer) number of days after the simulation's start in which the peak is observed.

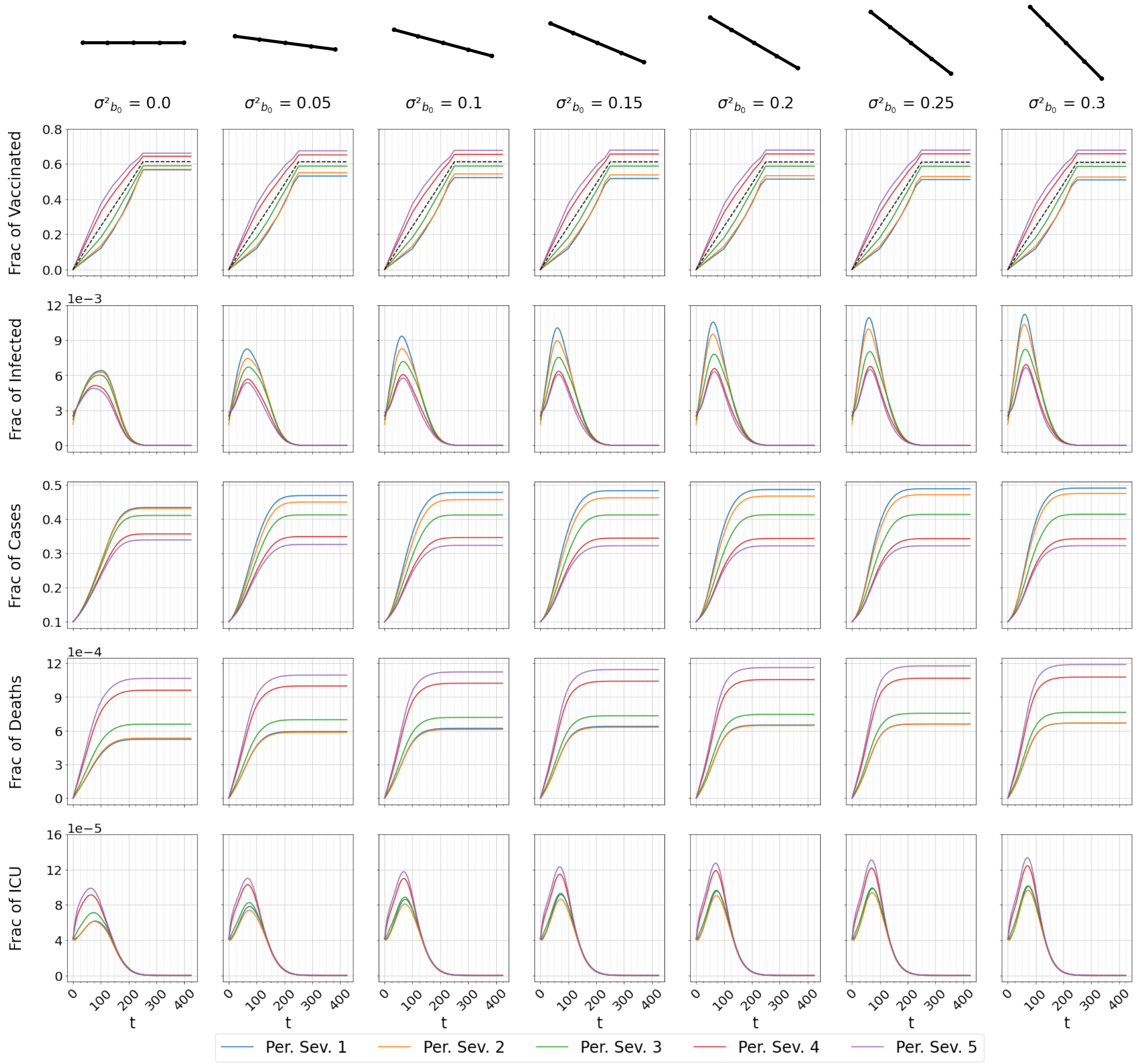

Figure S23: Fraction of vaccinated individuals (first row - the black dashed line reports the global fraction of vaccinated individuals in the population), infected individuals (second row), cases (third row - obtained as the sum of recovered individuals and deaths), deaths (fourth row) and individuals in ICU (fifth row) as a function of time (days), for each perceived severity group. Each column corresponds to a different value of the variance $\sigma^2_{b_0}$, going from 0 to 0.3, with $\overline{a_0} = 0.3$ and $\sigma^2_{a_0} = 0$. The other parameters are $\alpha = 10$, $\gamma = 5$, $\overline{b_0} = 0.8$.

#### S3.6 Both transition rates depending on perceived severity

In the previous sections, we introduced variations in behaviors based on perceived severity for only one of the two logistic curves describing the transition rates. In this section, we explore a scenario in which both transitions depend on perceived severity. In this case thus, both $\sigma^2_{a_0}$ and $\sigma^2_{b_0}$ are non-zero.

Figure S24 shows the temporal evolution of the fraction of infected for various values of $\sigma^2_{a_0}$ (x-axis) and $\sigma^2_{b_0}$ (y-axis) using the linear function between perceived severity and the midpoint values, at fixed $\overline{a_0} = 0.6$ and $\overline{b_0} = 0.75$. The first row, where $\sigma^2_{b_0} = 0$, and the first column, where $\sigma^2_{a_0} = 0$, present results analyzed in previous figures, with the disappearance of the second infection peak and the amplification of the first peak with increasing variance. The other plots show that the influences of behavioral heterogeneities for the two transitions are cumulative. Specifically, when both variances are non-zero, the second peak is absent, and increasing either one of the variances heightens the first peak. Thus, in this scenario with high values of the midpoints, the spread is accelerated by individuals with low perceived severity, who relax their behaviors early and fail to readopt them even when ICU occupancy is high. The increased compliance of groups with high perceived severity groups and their prompt readoption of measures is not enough to counterbalance this effect.

Similar results are observed for the metrics describing the severe disease outcomes. Figure S25 presents the results for the three metrics as a function of $\overline{a_0}$ and $\sigma^2_{a_0}$ with $\overline{b_0} = 0.75$ and $\sigma^2_{b_0} = 0.1$. We observe trends analogous to the ones found in the main text with $\sigma^2_{b_0} = 0$.

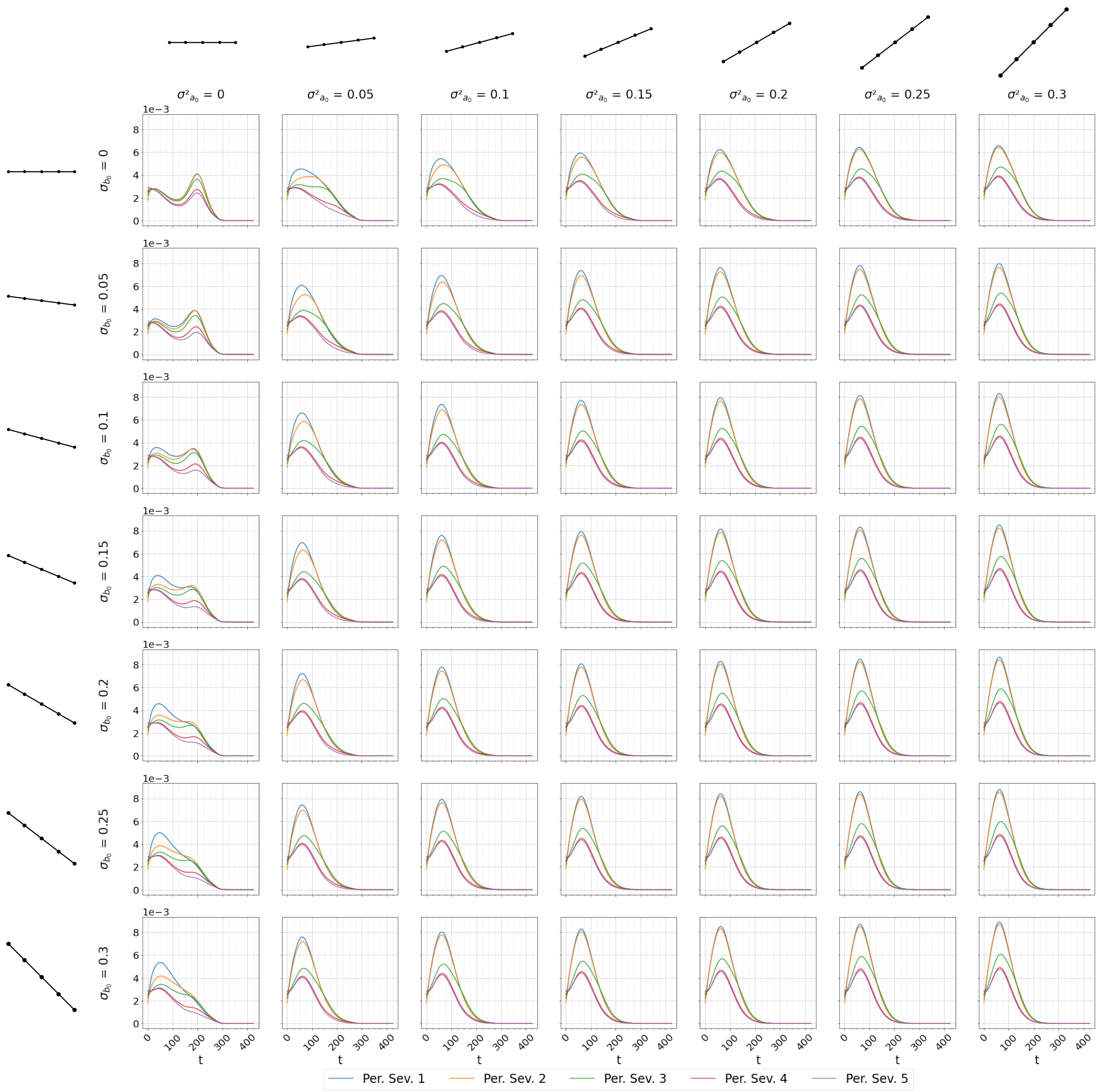

Figure S24: Fraction of infected individuals as a function of time (days) for each perceived severity group when both transitions depend on perceived severity. Each row corresponds to a different value of the variance $\sigma^2_{a_0}$, while each column corresponds to a different value of the variance $\sigma^2_{b_0}$, both going from 0 to 0.3. The other parameters used for the simulations are $\alpha = 10$, $\gamma = 5$, $\overline{a_0} = 0.6$, $\overline{b_0} = 0.75$, and we used the Linear function to model the dependency between the parameters and perceived severity.

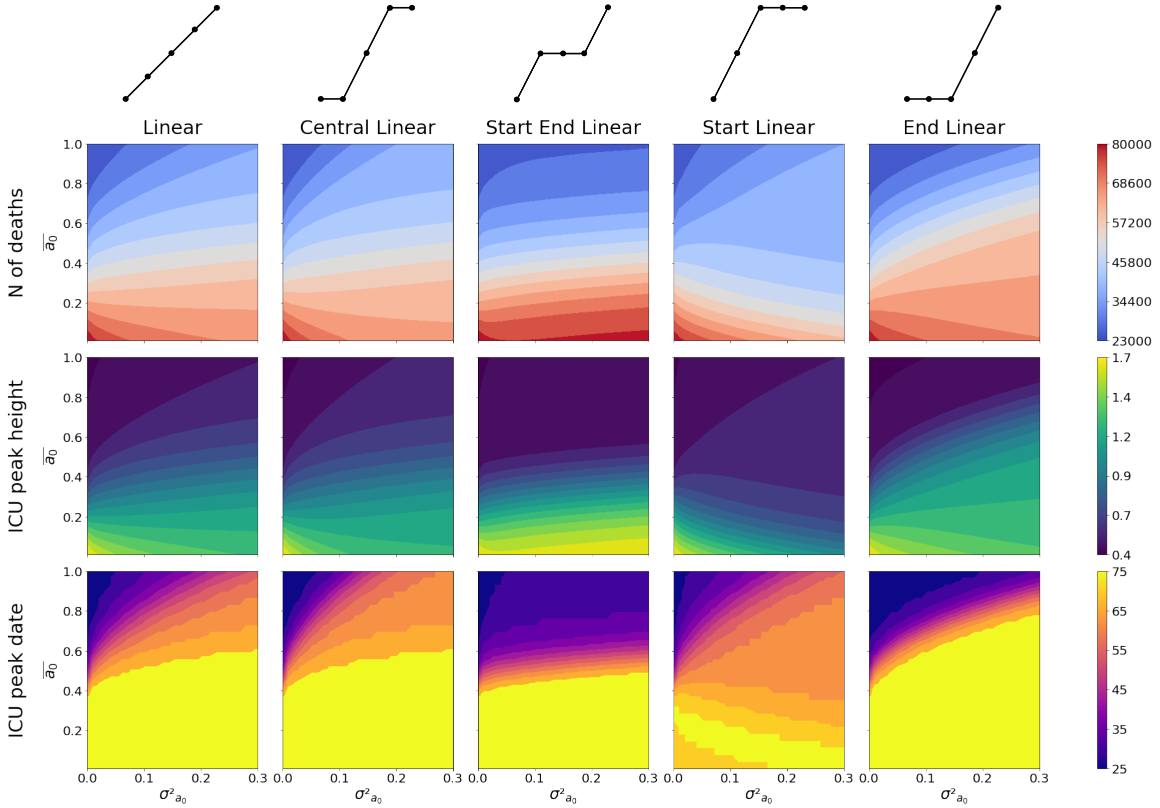

Figure S25: Heatmaps showing the number of deaths (first row), the height of the ICU peak (second row) and its date (third row) as a function of $\overline{a_0}$ and $\sigma^2_{a_0}$ with the transition NC to C depending on perceived severity as well ($\overline{b_0} = 0.75$ and $\sigma^2_{b_0} = 0.1$). Each column corresponds to one of the five functions linking perceived severity and midpoint of the logistic curve giving the transition rate from compliant to non-compliant compartments as a function of the fraction of vaccinated individuals. Above each column is a small diagram of the function, showing how the midpoint $a_0$ varies, going from small to high perceived severity groups (left to right). The other parameters used for the simulations are $\alpha = 10$ and $\gamma = 5$. We employed a 900-value grid, with 30 values of $\overline{a_0}$ ranging from 0 to 1, and 30 values of $\sigma^2_{a_0}$ ranging from 0 to 0.3. The rugged profile of the curves related to the peak date is due to its discrete nature, with values representing the (integer) number of days after the simulation's start in which the peak is observed.

#### S4 Sensitivity Analysis - Epidemiological parameters

In the previous section, we investigated how changing behavioral parameters affected the outcomes of the model. Here, we explore different scenarios where we change the parameters related to the epidemic spread and to the vaccination campaign. In particular, we examine the impact of different vaccine efficacies, initial conditions, permanence in ICU compartments, maximum number of ICU beds. We finally consider a different COVID-19 variant.

##### S4.1 Vaccine efficacy

In the main text, we have considered a very effective vaccine, with values taken from the literature, and a high protection against transmission and severe symptoms, obtained from a specific combination of $VE_S$, $VE_{Symp}$, and $VE_D$. We have first considered the case with the same high vaccine efficacy (90%) but different values of $VE_S$, $VE_{Symp}$, and $VE_D$. Specifically, we put in turn one of these values to 0. We obtained results very similar to the ones shown in the main text. We also obtain very similar results in a scenario where we switched the values of $VE_S$ and $VE_D$ with respect to the main text with $VE_S = 0.4$ and $VE_D = 0.7$. We also considered different rates of vaccination campaign, with either a slower ($r_v = 0.1$) or a faster ($r_v = 0.4$) roll-out of the vaccines. The results are not affected, which is mainly due to the fact that in our model the compliance with protective behaviors is linked with the progress of the vaccination campaign. Therefore, to a slower rate of vaccination corresponds also a slower relaxation of behaviors, while a faster deployment of the vaccines causes a much prompt non compliance. For this reason, the results remain similar.

In an hypothetical new pandemic or for a different strain however, the vaccine efficacy can be lower than the one considered in the main text. Here we examine scenarios in which the vaccine has a total efficacy of 70%, resulting from an efficacy against infection $VE_S = 0.4$, an efficacy against symptoms $VE_{Symp} = 0.3$, and an efficacy against severe outcomes $VE_D = 0.3$.

In Figure S26, we present the three metrics as a function of $\overline{a_0}$ and $\sigma^2_{a_0}$ for a lower vaccine efficacy of 70%. Compared to the results obtained with a vaccine efficacy of 90% shown in the main text, the number of deaths and the height of the ICU peak can reach higher values (100,000 versus 80,000 and 1.8 versus 1.6, respectively). Additionally, the metrics change less strongly when the variance is increased. This can be attributed to the reduced protection offered by the vaccine, resulting in a more significant impact of the second peak of infections on ICU admissions and fatalities when $\sigma^2_{a_0} = 0$. Consequently, for small values of the variance, we observe much higher metric values compared to the scenario with an overall vaccine efficacy of 90%. Conversely, for high variances, the metrics are less affected by the decrease in vaccine effectiveness because most infections occur early, when most individuals are not yet protected. These observations are confirmed in Figure S27, where we note a higher second peak in the ICU curves for $\sigma^2_{a_0} = 0$ compared to the corresponding plot in the main text. Consequently, in this scenario, we observe a transition in the dynamics from two peaks to one, not only for the curve of the infected population but also for the curve giving the temporal evolution of ICU occupancy. A very similar scenario (not shown) is obtained for an even lower total vaccine efficacy of 55% (obtained with $VE_S = 0.3$, $VE_{Symp} = 0.2$, and $VE_D = 0.2$), resulting in even higher numbers of deaths and second peaks of infected and ICU occupancy.

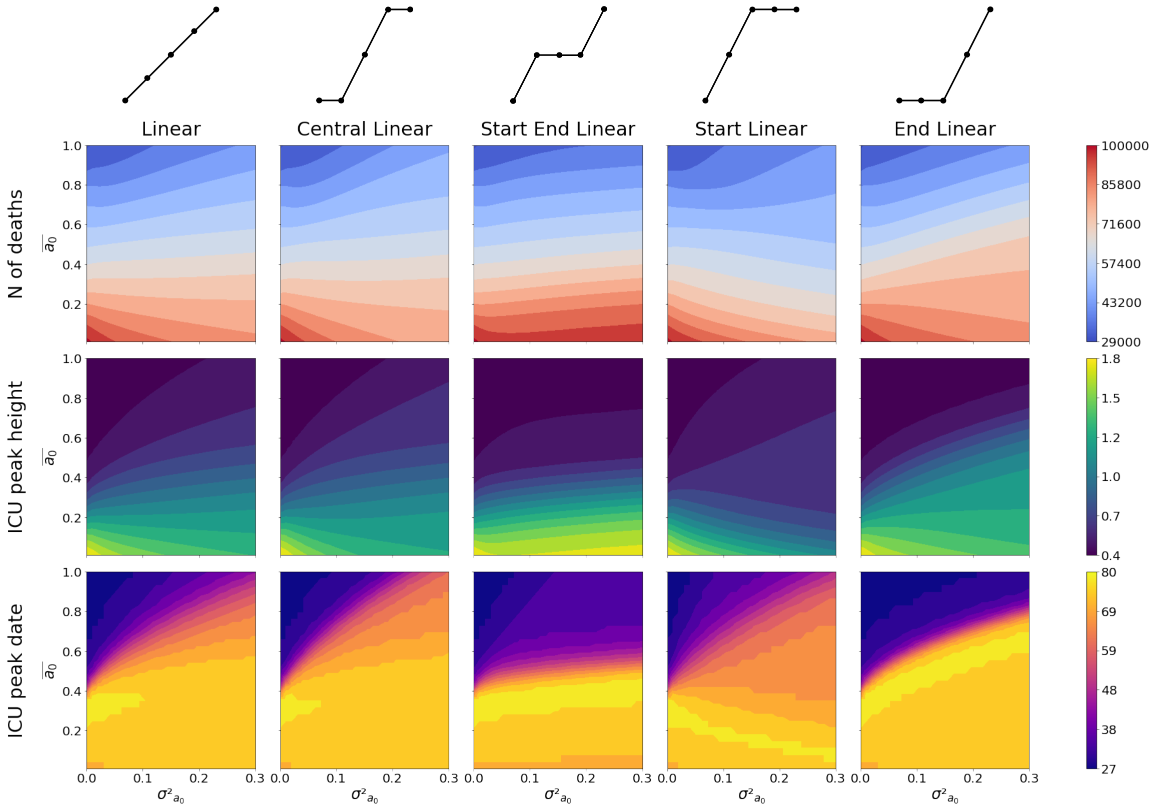

Figure S26: Heatmaps showing the number of deaths (first row), the height of the ICU peak (second row) and its date (third row) as a function of $\overline{a_0}$ and $\sigma^2_{a_0}$ with a lower total vaccine efficacy of 70% (given by $VE_S = 0.4$, $VE_{Symp} = 0.3$, and $VE_D = 0.3$). Each column corresponds to one of the five functions linking perceived severity and midpoint of the logistic curve giving the transition rate from compliant to non-compliant compartments as a function of the fraction of vaccinated individuals. Above each column is a small diagram of the function, showing how the midpoint $a_0$ varies, going from small to high perceived severity groups (left to right). The other parameters used for the simulations are $\alpha = 10$, $\gamma = 5$, $\overline{b_0} = 0.75$, and $\sigma^2_{b_0} = 0$. We employed a 900-value grid, with 30 values of $\overline{a_0}$ ranging from 0 to 1, and 30 values of $\sigma^2_{a_0}$ ranging from 0 to 0.3. The rugged profile of the curves related to the peak date is due to its discrete nature, with values representing the (integer) number of days after the simulation's start in which the peak is observed.

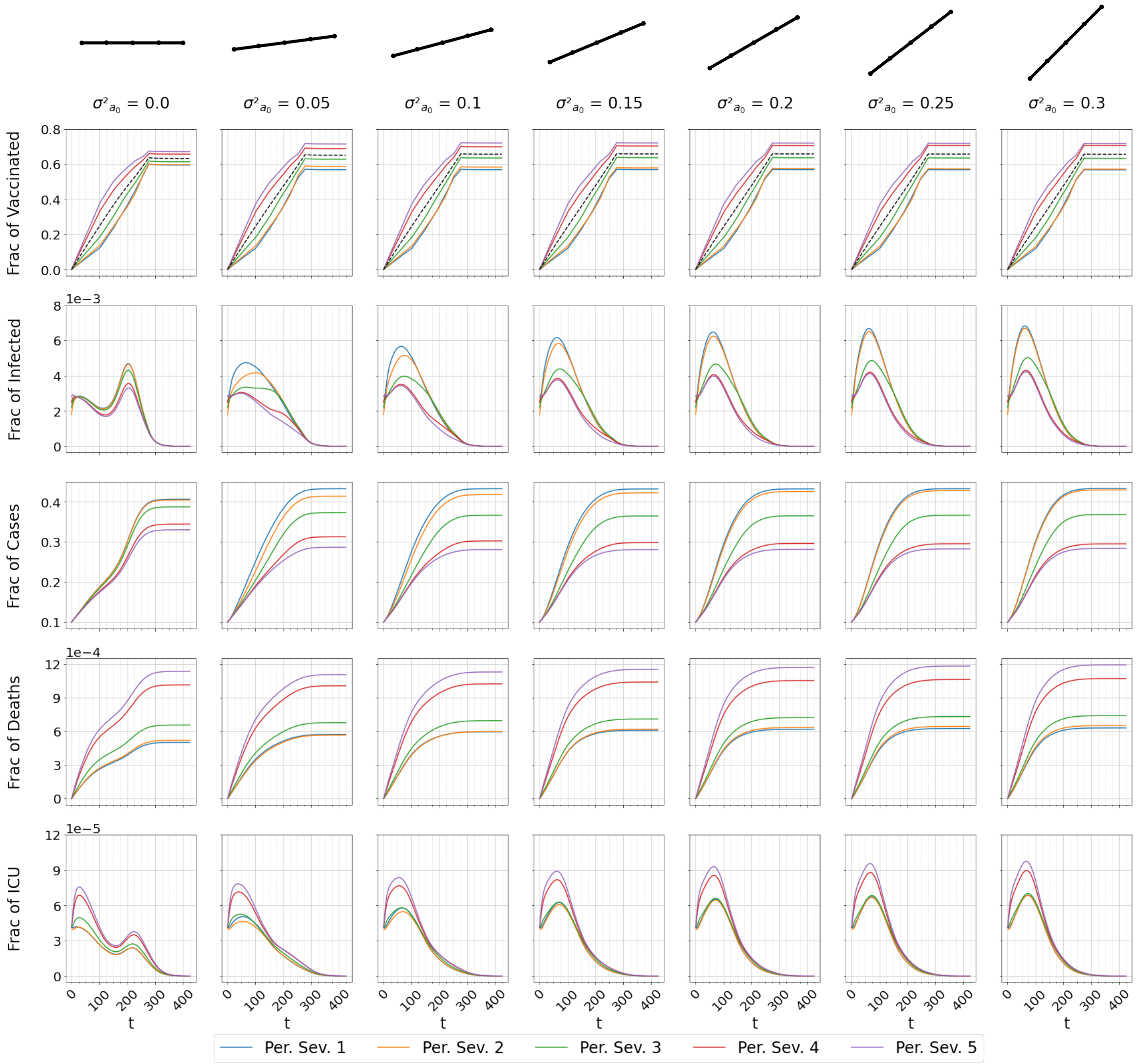

Figure S27: Fraction of vaccinated individuals (first row - the black dashed line reports the global fraction of vaccinated individuals in the population), infected individuals (second row), cases (third row - obtained as the sum of recovered individuals and deaths), deaths (fourth row) and individuals in ICU (fifth row) as a function of time (days), for each perceived severity group. Each column corresponds to a different value of the variance $\sigma^2_{a_0}$, going from 0 to 0.3, with the total vaccine efficacy reduced to 70% (given by $VE_S = 0.4$, $VE_{Symp} = 0.3$, and $VE_D = 0.3$). The other parameters are $\alpha = 10$, $\gamma = 5$, $\overline{a_0} = 0.6$, $\overline{b_0} = 0.75$, and $\sigma^2_{b_0} = 0$.

#### S4.2 Initial conditions

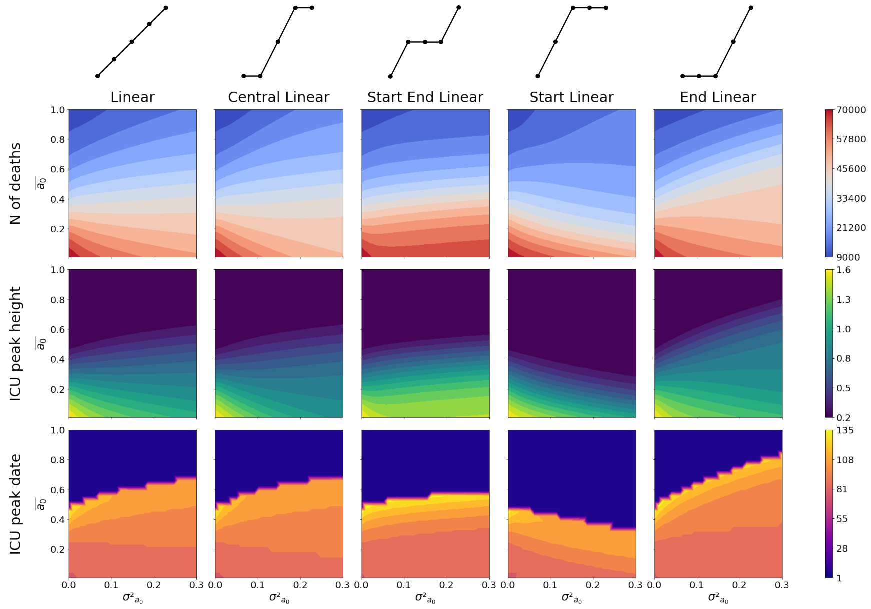

Figure S28: Heatmaps showing the number of deaths (first row), the height of the ICU peak (second row) and its date (third row) as a function of $\overline{a_0}$ and $\sigma^2_{a_0}$ with an initial number of individuals in the infected compartments $i_0 = 200,000$. Each column corresponds to one of the five functions linking perceived severity and midpoint of the logistic curve giving the transition rate from compliant to non-compliant compartments as a function of the fraction of vaccinated individuals. Above each column is a small diagram of the function, showing how the midpoint $a_0$ varies, going from small to high perceived severity groups (left to right). The other parameters used for the simulations are $\alpha = 10$, $\gamma = 5$, $\overline{b_0} = 0.75$, and $\sigma^2_{b_0} = 0$. We employed a 900-value grid, with 30 values of $\overline{a_0}$ ranging from 0 to 1, and 30 values of $\sigma^2_{a_0}$ ranging from 0 to 0.3. The rugged profile of the curves related to the peak date is due to its discrete nature, with values representing the (integer) number of days after the simulation's start in which the peak is observed.

In Figure S28, we present heatmaps depicting the three metrics with simulations starting from a smaller number of individuals distributed in the infected compartments, with $i_0 = 200,000$. We then observe a reduced number of deaths and ICU cases. The lower initial infected population, coupled with high initial compliance to safety measures, diminishes severe outcomes. This effect is particularly evident for high values of $\overline{a_0}$: when behavior relaxation occurs, the population is already protected, and the limited number of infected individuals does not significantly spread the virus.

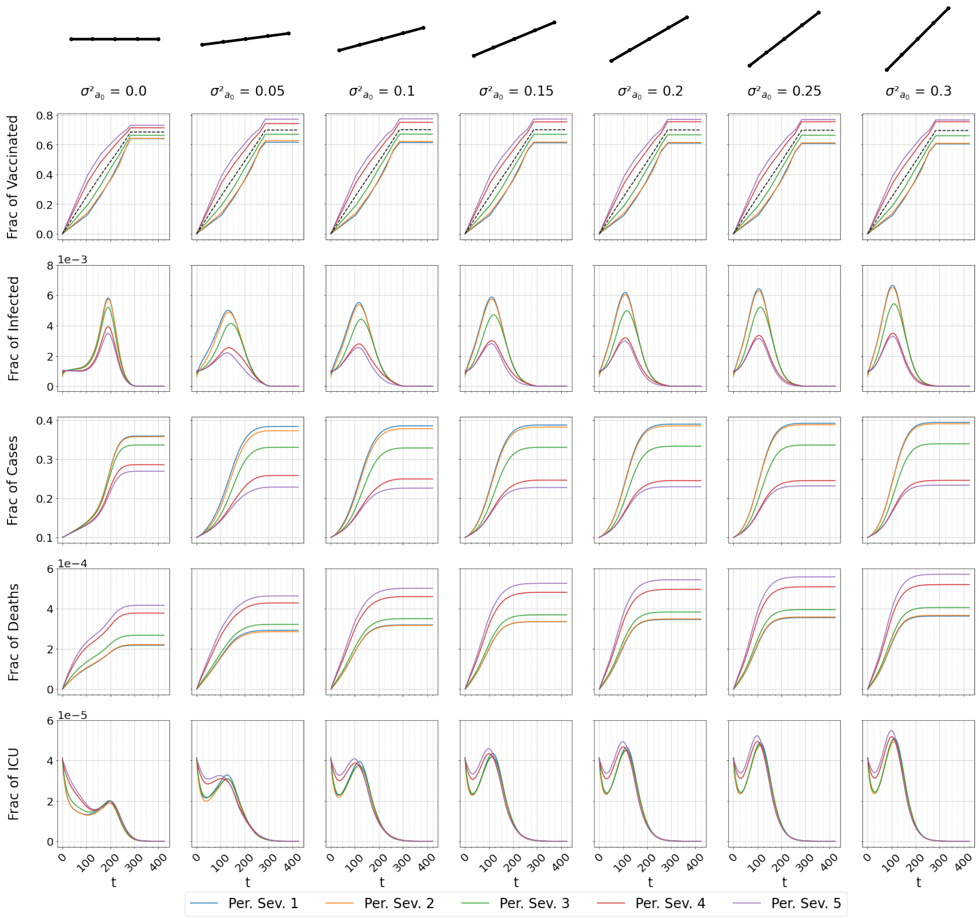

Figure S29: Fraction of vaccinated individuals (first row - the black dashed line reports the global fraction of vaccinated individuals in the population), infected individuals (second row), cases (third row - obtained as the sum of recovered individuals and deaths), deaths (fourth row) and individuals in ICU (fifth row) as a function of time (days), for each perceived severity group. Each column corresponds to a different value of the variance $\sigma^2_{a_0}$, going from 0 to 0.3, with the initial number of individuals in infected compartments reduced to $i_0 = 200000$. The other parameters are $\alpha = 10$, $\gamma = 5$, $\overline{a_0} = 0.6$, $\overline{b_0} = 0.75$, and $\sigma^2_{b_0} = 0$.

The epidemiological curves in Figure S29, obtained for $\overline{a_0} = 0.6$, show that in this case, the first peak in the fraction of infected is in fact absent for $\sigma^2_{a_0} = 0$. Consequently, the fraction of ICU occupancy initially decreases, followed by a minor peak corresponding to the rise in infected individuals due to the relaxation of behaviors. However, this peak's height does not surpass the initial number of ICU individuals. With increasing variance, contagion shifts towards the early pandemic phase due to premature relaxation among groups with low perceived severity, leading to a growing ICU peak that eventually surpasses $icu_0$ for high variances.

Note that we do not show here the scenario with a higher initial number of infected individuals ($i_0 = 1,000,000$), as the dynamics remain unchanged compared to those described in the main text, albeit with slight metric increases.

#### S4.3 Permanence in ICU: $\Delta$

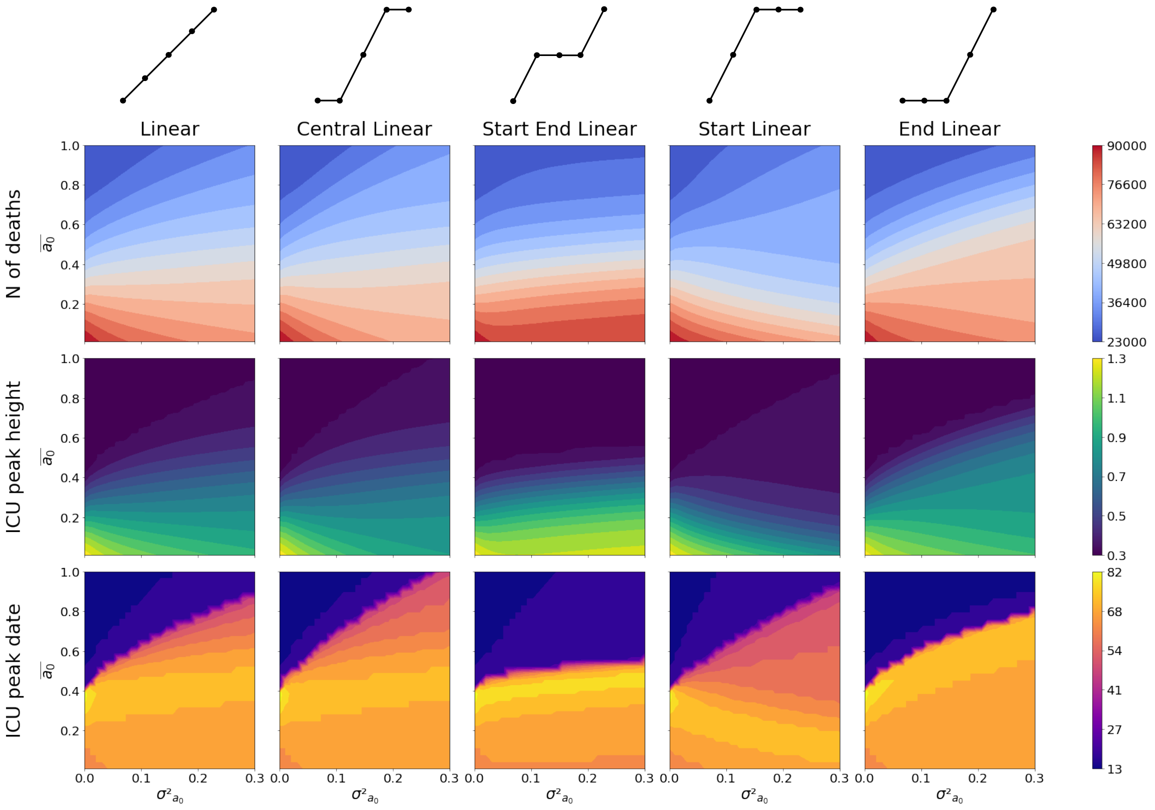

Figure S30: Heatmaps showing the number of deaths (first row), the height of the ICU peak (second row) and its date (third row) as a function of $\overline{a_0}$ and $\sigma^2_{a_0}$ with a shorter mean days of permanence in ICU compartment $\Delta = 10$ days. Each column corresponds to one of the five functions linking perceived severity and midpoint of the logistic curve giving the transition rate from compliant to non-compliant compartments as a function of the fraction of vaccinated individuals. Above each column is a small diagram of the function, showing how the midpoint $a_0$ varies, going from small to high perceived severity groups (left to right). The other parameters used for the simulations are $\alpha = 10$, $\gamma = 5$, $\overline{b_0} = 0.75$, and $\sigma^2_{b_0} = 0$. We employed a 900-value grid, with 30 values of $\overline{a_0}$ ranging from 0 to 1, and 30 values of $\sigma^2_{a_0}$ ranging from 0 to 0.3. The rugged profile of the curves related to the peak date is due to its discrete nature, with values representing the (integer) number of days after the simulation's start in which the peak is observed.

In this subsection, we investigate a scenario where we reduce the duration of stay in ICU compartments, decreasing $\Delta$ from 15 to 10 days. In Figure S30, we display the three metrics as functions of $\overline{a_0}$ and $\sigma^2_{a_0}$. The numbers of deaths are slightly higher than those for $\Delta = 15$ days, while ICU peak heights are smaller. This occurs because the shorter occupancy of ICU beds helps prevent significant hospital saturation, but in return, the transition rate from NC to C is lower. A decreased occupancy means individuals are less likely to revert to compliant behavior compared to the scenario analyzed in the main text.

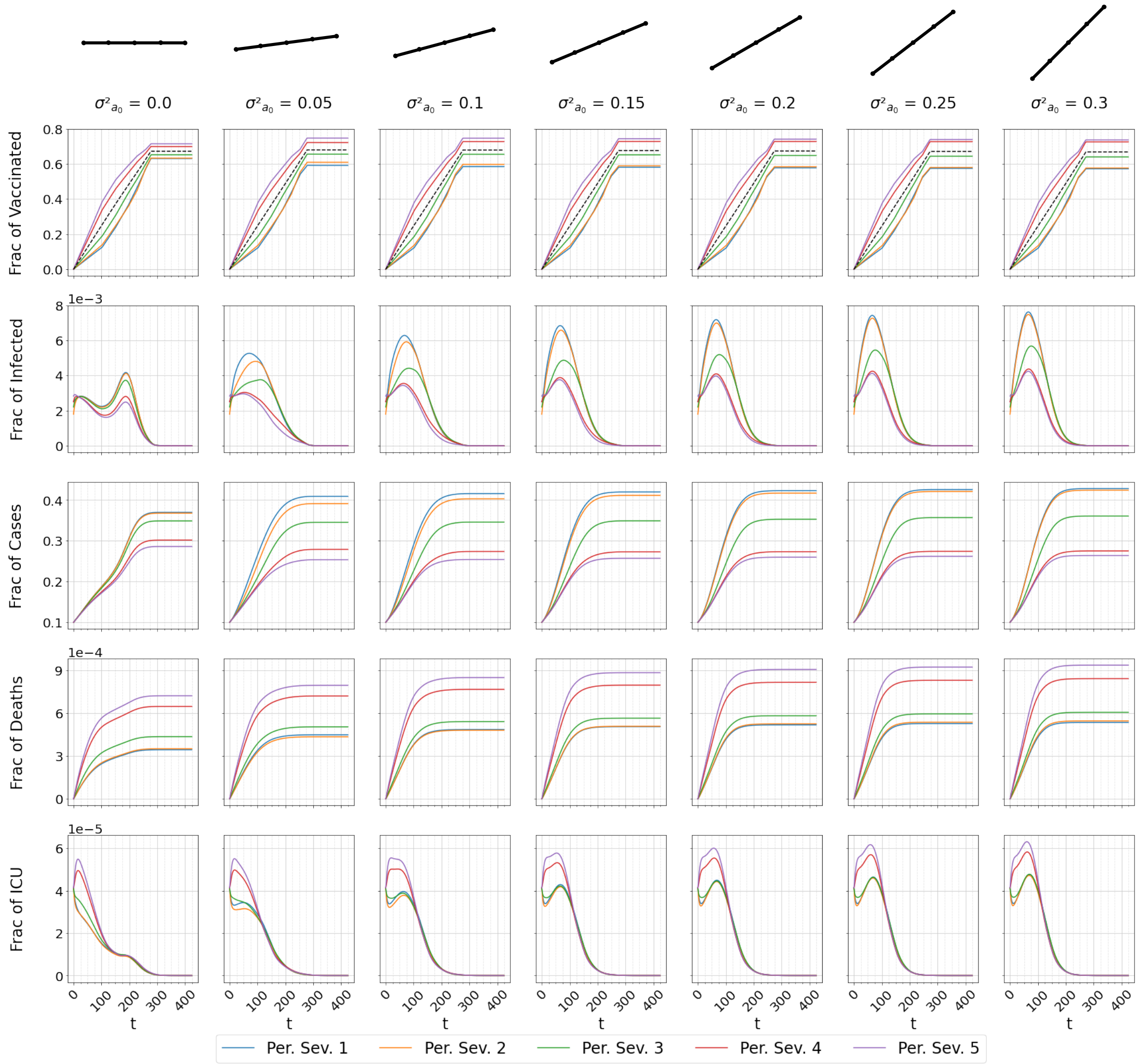

Figure S31: Fraction of vaccinated individuals (first row - the black dashed line reports the global fraction of vaccinated individuals in the population), infected individuals (second row), cases (third row - obtained as the sum of recovered individuals and deaths), deaths (fourth row) and individuals in ICU (fifth row) as a function of time (days), for each perceived severity group. Each column corresponds to a different value of the variance $\sigma^2_{a_0}$, going from 0 to 0.3, with the mean days of permanence in ICU compartments reduced to $\Delta = 10$ days. The other parameters are $\alpha = 10$, $\gamma = 5$, $\overline{a_0} = 0.6$, $\overline{b_0} = 0.75$, and $\sigma^2_{b_0} = 0$.

The epidemiological curves in Figure S31 display similar trends to those in the main text, with the transition from a double-peak profile to a single peak as variance increases.

The scenario with a longer ICU stay ($\Delta = 20$ days), not shown here, exhibits a dynamic similar to that of $\Delta = 15$ days, with a comparable number of deaths and a higher ICU peak.

#### S4.4 Available number of ICU beds: $ICU_{max}$

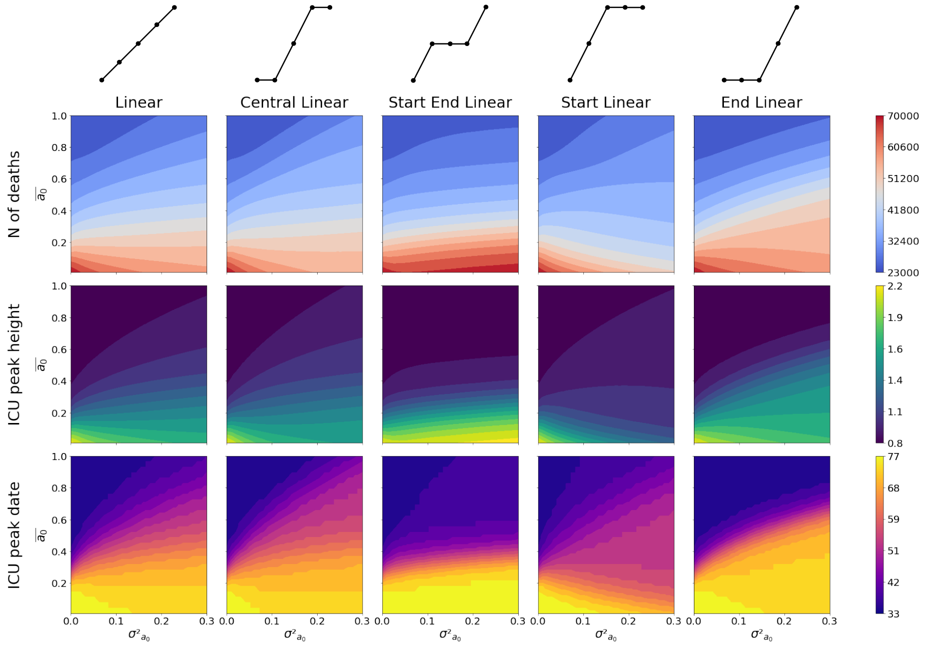

Figure S32: Heatmaps showing the number of deaths (first row), the height of the ICU peak (second row) and its date (third row) as a function of $\overline{a_0}$ and $\sigma^2_{a_0}$ with a smaller number of beds available in ICU $ICU_{max} = 5,000$. Each column corresponds to one of the five functions linking perceived severity and midpoint of the logistic curve giving the transition rate from compliant to non-compliant compartments as a function of the fraction of vaccinated individuals. Above each column is a small diagram of the function, showing how the midpoint $a_0$ varies, going from small to high perceived severity groups (left to right). The other parameters used for the simulations are $\alpha = 10$, $\gamma = 5$, $\overline{b_0} = 0.75$, and $\sigma^2_{b_0} = 0$. We employed a 900-value grid, with 30 values of $\overline{a_0}$ ranging from 0 to 1, and 30 values of $\sigma^2_{a_0}$ ranging from 0 to 0.3. The rugged profile of the curves related to the peak date is due to its discrete nature, with values representing the (integer) number of days after the simulation's start in which the peak is observed.

In Figure S32, we present the results of the three metrics in a scenario where the number of available beds is the same as at the beginning of the pandemic in Spring 2020, with around 5,000 beds. As expected, the height of the ICU peak shows an increase compared to the case of $ICU_{max} = 7,200$, reaching more than twice the maximum occupancy for some combinations of $\overline{a_0}$ and $\sigma^2_{a_0}$. However, it is interesting to note that the number of deaths is similar to or slightly smaller than those in the main article. This can be explained by the earlier return to compliant compartments by individuals who relaxed their behaviors. Indeed, the limited availability of beds accelerates the transition from NC to C. Therefore, as soon as ICU occupancy begins to rise, individuals promptly revert to safer behaviors, reducing their contacts.

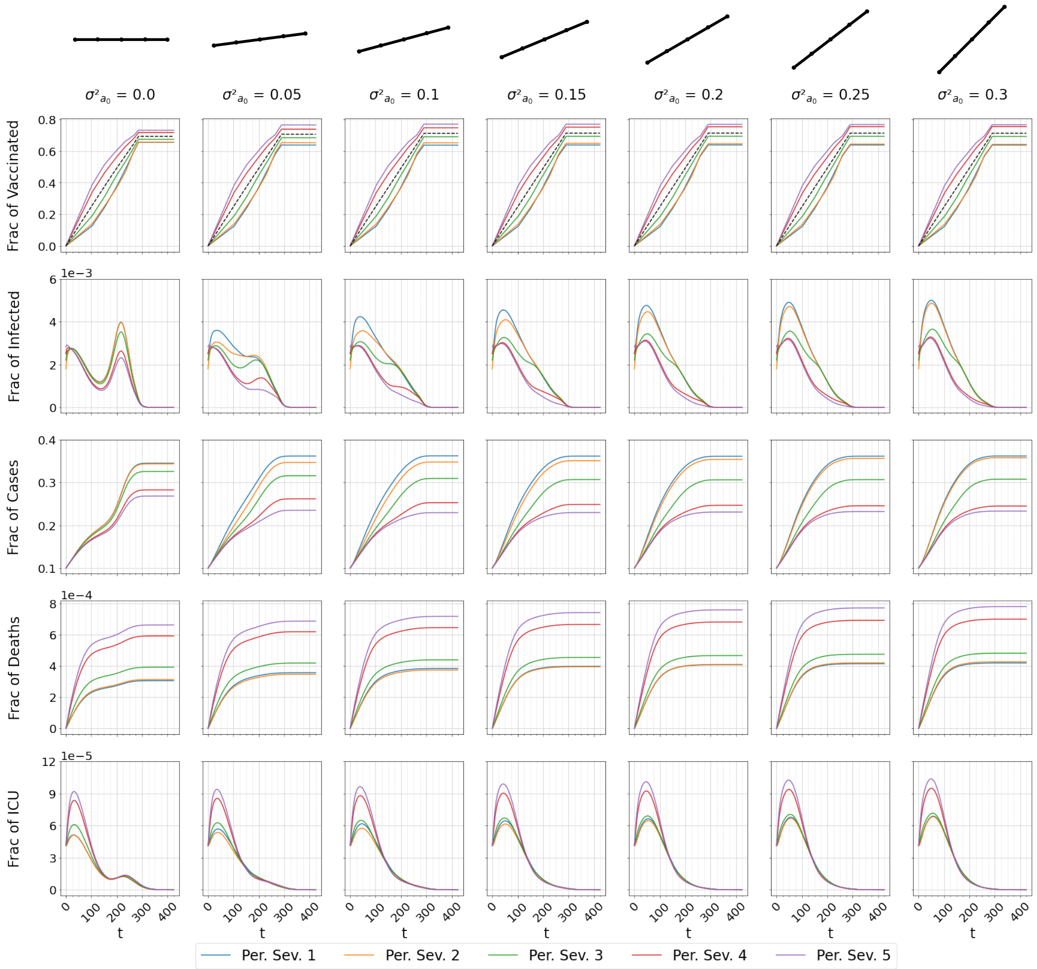

Figure S33: Fraction of vaccinated individuals (first row - the black dashed line reports the global fraction of vaccinated individuals in the population), infected individuals (second row), cases (third row - obtained as the sum of recovered individuals and deaths), deaths (fourth row) and individuals in ICU (fifth row) as a function of time (days), for each perceived severity group. Each column corresponds to a different value of the variance $\sigma^2_{a_0}$, going from 0 to 0.3, with the available number of beds in ICU reduced to $ICU_{max} = 5000$. The other parameters are $\alpha = 10$, $\gamma = 5$, $\overline{a_0} = 0.6$, $\overline{b_0} = 0.75$, and $\sigma^2_{b_0} = 0$.

In Figure S33, we show the epidemiological curves. The trends are similar to those observed with a higher $ICU_{max}$, particularly for the fraction of individuals in ICU.

Increasing $ICU_{max}$ to $10,000$ causes almost no change in dynamics or metrics. The higher number of beds and the consequent increased non-compliance due to the smaller rate from NC to C compensate, resulting in numbers similar to those obtained for $ICU_{max} = 7200$.

#### S4.5 Alpha variant

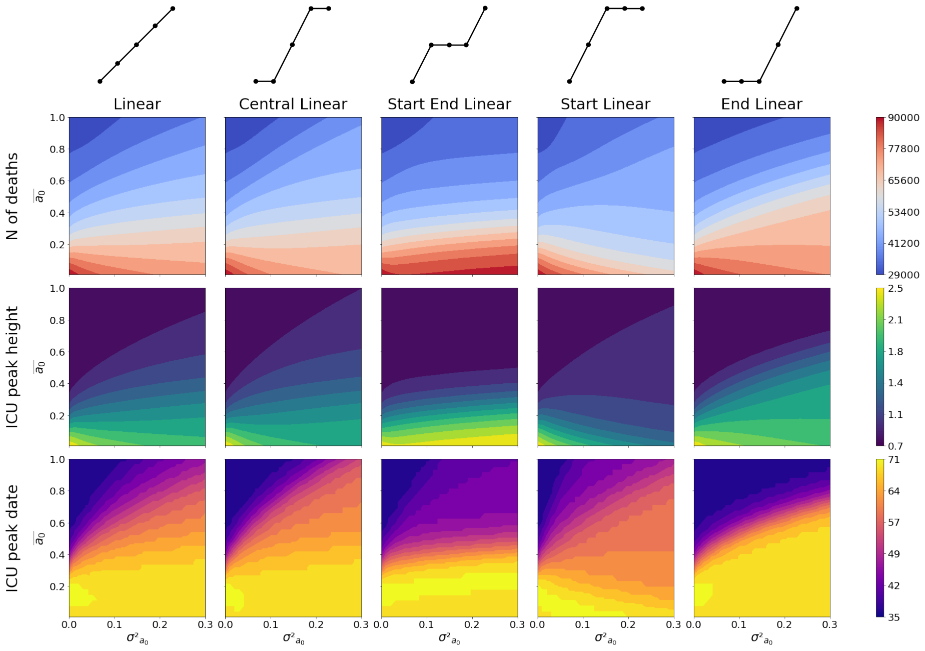

Figure S34: Heatmaps showing the number of deaths (first row), the height of the ICU peak (second row) and its date (third row) as a function of $\overline{a_0}$ and $\sigma^2_{a_0}$ using the epidemiological parameters of the alpha variant $\epsilon^{-1} = 2.9$ days and $\omega^{-1} = 2.3$ days. Each column corresponds to one of the five functions linking perceived severity and midpoint of the logistic curve giving the transition rate from compliant to non-compliant compartments as a function of the fraction of vaccinated individuals. Above each column is a small diagram of the function, showing how the midpoint $a_0$ varies, going from small to high perceived severity groups (left to right). The other parameters used for the simulations are $\alpha = 10$, $\gamma = 5$, $\overline{b_0} = 0.75$, and $\sigma^2_{b_0} = 0$. We employed a 900-value grid, with 30 values of $\overline{a_0}$ ranging from 0 to 1, and 30 values of $\sigma^2_{a_0}$ ranging from 0 to 0.3. The rugged profile of the curves related to the peak date is due to its discrete nature, with values representing the (integer) number of days after the simulation's start in which the peak is observed.

We finally simulated the spread of the virus in a scenario where the vaccination campaign occurs with the Alpha variant being the dominant Variant of Concern (VoC). Therefore, we used a shorter average latent period of $\epsilon^{-1} = 2.9$ days but a longer average pre-symptomatic period of $\omega^{-1} = 2.3$ days, sourced from the same references used for the Delta variant values. From Figure S34 and the presented metrics, we observe a very similar phenomenology as for the Delta variant, despite an increase in both the height of the ICU peak and in the number of deaths (reaching 110,000 individuals compared to the 80,000 of the Delta variant). However, the robustness of our model is confirmed by the epidemiological curves depicted in Figure S35, which show almost no difference in dynamics compared to those obtained for the Delta variant.

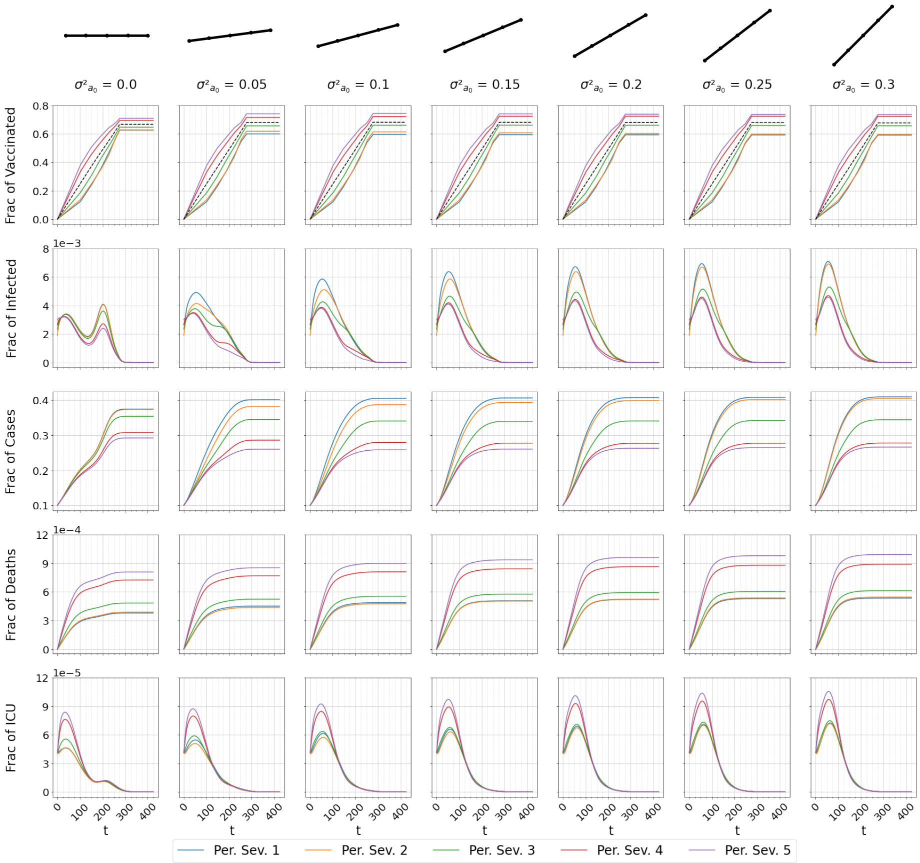

Figure S35: Fraction of vaccinated individuals (first row - the black dashed line reports the global fraction of vaccinated individuals in the population), infected individuals (second row), cases (third row - obtained as the sum of recovered individuals and deaths), deaths (fourth row) and individuals in ICU (fifth row) as a function of time (days), for each perceived severity group. Each column corresponds to a different value of the variance $\sigma^2_{a_0}$, going from 0 to 0.3, and we used the epidemiological parameters of the alpha variant $\epsilon^{-1} = 2.9$ days and $\omega^{-1} = 2.3$ days. The other parameters are $\alpha = 10$, $\gamma = 5$, $\overline{a_0} = 0.6$, $\overline{b_0} = 0.75$, and $\sigma^2_{b_0} = 0$.

### References

[1] F. Verelst, L. Hermans, S. Vercruysse, A. Gimma, P. Coletti, J. A. Backer, K. L. Wong, J. Wambua, K. van Zandvoort, L. Willem, *et al.*, "Socrates-comix: a platform for timely and open-source contact mixing data during and in between covid-19 surges and interventions in over 20 european countries," *BMC medicine*, vol. 19, no. 1, pp. 1–7, 2021.

[2] E. Commodari and V. L. La Rosa, "Adolescents in quarantine during covid-19 pandemic in italy: perceived health risk, beliefs, psychological experiences and expectations for the future," *Frontiers in psychology*, vol. 11, p. 559951, 2020.
